# Correction of LMS Parameter Errors in Published Noonan Syndrome Growth Charts

**DOI:** 10.1101/2025.07.29.25332354

**Authors:** Joseph H Chou

**Affiliations:** Mass General Brigham for Children, Newborn Medicine Harvard Medical School, Boston MA

**Keywords:** Noonan syndrome, growth chart, LMS parameters

## Abstract

Growth reference curves were recently published by Cappa et al 2024 for children with Noonan Syndrome, including the LMS parameters which should allow calculation of expected values at any centile. However, closer inspection revealed significant problems with the LMS parameters, including calculated values that diverged to infinity or to imaginary numbers. In this brief report, I describe issues found, recalculate the LMS parameters based on the provided expected centile values, and quantify the improvement in performance and identify where the greatest discrepancies were found between the published and re-estimated LMS values. It is hoped that the Cappa et al will provide updated and corrected parameters to allow more confident use of this important clinical resource.

## Introduction

Growth reference curves for Noonan Syndrome were recently developed from a 15-year single center study of 190 participants, including charts for four measures (height velocity, weight, height, and BMI) for males and females^1^. The supplementary materials included expected values at a number of centiles as well as LMS parameters to mathematically recreate the curves at arbitrary centiles^2^.

During assessment of an electronic calculator created to facilitate use of these charts, some of the charts exhibited significant discrepancies between the expected values and values and those calculated from the LMS parameters.

This short technical brief describes the issues discovered and re-estimates LMS parameters from the reported centile data.

## Methods

Source data was obtained from the Supplementary Tables S1a through S1h^1^ and included for each measure, sex, and age the LMS parameters as well as expected measure values at the 3^rd^, 10^th^, 25^th^, 50^th^, 75^th^, 90^th^, and 97^th^ centiles. The LMS method^2^ allows calculation of measurement values X for any given L, M, S, and Z value. The parameters represent the Box-Cox power transformation for normalization of skew (L), the median value (M), and the coefficient of variation (S). Of note a Z score of 0 should represent the median (50^th^ percentile), as can be seen from the equations (Figure 1).

**Figure 1.**
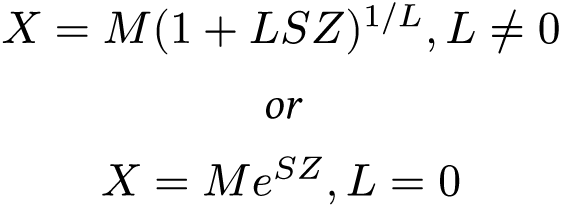
Calculation of growth measurement X from parameters L, M, S, and Z

Those same equations can also be used to re-estimate the LMS parameters, given values of X and Z. To estimate the three LMS parameters, a minimum of three points would be required – seven X / Z pairs were available (from the seven expected centiles) for each LMS parameter. Regression estimation of the LMS parameters was performed using the `minpack.lm` R package for non-linear least squares regression^3^. Some convenience functions for LMS calculations were taken from the `peditools` R package^4^.

## Results

### Discrepancies calculating values from LMS

The eight growth chart curves at the seven centile values were recreated by using the published LMS parameters (Figure 2 see also Supplementary Materials for larger versions of each panel). While most of the calculated values (red dots) were coincident with the expected centile values (black lines), there were discrepancies seen with the male BMI and weight curves, with the 97^th^ percentile male BMI curve approaching infinite (and then imaginary) BMI after 17 years, indicating likely problems with the LMS parameters themselves. Less dramatic but clearly visible, the 90^th^ percentile male BMI and higher percentile male weight curves all showed calculated values diverging away from the expected values.

**Figure 2.**
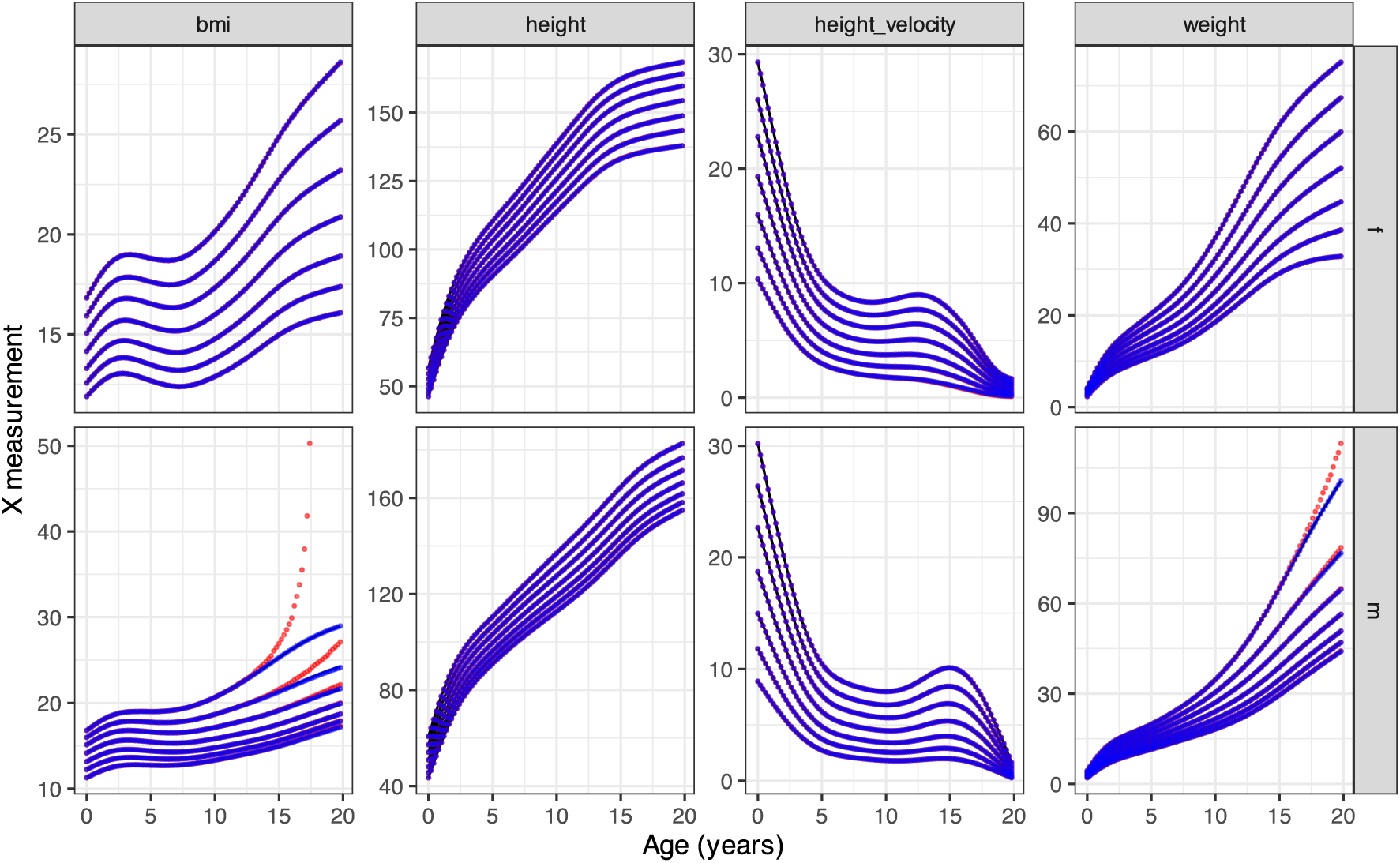
Discrepancies between reported and calculated values. The reported values for the 3rd, 10th, 25th, 50th, 75th, 90th, and 97th percentile curves are in black; red dots are calculated from the published LMS values; blue dots are calculated from the re-estimated LMS values.

By the definition of the LMS parameterization (Figure 1), a Z score of 0 should be identical to the M parameter. Plotting the difference between the M parameter and the reported 50^th^ percentile for each chart and both genders at all ages revealed internal inconsistencies, most notable for the male BMI and weight curves at higher ages, but also with the female height velocity curve, also at higher ages (Figure 3).

**Figure 3.**
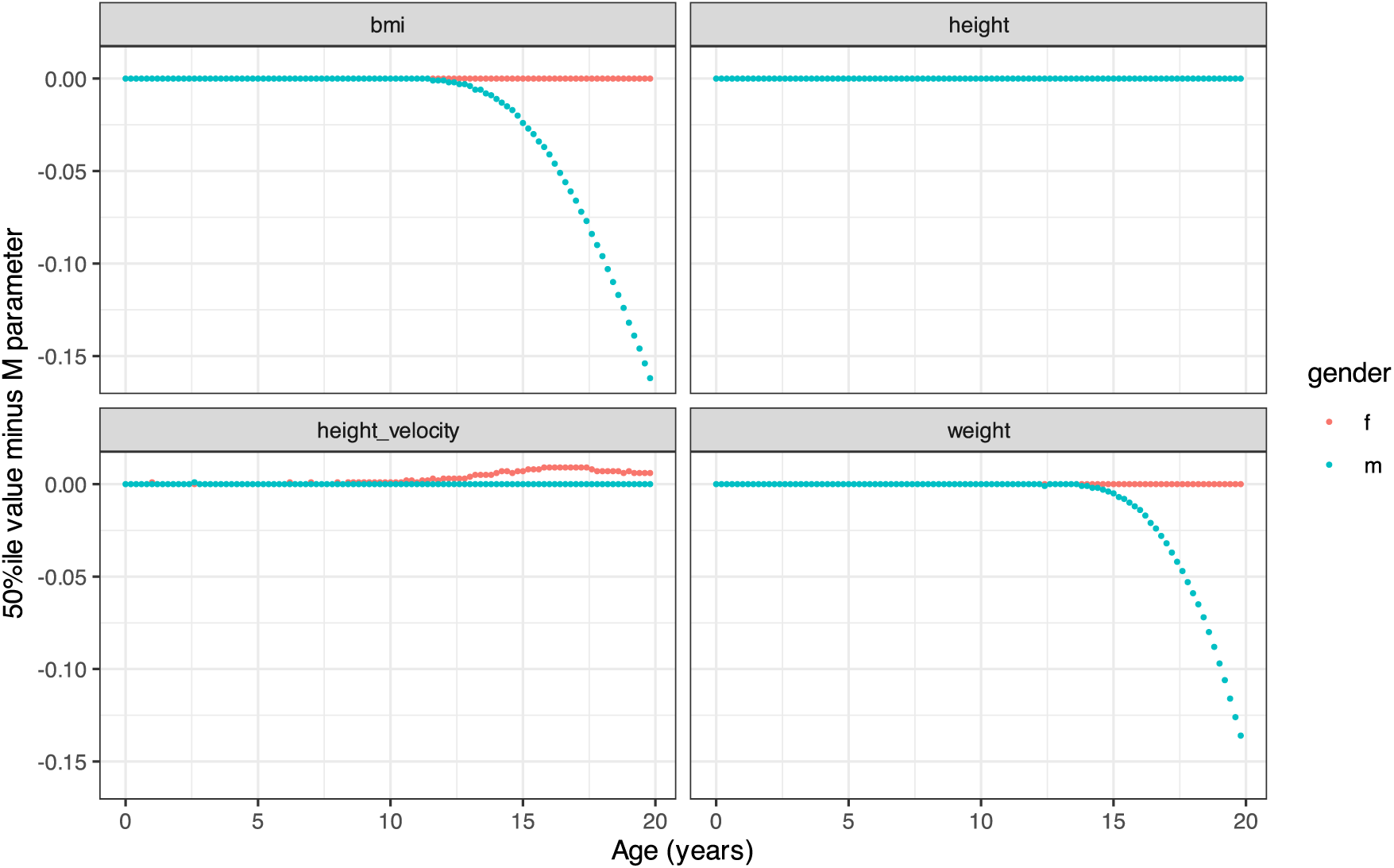
Difference between the reported 50th percentile value and the M parameter, which should be identical.

### Recalculation of LMS parameters based on reported centile values

Assuming that the reported centile values for all the curves were accurate (clearly, the male BMI curve approaching infinity was not accurate), the LMS parameters were re-estimated by least squares nonlinear regression from the seven available X / Z values from the centile curves, for all curves, ages, and sexes.

### Recreated growth curves

The re-estimated LMS values were then used to recreate each of the percentile curves; the blue dots of the calculated values from the re-estimated LMS values now nearly precisely recreate the reported centile values (Figure 2).

### Quantification of improved prediction

The mean absolute error (MAE) and maximum absolute error from the reported percentile values were also calculated for each of the calculated X values from both the originally published and the re-estimated LMS values (Table 1). For all charts, the re-estimated LMS parameters were non-inferior to the published LMS parameters at reproducing the expected values.

**Table 1.** Differences between reported values and values calculated from the published (`calc`) vs re-estimated (`est`) LMS values. The mean absolute value of the difference (MAE) and the maximum absolute of the differnece (MAX) are reported. The male BMI discrepancies from the published LMS values could not be calculated because of non-real number results.

| measure | gender | calcMAE | estMAE | calcMAX | estMAX |
| --- | --- | --- | --- | --- | --- |
| bmi (kg/m2) | f | 0.005 | 0.000 | 0.032 | 0.001 |
| bmi (kg/m2) | m | NaN | 0.013 | NaN | 0.132 |
| height (cm) | f | 0.034 | 0.000 | 0.137 | 0.001 |
| height (cm) | m | 0.032 | 0.000 | 0.213 | 0.001 |
| height_velocity (cm/year) | f | 0.008 | 0.002 | 0.080 | 0.010 |
| height_velocity (cm/year) | m | 0.002 | 0.000 | 0.013 | 0.001 |
| weight (kg) | f | 0.008 | 0.000 | 0.064 | 0.001 |
| weight (kg) | m | 0.177 | 0.012 | 12.574 | 0.215 |

### Comparison of published and re-estimated LMS parameters

The difference between the published LMS and the re-estimated LMS parameters was visualized (Figure 4). As expected, the greatest differences were seen in the plots with the greatest discrepancies seen in Figure 2 (red dots), but interestingly, the issues were not consistent for which LMS parameters were predominantly affected: the male BMI L, M, and S parameters; male weight L and M parameters; and female height velocity S parameters seemed to change most when re-estimated.

**Figure 4.**
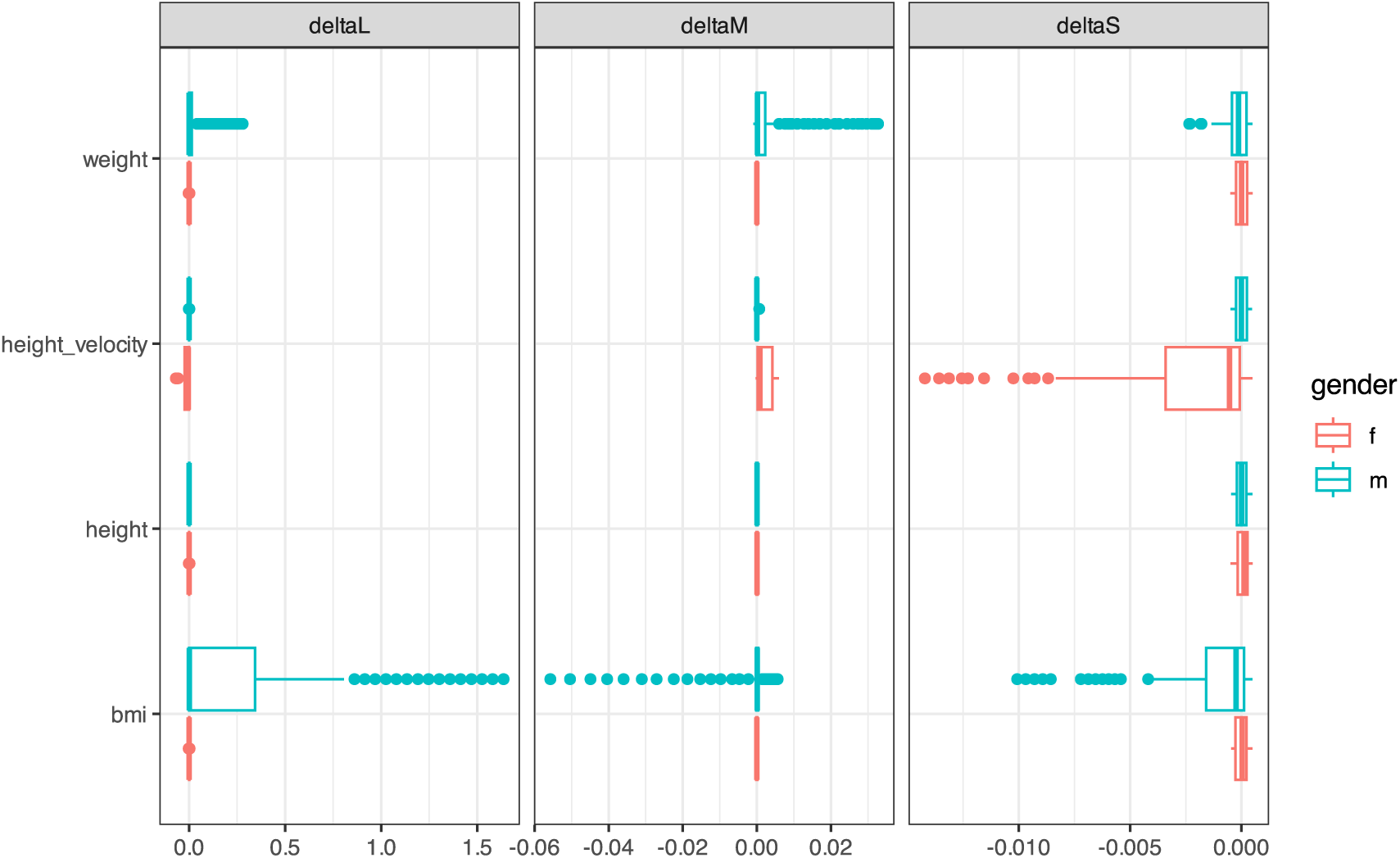
Differences between published and estimated LMS parameters.

## Discussion

The work of Cappa et al. addresses an important clinical need of evaluating expected growth outcomes in children with Noonan Syndrome, but there were significant inconsistencies between values calculated from the published LMS parameters and the expected percentile values, which compromise the clinical utility of these growth charts.

Most critically, the male BMI curve parameters produced mathematically impossible values (infinity and imaginary numbers) at higher ages and percentiles, with similar but less extreme divergence with the male weight curve. The M parameter diverged significantly from the 50th percentile values for male BMI and weight curves at higher ages, with milder divergence for the female height velocity curve.

To address these issues, nonlinear regression was used to independently estimate LMS parameters from the published expected percentile values in the supplementary materials. The re-estimated LMS parameters predicted the expected percentiles with substantially improved accuracy.

Given that growth charts are clinical tools that could directly impact patient care decisions, I believe it is important to bring these data inconsistencies to the attention of the medical community. Incorrect LMS parameters could lead to misinterpretation of growth patterns in children with Noonan syndrome. I attempted to contact the corresponding author via email in December 2024 and February 2025 to discuss these findings, but received no response. My hope is that the authors consider providing corrected supplementary materials to ensure the clinical utility and accuracy of this important resource.

## Data Availability

All data produced in the present study are available upon reasonable request to the authors

## Supplementary Materials

### R code to estimate LMS parameters

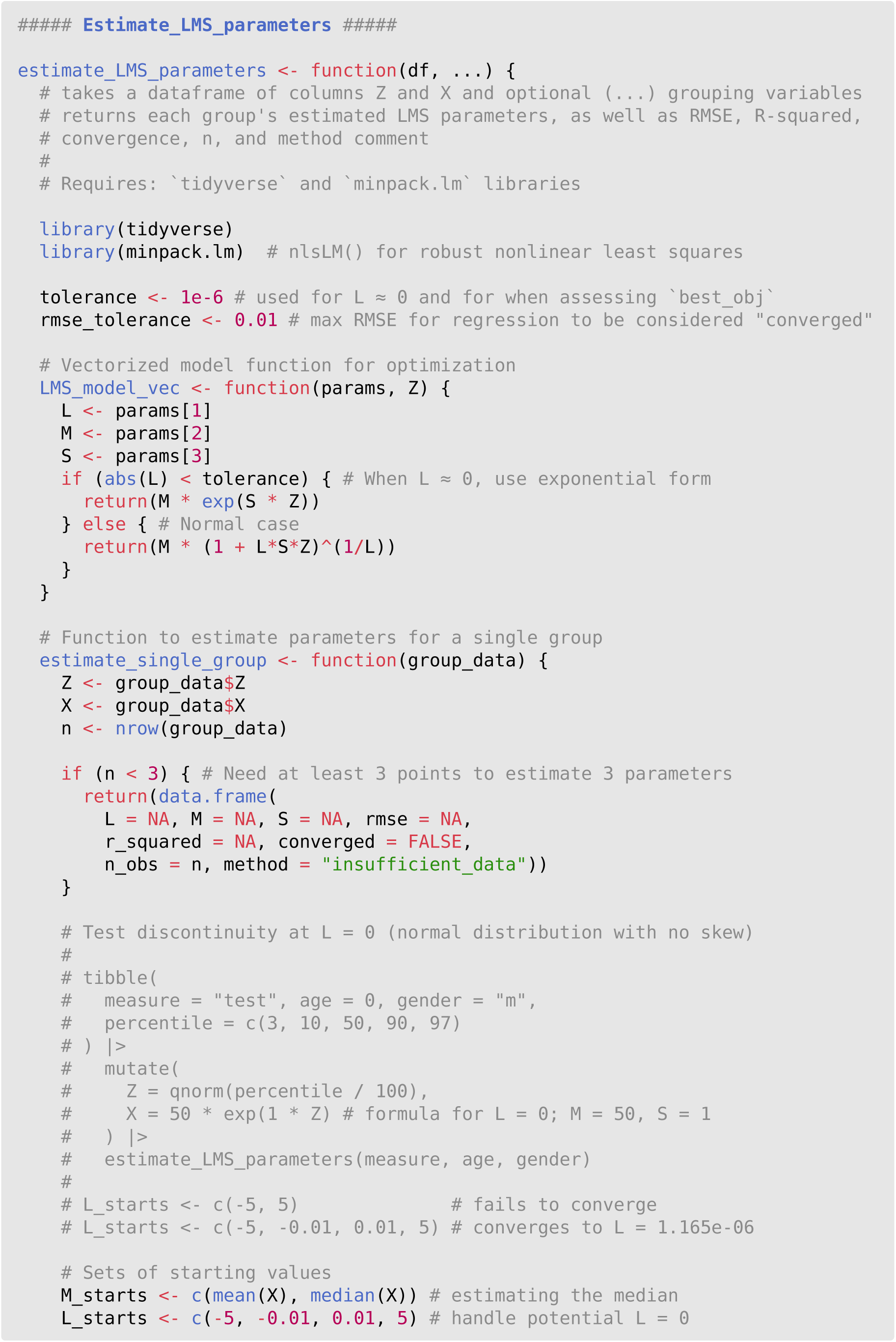
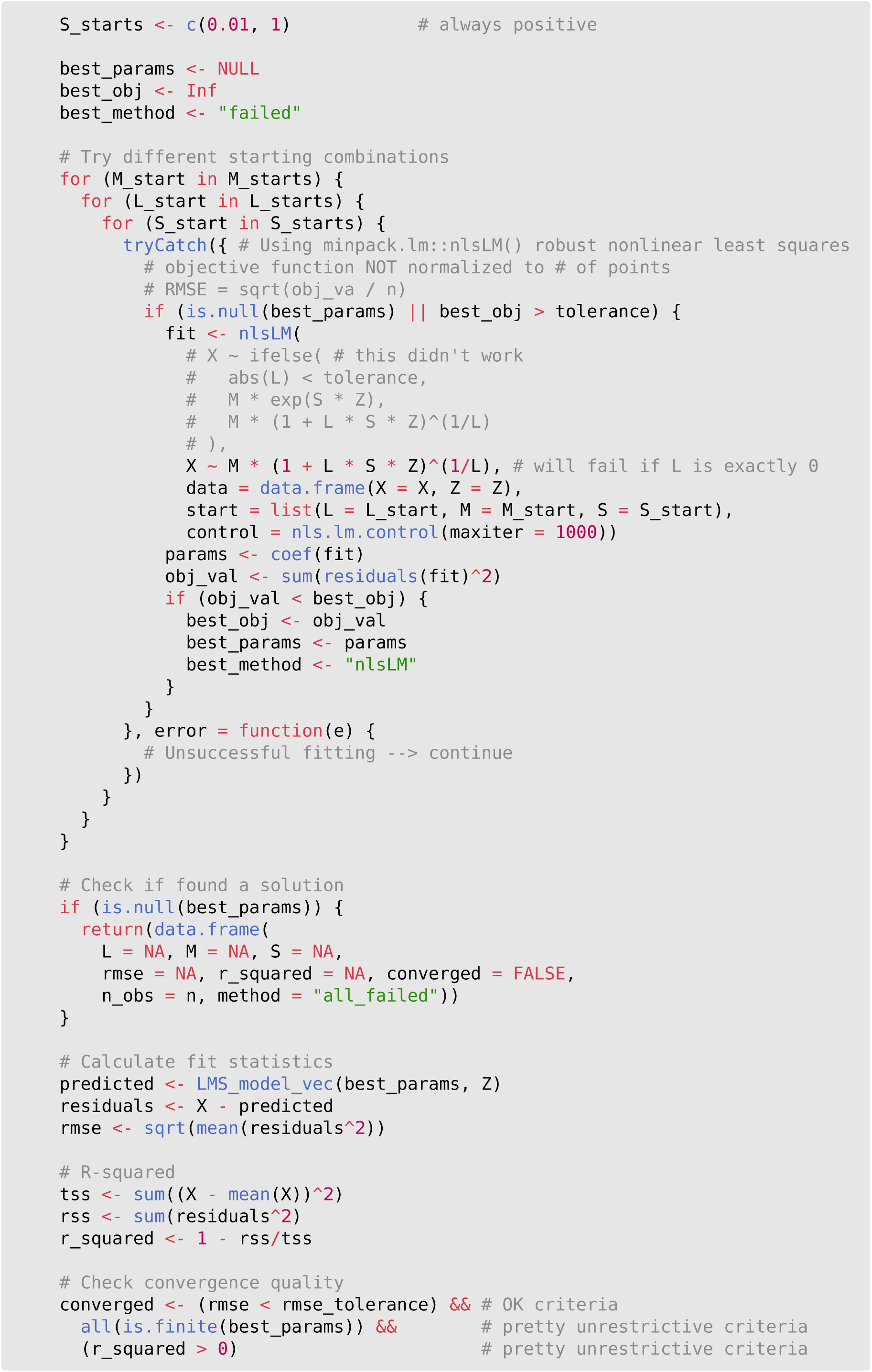
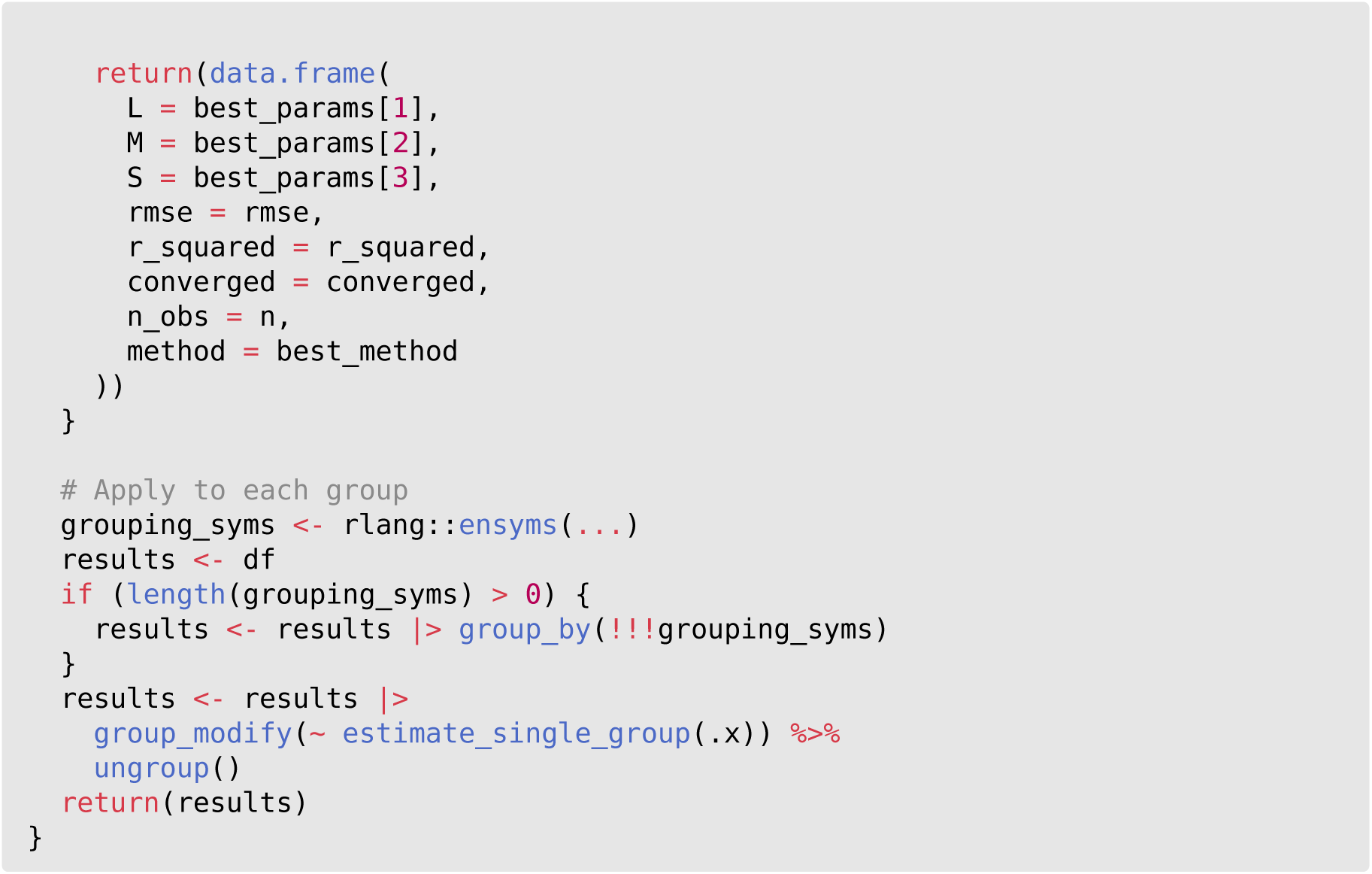

### Example of estimate_LMS_parameters() usage

Starting data

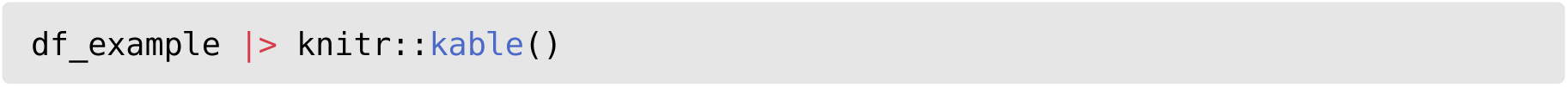

| measure | gender | age | p_3 | p_10 | p_25 | p_50 | p_75 | p_90 | p_97 |
| --- | --- | --- | --- | --- | --- | --- | --- | --- | --- |
| height | f | 0.0 | 46.274 | 47.570 | 48.995 | 50.731 | 52.657 | 54.582 | 56.696 |
| height | f | 0.2 | 49.346 | 50.733 | 52.255 | 54.108 | 56.160 | 58.207 | 60.450 |
| height | m | 0.0 | 43.485 | 45.636 | 48.002 | 50.882 | 54.062 | 57.218 | 60.648 |
| height | m | 0.2 | 46.518 | 48.748 | 51.199 | 54.177 | 57.461 | 60.717 | 64.250 |

Convert to long format and percentiles to Z scores

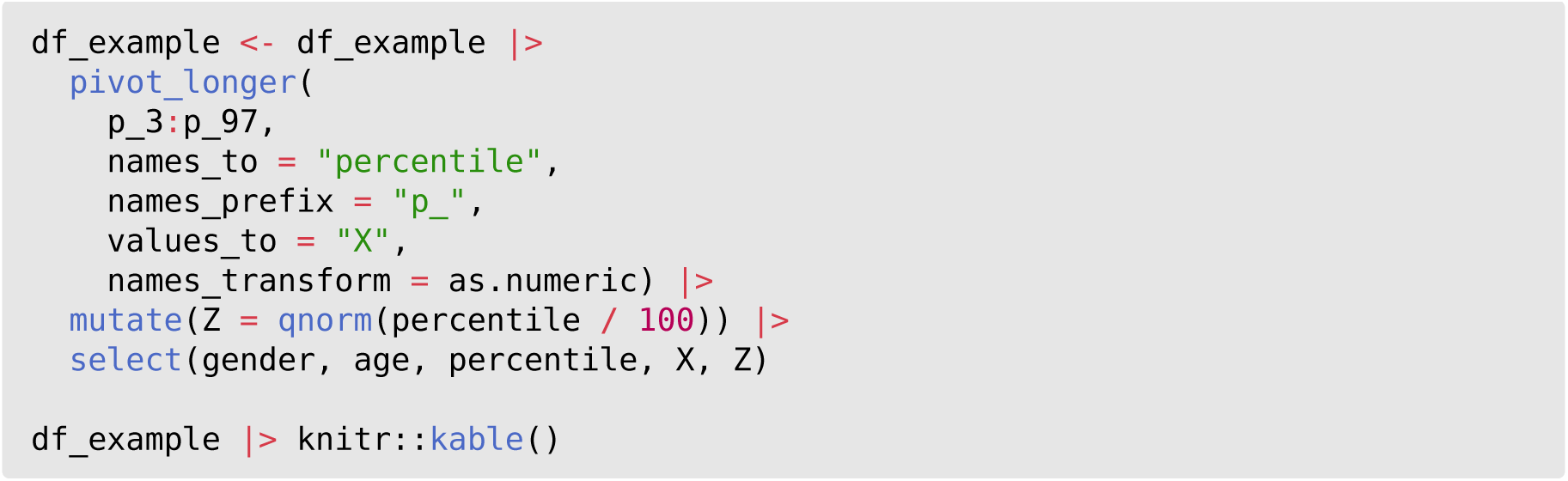

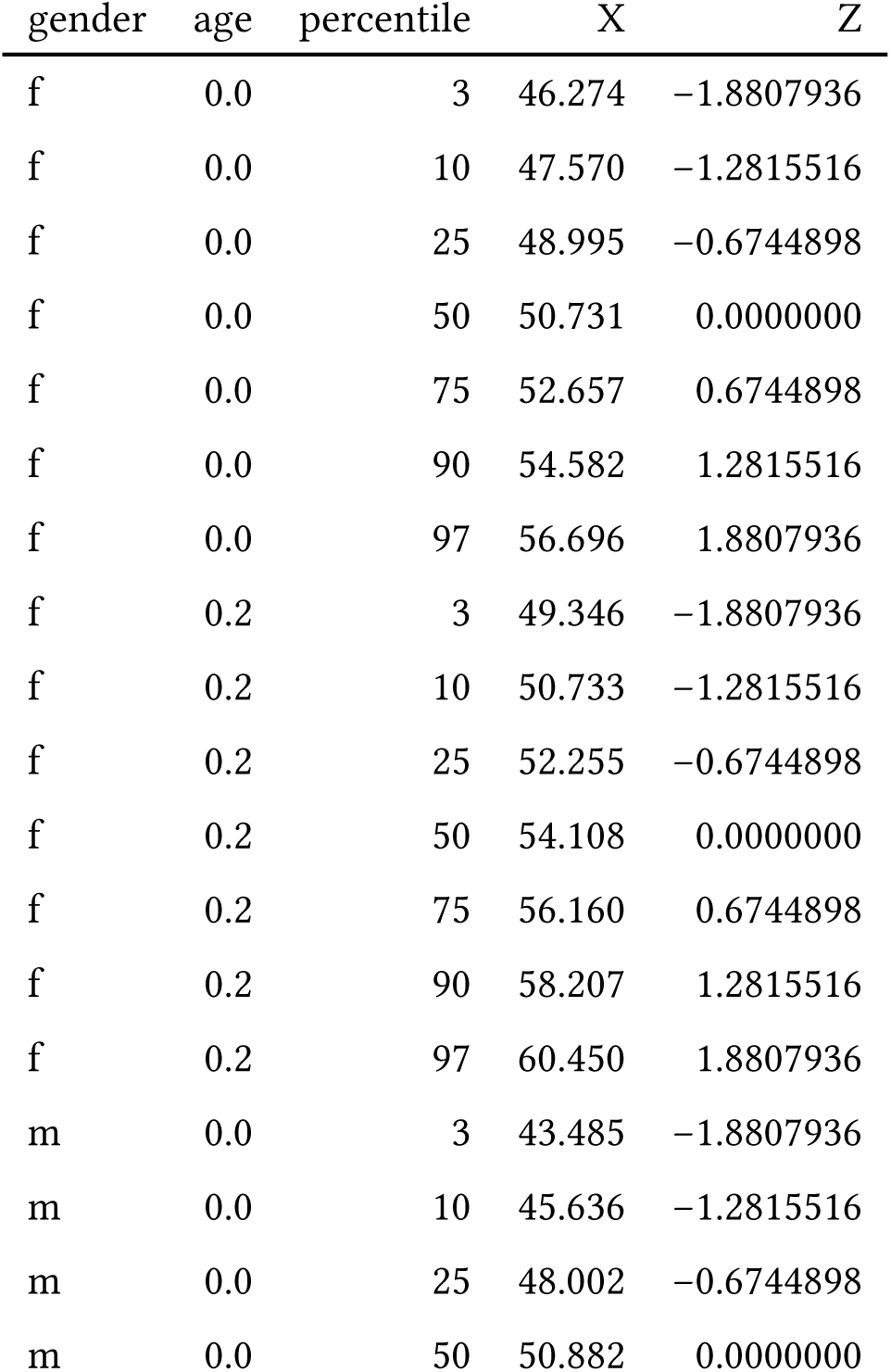

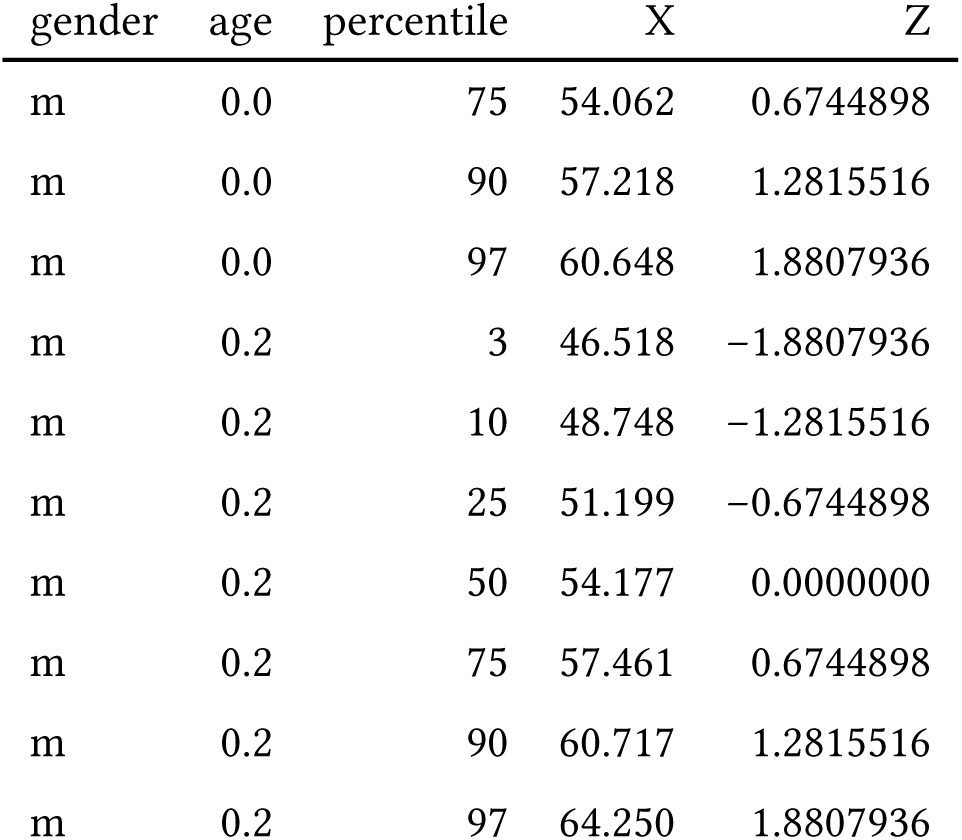

Estimate LMS parameters

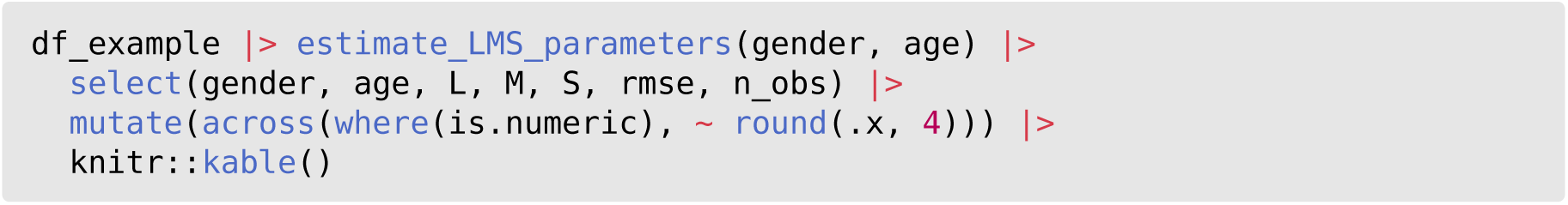

| gender | age | L | M | S | rmse | n_obs |
| --- | --- | --- | --- | --- | --- | --- |
| f | 0.0 | -1.8734 | 50.7311 | 0.0534 | 0.0002 | 7 |
| f | 0.2 | -1.8272 | 54.1081 | 0.0533 | 0.0003 | 7 |
| m | 0.0 | -0.6696 | 50.8818 | 0.0881 | 0.0002 | 7 |
| m | 0.2 | -0.6960 | 54.1769 | 0.0855 | 0.0001 | 7 |

### Individual plots

Individual plots from each panel from Figure 2. Discrepancies between reported and calculated values. The reported values for the 3^rd^, 10^th^, 25^th^, 50^th^, 75^th^, 90^th^, and 97^th^ percentile curves are in black; red dots are calculated from the published LMS values; blue dots are calculated from the re-estimated LMS values.

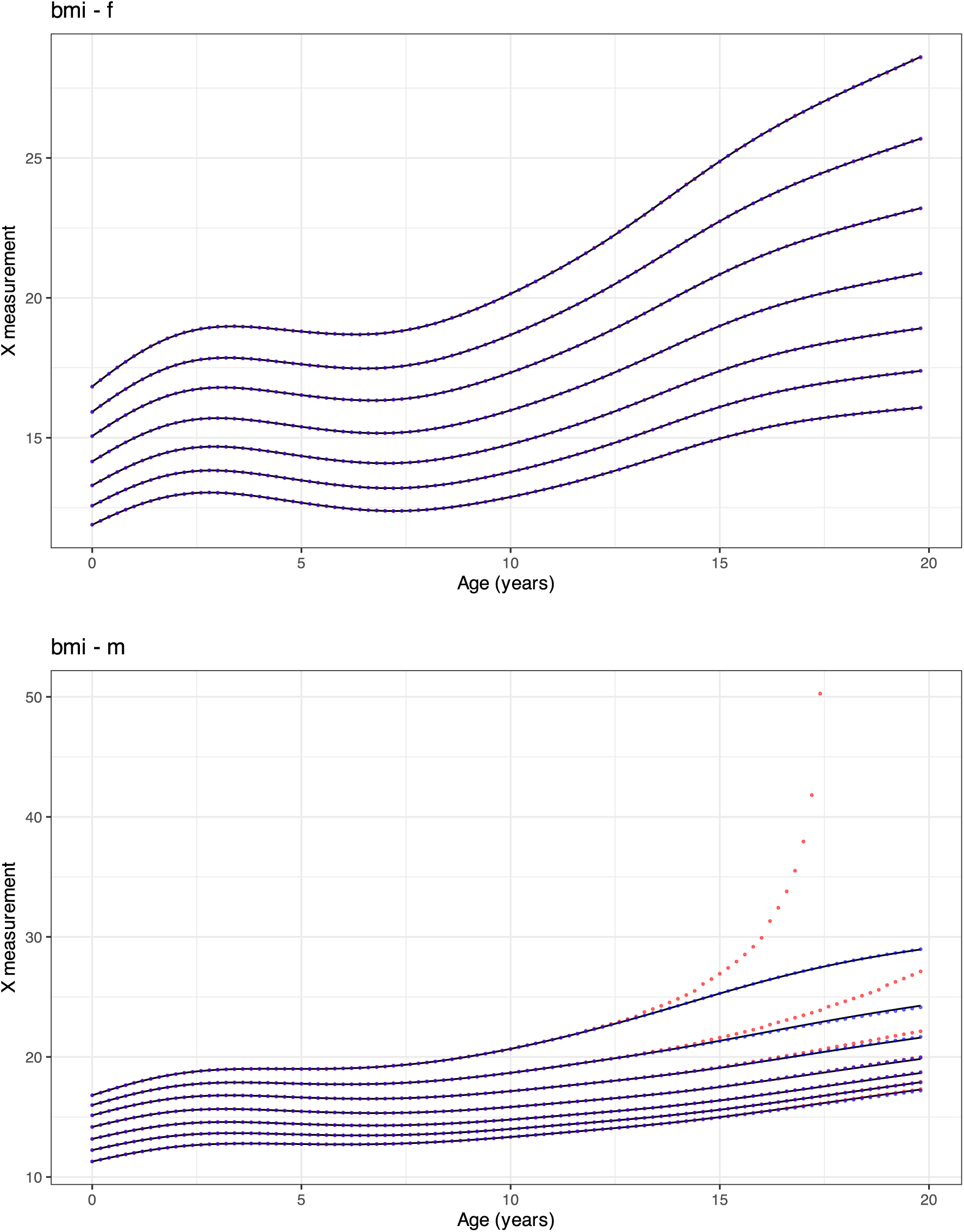
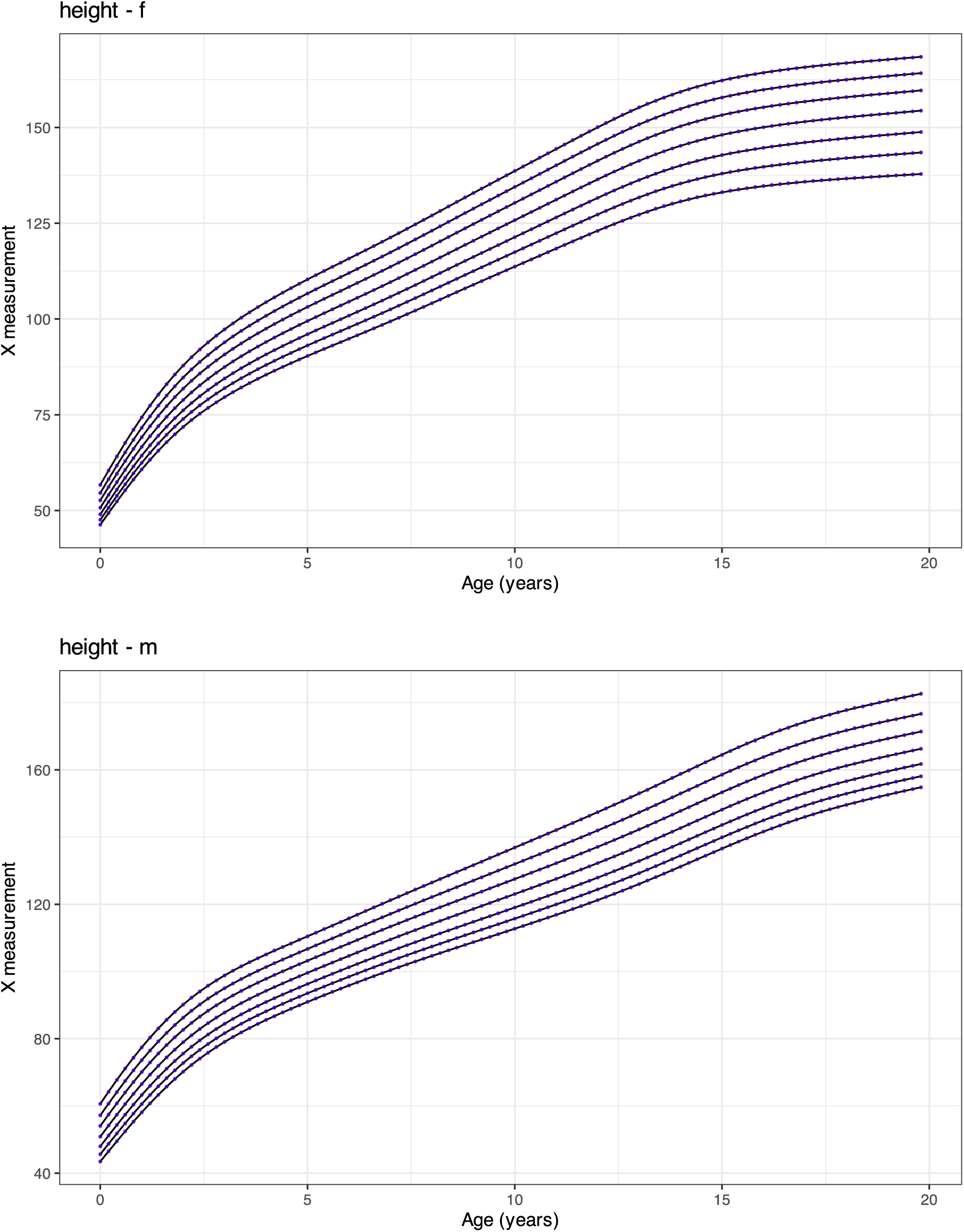
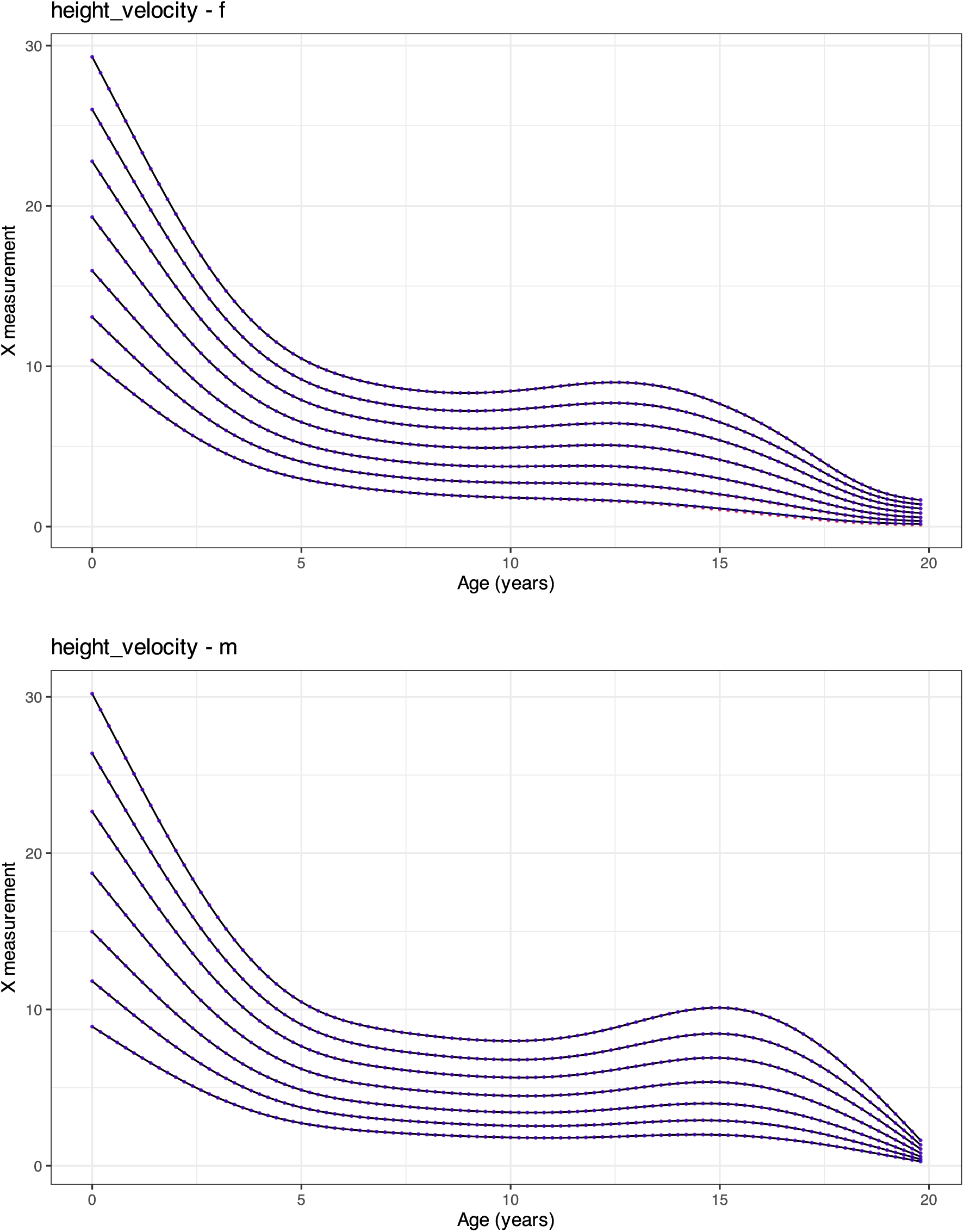
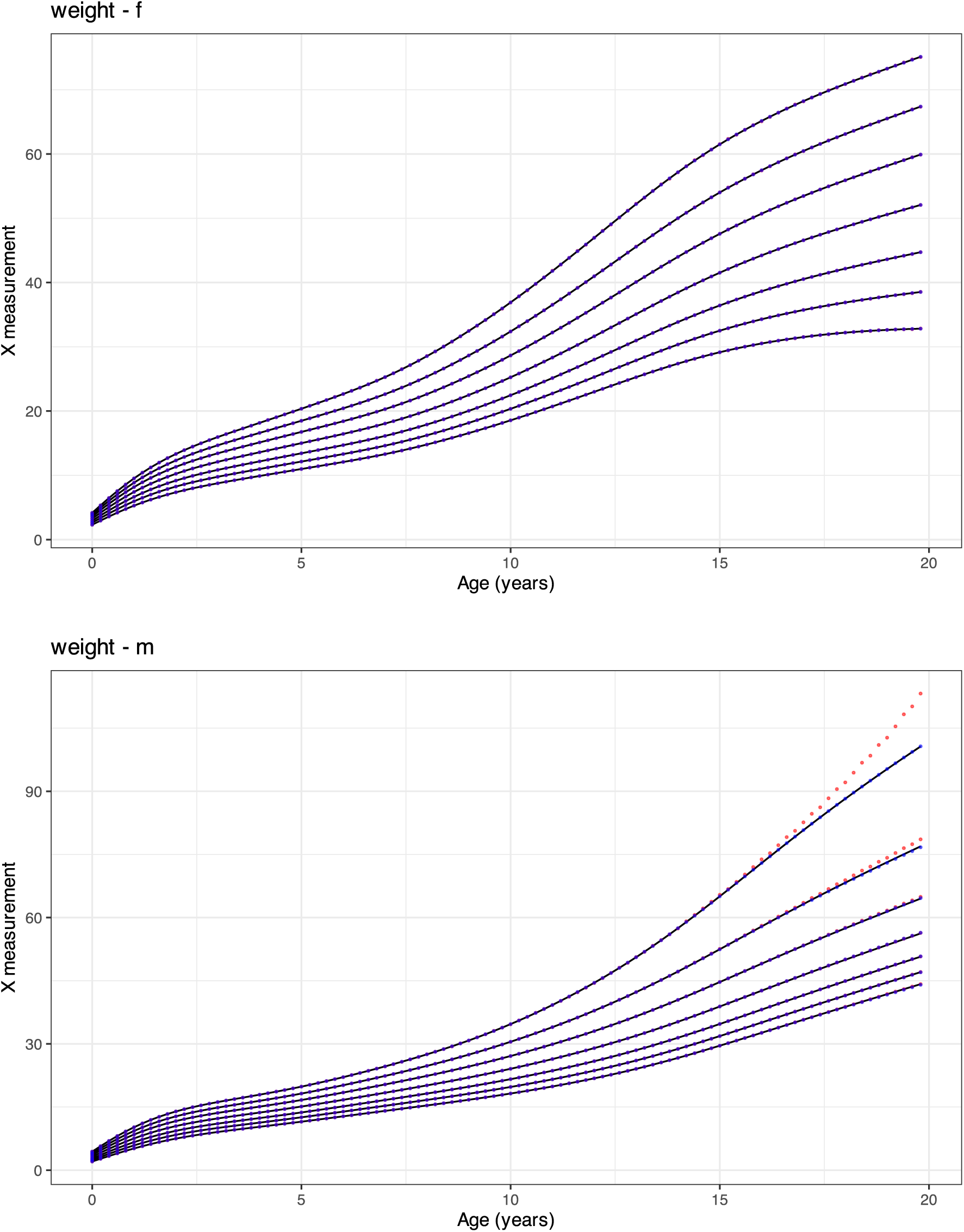

### Re-estimated LMS parameters

Tables of recalculated L, M, and S growth chart parameters for females and males with Noonan Syndrome (NS) for height velocity, weight, height, and body mass index (BMI), derived from published estimates at the 3rd, 10th, 25th, 50th, 75th, 90th, 97th centiles.

**Table.**
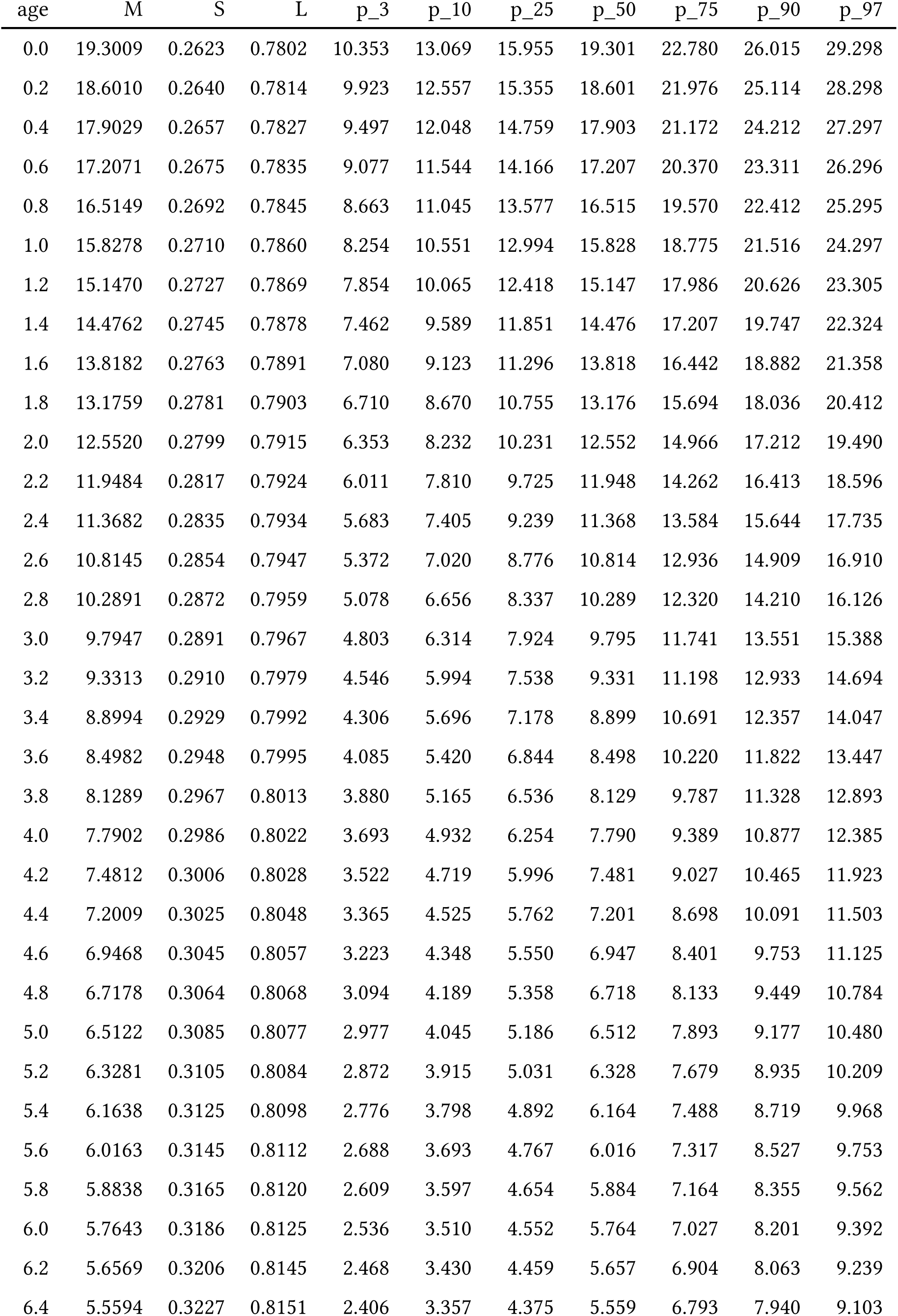
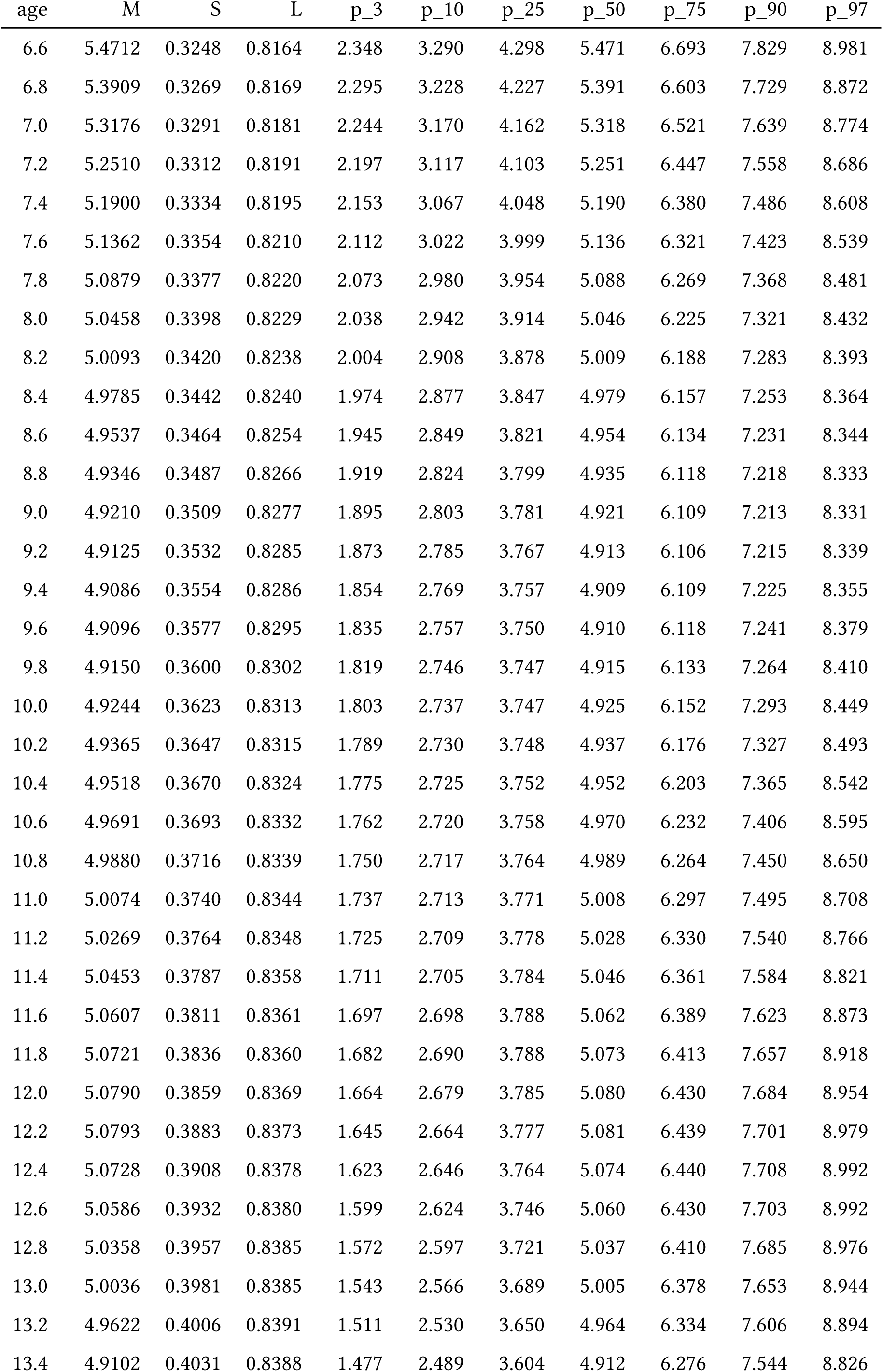
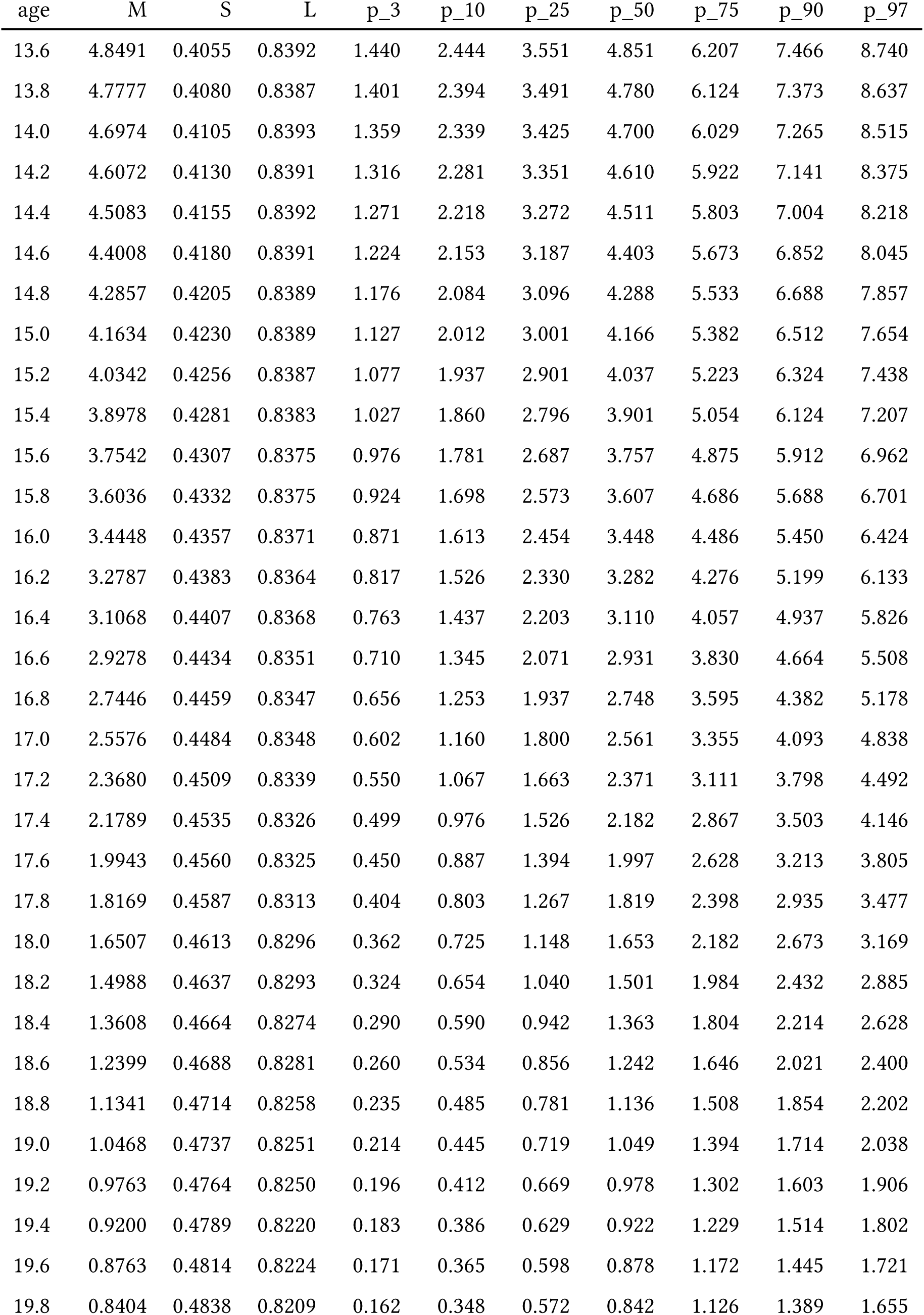
Height velocity (cm/year) for females with NS: Estimated M, S, L parameters from centile values by age (3rd, 10th, 25th, 50th, 75th, 90th, 97th)

| age | M | S | L | p_3 | p_10 | p_25 | p_50 | p_75 | p_90 | p_97 |
| --- | --- | --- | --- | --- | --- | --- | --- | --- | --- | --- |
| 0.0 | 19.3009 | 0.2623 | 0.7802 | 10.353 | 13.069 | 15.955 | 19.301 | 22.780 | 26.015 | 29.298 |
| 0.2 | 18.6010 | 0.2640 | 0.7814 | 9.923 | 12.557 | 15.355 | 18.601 | 21.976 | 25.114 | 28.298 |
| 0.4 | 17.9029 | 0.2657 | 0.7827 | 9.497 | 12.048 | 14.759 | 17.903 | 21.172 | 24.212 | 27.297 |
| 0.6 | 17.2071 | 0.2675 | 0.7835 | 9.077 | 11.544 | 14.166 | 17.207 | 20.370 | 23.311 | 26.296 |
| 0.8 | 16.5149 | 0.2692 | 0.7845 | 8.663 | 11.045 | 13.577 | 16.515 | 19.570 | 22.412 | 25.295 |
| 1.0 | 15.8278 | 0.2710 | 0.7860 | 8.254 | 10.551 | 12.994 | 15.828 | 18.775 | 21.516 | 24.297 |
| 1.2 | 15.1470 | 0.2727 | 0.7869 | 7.854 | 10.065 | 12.418 | 15.147 | 17.986 | 20.626 | 23.305 |
| 1.4 | 14.4762 | 0.2745 | 0.7878 | 7.462 | 9.589 | 11.851 | 14.476 | 17.207 | 19.747 | 22.324 |
| 1.6 | 13.8182 | 0.2763 | 0.7891 | 7.080 | 9.123 | 11.296 | 13.818 | 16.442 | 18.882 | 21.358 |
| 1.8 | 13.1759 | 0.2781 | 0.7903 | 6.710 | 8.670 | 10.755 | 13.176 | 15.694 | 18.036 | 20.412 |
| 2.0 | 12.5520 | 0.2799 | 0.7915 | 6.353 | 8.232 | 10.231 | 12.552 | 14.966 | 17.212 | 19.490 |
| 2.2 | 11.9484 | 0.2817 | 0.7924 | 6.011 | 7.810 | 9.725 | 11.948 | 14.262 | 16.413 | 18.596 |
| 2.4 | 11.3682 | 0.2835 | 0.7934 | 5.683 | 7.405 | 9.239 | 11.368 | 13.584 | 15.644 | 17.735 |
| 2.6 | 10.8145 | 0.2854 | 0.7947 | 5.372 | 7.020 | 8.776 | 10.814 | 12.936 | 14.909 | 16.910 |
| 2.8 | 10.2891 | 0.2872 | 0.7959 | 5.078 | 6.656 | 8.337 | 10.289 | 12.320 | 14.210 | 16.126 |
| 3.0 | 9.7947 | 0.2891 | 0.7967 | 4.803 | 6.314 | 7.924 | 9.795 | 11.741 | 13.551 | 15.388 |
| 3.2 | 9.3313 | 0.2910 | 0.7979 | 4.546 | 5.994 | 7.538 | 9.331 | 11.198 | 12.933 | 14.694 |
| 3.4 | 8.8994 | 0.2929 | 0.7992 | 4.306 | 5.696 | 7.178 | 8.899 | 10.691 | 12.357 | 14.047 |
| 3.6 | 8.4982 | 0.2948 | 0.7995 | 4.085 | 5.420 | 6.844 | 8.498 | 10.220 | 11.822 | 13.447 |
| 3.8 | 8.1289 | 0.2967 | 0.8013 | 3.880 | 5.165 | 6.536 | 8.129 | 9.787 | 11.328 | 12.893 |
| 4.0 | 7.7902 | 0.2986 | 0.8022 | 3.693 | 4.932 | 6.254 | 7.790 | 9.389 | 10.877 | 12.385 |
| 4.2 | 7.4812 | 0.3006 | 0.8028 | 3.522 | 4.719 | 5.996 | 7.481 | 9.027 | 10.465 | 11.923 |
| 4.4 | 7.2009 | 0.3025 | 0.8048 | 3.365 | 4.525 | 5.762 | 7.201 | 8.698 | 10.091 | 11.503 |
| 4.6 | 6.9468 | 0.3045 | 0.8057 | 3.223 | 4.348 | 5.550 | 6.947 | 8.401 | 9.753 | 11.125 |
| 4.8 | 6.7178 | 0.3064 | 0.8068 | 3.094 | 4.189 | 5.358 | 6.718 | 8.133 | 9.449 | 10.784 |
| 5.0 | 6.5122 | 0.3085 | 0.8077 | 2.977 | 4.045 | 5.186 | 6.512 | 7.893 | 9.177 | 10.480 |
| 5.2 | 6.3281 | 0.3105 | 0.8084 | 2.872 | 3.915 | 5.031 | 6.328 | 7.679 | 8.935 | 10.209 |
| 5.4 | 6.1638 | 0.3125 | 0.8098 | 2.776 | 3.798 | 4.892 | 6.164 | 7.488 | 8.719 | 9.968 |
| 5.6 | 6.0163 | 0.3145 | 0.8112 | 2.688 | 3.693 | 4.767 | 6.016 | 7.317 | 8.527 | 9.753 |
| 5.8 | 5.8838 | 0.3165 | 0.8120 | 2.609 | 3.597 | 4.654 | 5.884 | 7.164 | 8.355 | 9.562 |
| 6.0 | 5.7643 | 0.3186 | 0.8125 | 2.536 | 3.510 | 4.552 | 5.764 | 7.027 | 8.201 | 9.392 |
| 6.2 | 5.6569 | 0.3206 | 0.8145 | 2.468 | 3.430 | 4.459 | 5.657 | 6.904 | 8.063 | 9.239 |
| 6.4 | 5.5594 | 0.3227 | 0.8151 | 2.406 | 3.357 | 4.375 | 5.559 | 6.793 | 7.940 | 9.103 |
| 6.6 | 5.4712 | 0.3248 | 0.8164 | 2.348 | 3.290 | 4.298 | 5.471 | 6.693 | 7.829 | 8.981 |
| 6.8 | 5.3909 | 0.3269 | 0.8169 | 2.295 | 3.228 | 4.227 | 5.391 | 6.603 | 7.729 | 8.872 |
| 7.0 | 5.3176 | 0.3291 | 0.8181 | 2.244 | 3.170 | 4.162 | 5.318 | 6.521 | 7.639 | 8.774 |
| 7.2 | 5.2510 | 0.3312 | 0.8191 | 2.197 | 3.117 | 4.103 | 5.251 | 6.447 | 7.558 | 8.686 |
| 7.4 | 5.1900 | 0.3334 | 0.8195 | 2.153 | 3.067 | 4.048 | 5.190 | 6.380 | 7.486 | 8.608 |
| 7.6 | 5.1362 | 0.3354 | 0.8210 | 2.112 | 3.022 | 3.999 | 5.136 | 6.321 | 7.423 | 8.539 |
| 7.8 | 5.0879 | 0.3377 | 0.8220 | 2.073 | 2.980 | 3.954 | 5.088 | 6.269 | 7.368 | 8.481 |
| 8.0 | 5.0458 | 0.3398 | 0.8229 | 2.038 | 2.942 | 3.914 | 5.046 | 6.225 | 7.321 | 8.432 |
| 8.2 | 5.0093 | 0.3420 | 0.8238 | 2.004 | 2.908 | 3.878 | 5.009 | 6.188 | 7.283 | 8.393 |
| 8.4 | 4.9785 | 0.3442 | 0.8240 | 1.974 | 2.877 | 3.847 | 4.979 | 6.157 | 7.253 | 8.364 |
| 8.6 | 4.9537 | 0.3464 | 0.8254 | 1.945 | 2.849 | 3.821 | 4.954 | 6.134 | 7.231 | 8.344 |
| 8.8 | 4.9346 | 0.3487 | 0.8266 | 1.919 | 2.824 | 3.799 | 4.935 | 6.118 | 7.218 | 8.333 |
| 9.0 | 4.9210 | 0.3509 | 0.8277 | 1.895 | 2.803 | 3.781 | 4.921 | 6.109 | 7.213 | 8.331 |
| 9.2 | 4.9125 | 0.3532 | 0.8285 | 1.873 | 2.785 | 3.767 | 4.913 | 6.106 | 7.215 | 8.339 |
| 9.4 | 4.9086 | 0.3554 | 0.8286 | 1.854 | 2.769 | 3.757 | 4.909 | 6.109 | 7.225 | 8.355 |
| 9.6 | 4.9096 | 0.3577 | 0.8295 | 1.835 | 2.757 | 3.750 | 4.910 | 6.118 | 7.241 | 8.379 |
| 9.8 | 4.9150 | 0.3600 | 0.8302 | 1.819 | 2.746 | 3.747 | 4.915 | 6.133 | 7.264 | 8.410 |
| 10.0 | 4.9244 | 0.3623 | 0.8313 | 1.803 | 2.737 | 3.747 | 4.925 | 6.152 | 7.293 | 8.449 |
| 10.2 | 4.9365 | 0.3647 | 0.8315 | 1.789 | 2.730 | 3.748 | 4.937 | 6.176 | 7.327 | 8.493 |
| 10.4 | 4.9518 | 0.3670 | 0.8324 | 1.775 | 2.725 | 3.752 | 4.952 | 6.203 | 7.365 | 8.542 |
| 10.6 | 4.9691 | 0.3693 | 0.8332 | 1.762 | 2.720 | 3.758 | 4.970 | 6.232 | 7.406 | 8.595 |
| 10.8 | 4.9880 | 0.3716 | 0.8339 | 1.750 | 2.717 | 3.764 | 4.989 | 6.264 | 7.450 | 8.650 |
| 11.0 | 5.0074 | 0.3740 | 0.8344 | 1.737 | 2.713 | 3.771 | 5.008 | 6.297 | 7.495 | 8.708 |
| 11.2 | 5.0269 | 0.3764 | 0.8348 | 1.725 | 2.709 | 3.778 | 5.028 | 6.330 | 7.540 | 8.766 |
| 11.4 | 5.0453 | 0.3787 | 0.8358 | 1.711 | 2.705 | 3.784 | 5.046 | 6.361 | 7.584 | 8.821 |
| 11.6 | 5.0607 | 0.3811 | 0.8361 | 1.697 | 2.698 | 3.788 | 5.062 | 6.389 | 7.623 | 8.873 |
| 11.8 | 5.0721 | 0.3836 | 0.8360 | 1.682 | 2.690 | 3.788 | 5.073 | 6.413 | 7.657 | 8.918 |
| 12.0 | 5.0790 | 0.3859 | 0.8369 | 1.664 | 2.679 | 3.785 | 5.080 | 6.430 | 7.684 | 8.954 |
| 12.2 | 5.0793 | 0.3883 | 0.8373 | 1.645 | 2.664 | 3.777 | 5.081 | 6.439 | 7.701 | 8.979 |
| 12.4 | 5.0728 | 0.3908 | 0.8378 | 1.623 | 2.646 | 3.764 | 5.074 | 6.440 | 7.708 | 8.992 |
| 12.6 | 5.0586 | 0.3932 | 0.8380 | 1.599 | 2.624 | 3.746 | 5.060 | 6.430 | 7.703 | 8.992 |
| 12.8 | 5.0358 | 0.3957 | 0.8385 | 1.572 | 2.597 | 3.721 | 5.037 | 6.410 | 7.685 | 8.976 |
| 13.0 | 5.0036 | 0.3981 | 0.8385 | 1.543 | 2.566 | 3.689 | 5.005 | 6.378 | 7.653 | 8.944 |
| 13.2 | 4.9622 | 0.4006 | 0.8391 | 1.511 | 2.530 | 3.650 | 4.964 | 6.334 | 7.606 | 8.894 |
| 13.4 | 4.9102 | 0.4031 | 0.8388 | 1.477 | 2.489 | 3.604 | 4.912 | 6.276 | 7.544 | 8.826 |
| 13.6 | 4.8491 | 0.4055 | 0.8392 | 1.440 | 2.444 | 3.551 | 4.851 | 6.207 | 7.466 | 8.740 |
| 13.8 | 4.7777 | 0.4080 | 0.8387 | 1.401 | 2.394 | 3.491 | 4.780 | 6.124 | 7.373 | 8.637 |
| 14.0 | 4.6974 | 0.4105 | 0.8393 | 1.359 | 2.339 | 3.425 | 4.700 | 6.029 | 7.265 | 8.515 |
| 14.2 | 4.6072 | 0.4130 | 0.8391 | 1.316 | 2.281 | 3.351 | 4.610 | 5.922 | 7.141 | 8.375 |
| 14.4 | 4.5083 | 0.4155 | 0.8392 | 1.271 | 2.218 | 3.272 | 4.511 | 5.803 | 7.004 | 8.218 |
| 14.6 | 4.4008 | 0.4180 | 0.8391 | 1.224 | 2.153 | 3.187 | 4.403 | 5.673 | 6.852 | 8.045 |
| 14.8 | 4.2857 | 0.4205 | 0.8389 | 1.176 | 2.084 | 3.096 | 4.288 | 5.533 | 6.688 | 7.857 |
| 15.0 | 4.1634 | 0.4230 | 0.8389 | 1.127 | 2.012 | 3.001 | 4.166 | 5.382 | 6.512 | 7.654 |
| 15.2 | 4.0342 | 0.4256 | 0.8387 | 1.077 | 1.937 | 2.901 | 4.037 | 5.223 | 6.324 | 7.438 |
| 15.4 | 3.8978 | 0.4281 | 0.8383 | 1.027 | 1.860 | 2.796 | 3.901 | 5.054 | 6.124 | 7.207 |
| 15.6 | 3.7542 | 0.4307 | 0.8375 | 0.976 | 1.781 | 2.687 | 3.757 | 4.875 | 5.912 | 6.962 |
| 15.8 | 3.6036 | 0.4332 | 0.8375 | 0.924 | 1.698 | 2.573 | 3.607 | 4.686 | 5.688 | 6.701 |
| 16.0 | 3.4448 | 0.4357 | 0.8371 | 0.871 | 1.613 | 2.454 | 3.448 | 4.486 | 5.450 | 6.424 |
| 16.2 | 3.2787 | 0.4383 | 0.8364 | 0.817 | 1.526 | 2.330 | 3.282 | 4.276 | 5.199 | 6.133 |
| 16.4 | 3.1068 | 0.4407 | 0.8368 | 0.763 | 1.437 | 2.203 | 3.110 | 4.057 | 4.937 | 5.826 |
| 16.6 | 2.9278 | 0.4434 | 0.8351 | 0.710 | 1.345 | 2.071 | 2.931 | 3.830 | 4.664 | 5.508 |
| 16.8 | 2.7446 | 0.4459 | 0.8347 | 0.656 | 1.253 | 1.937 | 2.748 | 3.595 | 4.382 | 5.178 |
| 17.0 | 2.5576 | 0.4484 | 0.8348 | 0.602 | 1.160 | 1.800 | 2.561 | 3.355 | 4.093 | 4.838 |
| 17.2 | 2.3680 | 0.4509 | 0.8339 | 0.550 | 1.067 | 1.663 | 2.371 | 3.111 | 3.798 | 4.492 |
| 17.4 | 2.1789 | 0.4535 | 0.8326 | 0.499 | 0.976 | 1.526 | 2.182 | 2.867 | 3.503 | 4.146 |
| 17.6 | 1.9943 | 0.4560 | 0.8325 | 0.450 | 0.887 | 1.394 | 1.997 | 2.628 | 3.213 | 3.805 |
| 17.8 | 1.8169 | 0.4587 | 0.8313 | 0.404 | 0.803 | 1.267 | 1.819 | 2.398 | 2.935 | 3.477 |
| 18.0 | 1.6507 | 0.4613 | 0.8296 | 0.362 | 0.725 | 1.148 | 1.653 | 2.182 | 2.673 | 3.169 |
| 18.2 | 1.4988 | 0.4637 | 0.8293 | 0.324 | 0.654 | 1.040 | 1.501 | 1.984 | 2.432 | 2.885 |
| 18.4 | 1.3608 | 0.4664 | 0.8274 | 0.290 | 0.590 | 0.942 | 1.363 | 1.804 | 2.214 | 2.628 |
| 18.6 | 1.2399 | 0.4688 | 0.8281 | 0.260 | 0.534 | 0.856 | 1.242 | 1.646 | 2.021 | 2.400 |
| 18.8 | 1.1341 | 0.4714 | 0.8258 | 0.235 | 0.485 | 0.781 | 1.136 | 1.508 | 1.854 | 2.202 |
| 19.0 | 1.0468 | 0.4737 | 0.8251 | 0.214 | 0.445 | 0.719 | 1.049 | 1.394 | 1.714 | 2.038 |
| 19.2 | 0.9763 | 0.4764 | 0.8250 | 0.196 | 0.412 | 0.669 | 0.978 | 1.302 | 1.603 | 1.906 |
| 19.4 | 0.9200 | 0.4789 | 0.8220 | 0.183 | 0.386 | 0.629 | 0.922 | 1.229 | 1.514 | 1.802 |
| 19.6 | 0.8763 | 0.4814 | 0.8224 | 0.171 | 0.365 | 0.598 | 0.878 | 1.172 | 1.445 | 1.721 |
| 19.8 | 0.8404 | 0.4838 | 0.8209 | 0.162 | 0.348 | 0.572 | 0.842 | 1.126 | 1.389 | 1.655 |

**Table.** Height velocity (cm/year) for males with NS: Estimated M, S, L parameters from centile values by age (3rd, 10th, 25th, 50th, 75th, 90th, 97th)

| age | M | S | L | p_3 | p_10 | p_25 | p_50 | p_75 | p_90 | p_97 |
| --- | --- | --- | --- | --- | --- | --- | --- | --- | --- | --- |
| 0.0 | 18.7102 | 0.3047 | 0.7302 | 8.906 | 11.816 | 14.975 | 18.710 | 22.659 | 26.380 | 30.198 |
| 0.2 | 18.0408 | 0.3059 | 0.7279 | 8.562 | 11.372 | 14.427 | 18.041 | 21.864 | 25.469 | 29.170 |
| 0.4 | 17.3741 | 0.3070 | 0.7257 | 8.220 | 10.932 | 13.882 | 17.374 | 21.071 | 24.559 | 28.142 |
| 0.6 | 16.7095 | 0.3082 | 0.7233 | 7.882 | 10.494 | 13.339 | 16.710 | 20.280 | 23.651 | 27.115 |
| 0.8 | 16.0490 | 0.3094 | 0.7211 | 7.547 | 10.061 | 12.800 | 16.049 | 19.493 | 22.746 | 26.090 |
| 1.0 | 15.3931 | 0.3106 | 0.7186 | 7.217 | 9.632 | 12.267 | 15.393 | 18.710 | 21.845 | 25.070 |
| 1.2 | 14.7446 | 0.3118 | 0.7166 | 6.891 | 9.209 | 11.739 | 14.745 | 17.935 | 20.952 | 24.057 |
| 1.4 | 14.1050 | 0.3130 | 0.7142 | 6.572 | 8.793 | 11.220 | 14.105 | 17.170 | 20.070 | 23.056 |
| 1.6 | 13.4771 | 0.3142 | 0.7119 | 6.260 | 8.386 | 10.711 | 13.477 | 16.418 | 19.202 | 22.070 |
| 1.8 | 12.8628 | 0.3154 | 0.7095 | 5.956 | 7.988 | 10.214 | 12.863 | 15.681 | 18.352 | 21.104 |
| 2.0 | 12.2643 | 0.3166 | 0.7071 | 5.662 | 7.602 | 9.730 | 12.264 | 14.963 | 17.522 | 20.159 |
| 2.2 | 11.6841 | 0.3178 | 0.7049 | 5.377 | 7.229 | 9.261 | 11.684 | 14.266 | 16.715 | 19.241 |
| 2.4 | 11.1235 | 0.3191 | 0.7024 | 5.103 | 6.869 | 8.809 | 11.123 | 13.592 | 15.935 | 18.353 |
| 2.6 | 10.5846 | 0.3203 | 0.7005 | 4.840 | 6.524 | 8.374 | 10.585 | 12.943 | 15.183 | 17.496 |
| 2.8 | 10.0689 | 0.3215 | 0.6978 | 4.590 | 6.194 | 7.959 | 10.069 | 12.322 | 14.464 | 16.676 |
| 3.0 | 9.5791 | 0.3228 | 0.6956 | 4.353 | 5.881 | 7.565 | 9.579 | 11.732 | 13.779 | 15.895 |
| 3.2 | 9.1152 | 0.3240 | 0.6932 | 4.129 | 5.586 | 7.192 | 9.115 | 11.172 | 13.130 | 15.154 |
| 3.4 | 8.6783 | 0.3252 | 0.6908 | 3.919 | 5.308 | 6.841 | 8.678 | 10.645 | 12.518 | 14.455 |
| 3.6 | 8.2690 | 0.3265 | 0.6886 | 3.722 | 5.048 | 6.512 | 8.269 | 10.151 | 11.944 | 13.800 |
| 3.8 | 7.8881 | 0.3278 | 0.6862 | 3.539 | 4.806 | 6.206 | 7.888 | 9.691 | 11.410 | 13.190 |
| 4.0 | 7.5362 | 0.3291 | 0.6841 | 3.370 | 4.582 | 5.924 | 7.536 | 9.266 | 10.916 | 12.626 |
| 4.2 | 7.2126 | 0.3303 | 0.6817 | 3.215 | 4.377 | 5.664 | 7.213 | 8.875 | 10.462 | 12.108 |
| 4.4 | 6.9173 | 0.3316 | 0.6790 | 3.074 | 4.190 | 5.427 | 6.917 | 8.519 | 10.048 | 11.635 |
| 4.6 | 6.6497 | 0.3328 | 0.6772 | 2.945 | 4.020 | 5.212 | 6.650 | 8.195 | 9.673 | 11.206 |
| 4.8 | 6.4090 | 0.3342 | 0.6750 | 2.829 | 3.866 | 5.019 | 6.409 | 7.905 | 9.336 | 10.822 |
| 5.0 | 6.1939 | 0.3354 | 0.6722 | 2.726 | 3.729 | 4.846 | 6.194 | 7.646 | 9.036 | 10.480 |
| 5.2 | 6.0040 | 0.3367 | 0.6704 | 2.633 | 3.608 | 4.693 | 6.004 | 7.417 | 8.771 | 10.178 |
| 5.4 | 5.8362 | 0.3380 | 0.6682 | 2.551 | 3.500 | 4.557 | 5.836 | 7.216 | 8.538 | 9.913 |
| 5.6 | 5.6881 | 0.3394 | 0.6657 | 2.478 | 3.404 | 4.437 | 5.688 | 7.039 | 8.334 | 9.682 |
| 5.8 | 5.5584 | 0.3406 | 0.6632 | 2.414 | 3.320 | 4.332 | 5.558 | 6.884 | 8.156 | 9.480 |
| 6.0 | 5.4445 | 0.3419 | 0.6607 | 2.357 | 3.245 | 4.239 | 5.445 | 6.748 | 8.000 | 9.305 |
| 6.2 | 5.3449 | 0.3432 | 0.6587 | 2.306 | 3.179 | 4.158 | 5.345 | 6.630 | 7.865 | 9.153 |
| 6.4 | 5.2568 | 0.3446 | 0.6563 | 2.260 | 3.121 | 4.085 | 5.257 | 6.526 | 7.747 | 9.021 |
| 6.6 | 5.1781 | 0.3459 | 0.6537 | 2.219 | 3.068 | 4.020 | 5.178 | 6.434 | 7.642 | 8.904 |
| 6.8 | 5.1072 | 0.3472 | 0.6517 | 2.181 | 3.020 | 3.961 | 5.107 | 6.351 | 7.549 | 8.800 |
| 7.0 | 5.0420 | 0.3486 | 0.6494 | 2.146 | 2.975 | 3.907 | 5.042 | 6.275 | 7.463 | 8.706 |
| 7.2 | 4.9813 | 0.3499 | 0.6471 | 2.113 | 2.933 | 3.856 | 4.981 | 6.205 | 7.385 | 8.619 |
| 7.4 | 4.9246 | 0.3513 | 0.6446 | 2.082 | 2.894 | 3.808 | 4.925 | 6.139 | 7.312 | 8.539 |
| 7.6 | 4.8716 | 0.3526 | 0.6424 | 2.053 | 2.857 | 3.763 | 4.872 | 6.078 | 7.244 | 8.464 |
| 7.8 | 4.8216 | 0.3540 | 0.6398 | 2.025 | 2.821 | 3.721 | 4.822 | 6.021 | 7.181 | 8.396 |
| 8.0 | 4.7748 | 0.3553 | 0.6382 | 1.998 | 2.788 | 3.681 | 4.775 | 5.968 | 7.121 | 8.331 |
| 8.2 | 4.7300 | 0.3567 | 0.6353 | 1.973 | 2.756 | 3.643 | 4.730 | 5.917 | 7.066 | 8.271 |
| 8.4 | 4.6881 | 0.3581 | 0.6326 | 1.949 | 2.726 | 3.607 | 4.688 | 5.870 | 7.014 | 8.216 |
| 8.6 | 4.6500 | 0.3594 | 0.6313 | 1.926 | 2.698 | 3.574 | 4.650 | 5.827 | 6.967 | 8.165 |
| 8.8 | 4.6141 | 0.3609 | 0.6280 | 1.905 | 2.672 | 3.543 | 4.614 | 5.787 | 6.925 | 8.121 |
| 9.0 | 4.5823 | 0.3623 | 0.6260 | 1.885 | 2.648 | 3.515 | 4.582 | 5.752 | 6.888 | 8.082 |
| 9.2 | 4.5539 | 0.3636 | 0.6239 | 1.867 | 2.626 | 3.490 | 4.554 | 5.721 | 6.855 | 8.049 |
| 9.4 | 4.5290 | 0.3651 | 0.6215 | 1.850 | 2.606 | 3.467 | 4.529 | 5.695 | 6.829 | 8.023 |
| 9.6 | 4.5082 | 0.3665 | 0.6192 | 1.835 | 2.588 | 3.448 | 4.508 | 5.674 | 6.808 | 8.004 |
| 9.8 | 4.4917 | 0.3679 | 0.6174 | 1.822 | 2.573 | 3.431 | 4.492 | 5.658 | 6.794 | 7.991 |
| 10.0 | 4.4790 | 0.3693 | 0.6142 | 1.811 | 2.561 | 3.418 | 4.479 | 5.647 | 6.786 | 7.987 |
| 10.2 | 4.4719 | 0.3707 | 0.6129 | 1.801 | 2.551 | 3.409 | 4.472 | 5.643 | 6.785 | 7.991 |
| 10.4 | 4.4690 | 0.3721 | 0.6098 | 1.794 | 2.544 | 3.403 | 4.469 | 5.645 | 6.792 | 8.005 |
| 10.6 | 4.4722 | 0.3736 | 0.6077 | 1.789 | 2.540 | 3.402 | 4.472 | 5.654 | 6.808 | 8.028 |
| 10.8 | 4.4819 | 0.3750 | 0.6050 | 1.787 | 2.540 | 3.406 | 4.482 | 5.671 | 6.834 | 8.064 |
| 11.0 | 4.4981 | 0.3764 | 0.6031 | 1.787 | 2.543 | 3.415 | 4.498 | 5.697 | 6.869 | 8.111 |
| 11.2 | 4.5210 | 0.3779 | 0.6008 | 1.789 | 2.551 | 3.428 | 4.521 | 5.731 | 6.916 | 8.171 |
| 11.4 | 4.5508 | 0.3793 | 0.5986 | 1.795 | 2.562 | 3.447 | 4.551 | 5.774 | 6.973 | 8.243 |
| 11.6 | 4.5869 | 0.3808 | 0.5963 | 1.802 | 2.577 | 3.471 | 4.587 | 5.825 | 7.039 | 8.328 |
| 11.8 | 4.6290 | 0.3822 | 0.5934 | 1.813 | 2.595 | 3.499 | 4.629 | 5.884 | 7.116 | 8.424 |
| 12.0 | 4.6772 | 0.3838 | 0.5911 | 1.825 | 2.616 | 3.532 | 4.677 | 5.951 | 7.202 | 8.532 |
| 12.2 | 4.7309 | 0.3853 | 0.5890 | 1.839 | 2.640 | 3.568 | 4.731 | 6.025 | 7.297 | 8.650 |
| 12.4 | 4.7891 | 0.3867 | 0.5870 | 1.855 | 2.666 | 3.608 | 4.789 | 6.105 | 7.399 | 8.776 |
| 12.6 | 4.8502 | 0.3882 | 0.5846 | 1.872 | 2.694 | 3.650 | 4.850 | 6.189 | 7.506 | 8.909 |
| 12.8 | 4.9140 | 0.3897 | 0.5819 | 1.890 | 2.724 | 3.694 | 4.914 | 6.276 | 7.618 | 9.048 |
| 13.0 | 4.9791 | 0.3912 | 0.5799 | 1.908 | 2.753 | 3.739 | 4.979 | 6.365 | 7.732 | 9.189 |
| 13.2 | 5.0438 | 0.3927 | 0.5772 | 1.926 | 2.783 | 3.784 | 5.044 | 6.453 | 7.846 | 9.331 |
| 13.4 | 5.1068 | 0.3942 | 0.5754 | 1.942 | 2.811 | 3.827 | 5.107 | 6.540 | 7.957 | 9.470 |
| 13.6 | 5.1663 | 0.3957 | 0.5729 | 1.958 | 2.838 | 3.867 | 5.166 | 6.623 | 8.064 | 9.603 |
| 13.8 | 5.2208 | 0.3973 | 0.5710 | 1.971 | 2.861 | 3.903 | 5.221 | 6.699 | 8.163 | 9.727 |
| 14.0 | 5.2678 | 0.3988 | 0.5683 | 1.982 | 2.880 | 3.934 | 5.268 | 6.766 | 8.251 | 9.839 |
| 14.2 | 5.3070 | 0.4003 | 0.5662 | 1.989 | 2.895 | 3.959 | 5.307 | 6.823 | 8.326 | 9.936 |
| 14.4 | 5.3358 | 0.4019 | 0.5636 | 1.993 | 2.904 | 3.976 | 5.336 | 6.867 | 8.386 | 10.015 |
| 14.6 | 5.3530 | 0.4034 | 0.5612 | 1.992 | 2.907 | 3.984 | 5.353 | 6.896 | 8.428 | 10.072 |
| 14.8 | 5.3575 | 0.4050 | 0.5592 | 1.986 | 2.902 | 3.983 | 5.358 | 6.908 | 8.450 | 10.105 |
| 15.0 | 5.3472 | 0.4065 | 0.5566 | 1.975 | 2.890 | 3.971 | 5.347 | 6.902 | 8.449 | 10.111 |
| 15.2 | 5.3220 | 0.4081 | 0.5546 | 1.958 | 2.869 | 3.948 | 5.322 | 6.876 | 8.424 | 10.088 |
| 15.4 | 5.2801 | 0.4097 | 0.5523 | 1.935 | 2.840 | 3.912 | 5.280 | 6.829 | 8.373 | 10.034 |
| 15.6 | 5.2224 | 0.4113 | 0.5498 | 1.907 | 2.803 | 3.865 | 5.222 | 6.761 | 8.296 | 9.949 |
| 15.8 | 5.1471 | 0.4129 | 0.5471 | 1.873 | 2.756 | 3.805 | 5.147 | 6.670 | 8.192 | 9.831 |
| 16.0 | 5.0554 | 0.4145 | 0.5453 | 1.832 | 2.700 | 3.733 | 5.055 | 6.558 | 8.060 | 9.680 |
| 16.2 | 4.9461 | 0.4160 | 0.5429 | 1.786 | 2.636 | 3.648 | 4.946 | 6.422 | 7.900 | 9.494 |
| 16.4 | 4.8199 | 0.4176 | 0.5406 | 1.734 | 2.562 | 3.551 | 4.820 | 6.265 | 7.712 | 9.276 |
| 16.6 | 4.6769 | 0.4192 | 0.5381 | 1.676 | 2.481 | 3.441 | 4.677 | 6.085 | 7.498 | 9.024 |
| 16.8 | 4.5183 | 0.4208 | 0.5360 | 1.613 | 2.391 | 3.321 | 4.518 | 5.885 | 7.256 | 8.740 |
| 17.0 | 4.3439 | 0.4224 | 0.5337 | 1.545 | 2.293 | 3.189 | 4.344 | 5.663 | 6.989 | 8.424 |
| 17.2 | 4.1537 | 0.4241 | 0.5309 | 1.472 | 2.187 | 3.046 | 4.154 | 5.421 | 6.696 | 8.078 |
| 17.4 | 3.9502 | 0.4258 | 0.5289 | 1.394 | 2.075 | 2.893 | 3.950 | 5.161 | 6.380 | 7.702 |
| 17.6 | 3.7336 | 0.4274 | 0.5269 | 1.313 | 1.956 | 2.731 | 3.734 | 4.883 | 6.041 | 7.298 |
| 17.8 | 3.5047 | 0.4290 | 0.5247 | 1.227 | 1.832 | 2.561 | 3.505 | 4.588 | 5.681 | 6.869 |
| 18.0 | 3.2649 | 0.4307 | 0.5221 | 1.139 | 1.703 | 2.382 | 3.265 | 4.279 | 5.303 | 6.416 |
| 18.2 | 3.0147 | 0.4324 | 0.5198 | 1.048 | 1.568 | 2.197 | 3.015 | 3.955 | 4.906 | 5.940 |
| 18.4 | 2.7564 | 0.4340 | 0.5180 | 0.954 | 1.430 | 2.007 | 2.756 | 3.620 | 4.494 | 5.445 |
| 18.6 | 2.4902 | 0.4357 | 0.5151 | 0.859 | 1.289 | 1.811 | 2.490 | 3.274 | 4.068 | 4.933 |
| 18.8 | 2.2187 | 0.4374 | 0.5130 | 0.762 | 1.146 | 1.611 | 2.219 | 2.920 | 3.631 | 4.407 |
| 19.0 | 1.9423 | 0.4389 | 0.5103 | 0.665 | 1.001 | 1.409 | 1.942 | 2.559 | 3.185 | 3.868 |
| 19.2 | 1.6622 | 0.4408 | 0.5078 | 0.567 | 0.854 | 1.204 | 1.662 | 2.193 | 2.731 | 3.320 |
| 19.4 | 1.3798 | 0.4424 | 0.5060 | 0.469 | 0.707 | 0.998 | 1.380 | 1.822 | 2.272 | 2.763 |
| 19.6 | 1.0955 | 0.4441 | 0.5054 | 0.370 | 0.560 | 0.792 | 1.095 | 1.448 | 1.807 | 2.199 |
| 19.8 | 0.8091 | 0.4458 | 0.4993 | 0.273 | 0.413 | 0.584 | 0.809 | 1.071 | 1.337 | 1.630 |

**Table.** Weight (kg) for females with NS: Estimated M, S, L parameters from centile values by age (3rd, 10th, 25th, 50th, 75th, 90th, 97th)

| age | M | S | L | p_3 | p_10 | p_25 | p_50 | p_75 | p_90 | p_97 |
| --- | --- | --- | --- | --- | --- | --- | --- | --- | --- | --- |
| 0.0 | 3.2710 | 0.1494 | 1.2962 | 2.306 | 2.625 | 2.936 | 3.271 | 3.596 | 3.881 | 4.157 |
| 0.2 | 4.1820 | 0.1500 | 1.2402 | 2.955 | 3.357 | 3.754 | 4.182 | 4.600 | 4.969 | 5.327 |
| 0.4 | 5.0616 | 0.1506 | 1.1881 | 3.584 | 4.065 | 4.542 | 5.062 | 5.571 | 6.022 | 6.461 |
| 0.6 | 5.8992 | 0.1512 | 1.1329 | 4.186 | 4.740 | 5.294 | 5.899 | 6.497 | 7.028 | 7.548 |
| 0.8 | 6.6860 | 0.1517 | 1.0828 | 4.753 | 5.375 | 5.999 | 6.686 | 7.367 | 7.976 | 8.573 |
| 1.0 | 7.4128 | 0.1523 | 1.0284 | 5.280 | 5.962 | 6.650 | 7.413 | 8.173 | 8.856 | 9.528 |
| 1.2 | 8.0810 | 0.1529 | 0.9747 | 5.767 | 6.502 | 7.249 | 8.081 | 8.915 | 9.668 | 10.412 |
| 1.4 | 8.6943 | 0.1534 | 0.9227 | 6.216 | 6.999 | 7.798 | 8.694 | 9.598 | 10.416 | 11.229 |
| 1.6 | 9.2561 | 0.1540 | 0.8711 | 6.629 | 7.454 | 8.301 | 9.256 | 10.224 | 11.105 | 11.984 |
| 1.8 | 9.7692 | 0.1546 | 0.8189 | 7.009 | 7.870 | 8.761 | 9.769 | 10.797 | 11.738 | 12.680 |
| 2.0 | 10.2379 | 0.1551 | 0.7671 | 7.358 | 8.251 | 9.180 | 10.238 | 11.322 | 12.319 | 13.322 |
| 2.2 | 10.6676 | 0.1557 | 0.7171 | 7.679 | 8.601 | 9.564 | 10.668 | 11.804 | 12.855 | 13.916 |
| 2.4 | 11.0629 | 0.1563 | 0.6673 | 7.975 | 8.922 | 9.917 | 11.063 | 12.250 | 13.351 | 14.470 |
| 2.6 | 11.4300 | 0.1569 | 0.6178 | 8.252 | 9.222 | 10.245 | 11.430 | 12.664 | 13.815 | 14.989 |
| 2.8 | 11.7739 | 0.1575 | 0.5683 | 8.513 | 9.502 | 10.552 | 11.774 | 13.053 | 14.253 | 15.481 |
| 3.0 | 12.1010 | 0.1581 | 0.5204 | 8.761 | 9.769 | 10.844 | 12.101 | 13.424 | 14.671 | 15.954 |
| 3.2 | 12.4131 | 0.1587 | 0.4725 | 8.999 | 10.024 | 11.122 | 12.413 | 13.779 | 15.073 | 16.410 |
| 3.4 | 12.7149 | 0.1593 | 0.4256 | 9.229 | 10.270 | 11.391 | 12.715 | 14.123 | 15.464 | 16.856 |
| 3.6 | 13.0089 | 0.1599 | 0.3791 | 9.454 | 10.510 | 11.653 | 13.009 | 14.459 | 15.846 | 17.294 |
| 3.8 | 13.2981 | 0.1604 | 0.3337 | 9.675 | 10.747 | 11.910 | 13.298 | 14.790 | 16.224 | 17.728 |
| 4.0 | 13.5849 | 0.1611 | 0.2879 | 9.895 | 10.981 | 12.165 | 13.585 | 15.119 | 16.601 | 18.163 |
| 4.2 | 13.8711 | 0.1617 | 0.2436 | 10.114 | 11.215 | 12.420 | 13.871 | 15.447 | 16.978 | 18.599 |
| 4.4 | 14.1561 | 0.1623 | 0.2000 | 10.332 | 11.447 | 12.673 | 14.156 | 15.775 | 17.355 | 19.037 |
| 4.6 | 14.4399 | 0.1629 | 0.1559 | 10.550 | 11.679 | 12.925 | 14.440 | 16.102 | 17.732 | 19.477 |
| 4.8 | 14.7231 | 0.1635 | 0.1134 | 10.767 | 11.910 | 13.177 | 14.723 | 16.428 | 18.110 | 19.918 |
| 5.0 | 15.0052 | 0.1641 | 0.0717 | 10.983 | 12.140 | 13.427 | 15.005 | 16.754 | 18.488 | 20.361 |
| 5.2 | 15.2879 | 0.1647 | 0.0318 | 11.198 | 12.370 | 13.678 | 15.288 | 17.081 | 18.867 | 20.808 |
| 5.4 | 15.5713 | 0.1653 | -0.0089 | 11.415 | 12.601 | 13.929 | 15.571 | 17.409 | 19.250 | 21.259 |
| 5.6 | 15.8575 | 0.1659 | -0.0481 | 11.633 | 12.834 | 14.183 | 15.857 | 17.741 | 19.636 | 21.717 |
| 5.8 | 16.1472 | 0.1665 | -0.0861 | 11.854 | 13.069 | 14.439 | 16.147 | 18.077 | 20.029 | 22.182 |
| 6.0 | 16.4413 | 0.1672 | -0.1239 | 12.077 | 13.308 | 14.700 | 16.441 | 18.418 | 20.429 | 22.658 |
| 6.2 | 16.7423 | 0.1678 | -0.1604 | 12.306 | 13.552 | 14.966 | 16.742 | 18.768 | 20.838 | 23.145 |
| 6.4 | 17.0521 | 0.1684 | -0.1961 | 12.540 | 13.803 | 15.240 | 17.052 | 19.128 | 21.260 | 23.649 |
| 6.6 | 17.3739 | 0.1691 | -0.2295 | 12.782 | 14.063 | 15.524 | 17.374 | 19.502 | 21.698 | 24.171 |
| 6.8 | 17.7086 | 0.1697 | -0.2628 | 13.034 | 14.333 | 15.820 | 17.709 | 19.891 | 22.153 | 24.715 |
| 7.0 | 18.0591 | 0.1703 | -0.2955 | 13.297 | 14.616 | 16.130 | 18.059 | 20.298 | 22.631 | 25.285 |
| 7.2 | 18.4269 | 0.1710 | -0.3260 | 13.572 | 14.912 | 16.455 | 18.427 | 20.725 | 23.130 | 25.881 |
| 7.4 | 18.8109 | 0.1716 | -0.3568 | 13.859 | 15.222 | 16.794 | 18.811 | 21.171 | 23.652 | 26.505 |
| 7.6 | 19.2131 | 0.1722 | -0.3848 | 14.158 | 15.545 | 17.149 | 19.213 | 21.638 | 24.198 | 27.156 |
| 7.8 | 19.6320 | 0.1729 | -0.4122 | 14.469 | 15.881 | 17.519 | 19.632 | 22.124 | 24.767 | 27.835 |
| 8.0 | 20.0689 | 0.1735 | -0.4383 | 14.792 | 16.232 | 17.904 | 20.069 | 22.631 | 25.360 | 28.543 |
| 8.2 | 20.5234 | 0.1742 | -0.4627 | 15.128 | 16.595 | 18.305 | 20.523 | 23.159 | 25.976 | 29.278 |
| 8.4 | 20.9940 | 0.1748 | -0.4850 | 15.474 | 16.971 | 18.719 | 20.994 | 23.705 | 26.614 | 30.038 |
| 8.6 | 21.4798 | 0.1755 | -0.5063 | 15.830 | 17.359 | 19.147 | 21.480 | 24.268 | 27.273 | 30.823 |
| 8.8 | 21.9798 | 0.1762 | -0.5256 | 16.195 | 17.757 | 19.587 | 21.980 | 24.849 | 27.950 | 31.631 |
| 9.0 | 22.4926 | 0.1768 | -0.5439 | 16.569 | 18.165 | 20.038 | 22.493 | 25.444 | 28.646 | 32.460 |
| 9.2 | 23.0190 | 0.1775 | -0.5602 | 16.951 | 18.583 | 20.501 | 23.019 | 26.055 | 29.359 | 33.309 |
| 9.4 | 23.5579 | 0.1781 | -0.5744 | 17.340 | 19.010 | 20.974 | 23.558 | 26.681 | 30.089 | 34.178 |
| 9.6 | 24.1099 | 0.1788 | -0.5870 | 17.738 | 19.446 | 21.458 | 24.110 | 27.322 | 30.837 | 35.066 |
| 9.8 | 24.6746 | 0.1795 | -0.5984 | 18.144 | 19.891 | 21.953 | 24.675 | 27.978 | 31.602 | 35.975 |
| 10.0 | 25.2530 | 0.1801 | -0.6075 | 18.557 | 20.346 | 22.460 | 25.253 | 28.650 | 32.385 | 36.903 |
| 10.2 | 25.8445 | 0.1808 | -0.6149 | 18.978 | 20.811 | 22.977 | 25.844 | 29.338 | 33.185 | 37.850 |
| 10.4 | 26.4480 | 0.1815 | -0.6209 | 19.406 | 21.283 | 23.505 | 26.448 | 30.039 | 34.001 | 38.815 |
| 10.6 | 27.0632 | 0.1822 | -0.6249 | 19.840 | 21.764 | 24.042 | 27.063 | 30.754 | 34.832 | 39.795 |
| 10.8 | 27.6898 | 0.1829 | -0.6269 | 20.280 | 22.252 | 24.588 | 27.690 | 31.482 | 35.677 | 40.789 |
| 11.0 | 28.3260 | 0.1835 | -0.6270 | 20.724 | 22.746 | 25.142 | 28.326 | 32.222 | 36.534 | 41.795 |
| 11.2 | 28.9717 | 0.1842 | -0.6257 | 21.173 | 23.246 | 25.704 | 28.972 | 32.972 | 37.404 | 42.813 |
| 11.4 | 29.6268 | 0.1849 | -0.6220 | 21.625 | 23.751 | 26.273 | 29.627 | 33.734 | 38.285 | 43.841 |
| 11.6 | 30.2900 | 0.1856 | -0.6168 | 22.080 | 24.261 | 26.849 | 30.290 | 34.505 | 39.175 | 44.877 |
| 11.8 | 30.9609 | 0.1863 | -0.6096 | 22.537 | 24.775 | 27.430 | 30.961 | 35.285 | 40.075 | 45.920 |
| 12.0 | 31.6387 | 0.1870 | -0.6009 | 22.996 | 25.292 | 28.017 | 31.639 | 36.073 | 40.982 | 46.969 |
| 12.2 | 32.3224 | 0.1877 | -0.5906 | 23.456 | 25.812 | 28.608 | 32.322 | 36.868 | 41.896 | 48.021 |
| 12.4 | 33.0106 | 0.1884 | -0.5783 | 23.915 | 26.333 | 29.201 | 33.011 | 37.668 | 42.814 | 49.074 |
| 12.6 | 33.7011 | 0.1891 | -0.5643 | 24.371 | 26.853 | 29.796 | 33.701 | 38.471 | 43.734 | 50.125 |
| 12.8 | 34.3928 | 0.1898 | -0.5489 | 24.825 | 27.372 | 30.390 | 34.393 | 39.275 | 44.654 | 51.171 |
| 13.0 | 35.0840 | 0.1905 | -0.5312 | 25.273 | 27.887 | 30.983 | 35.084 | 40.079 | 45.570 | 52.208 |
| 13.2 | 35.7723 | 0.1912 | -0.5125 | 25.715 | 28.398 | 31.572 | 35.772 | 40.879 | 46.482 | 53.235 |
| 13.4 | 36.4560 | 0.1919 | -0.4921 | 26.149 | 28.902 | 32.156 | 36.456 | 41.674 | 47.385 | 54.248 |
| 13.6 | 37.1328 | 0.1927 | -0.4700 | 26.573 | 29.397 | 32.732 | 37.133 | 42.461 | 48.277 | 55.243 |
| 13.8 | 37.8004 | 0.1934 | -0.4464 | 26.985 | 29.882 | 33.299 | 37.800 | 43.238 | 49.155 | 56.217 |
| 14.0 | 38.4575 | 0.1941 | -0.4214 | 27.384 | 30.356 | 33.856 | 38.457 | 44.002 | 50.016 | 57.167 |
| 14.2 | 39.1015 | 0.1948 | -0.3952 | 27.769 | 30.816 | 34.400 | 39.101 | 44.751 | 50.859 | 58.092 |
| 14.4 | 39.7317 | 0.1956 | -0.3679 | 28.140 | 31.262 | 34.930 | 39.732 | 45.484 | 51.681 | 58.989 |
| 14.6 | 40.3480 | 0.1963 | -0.3391 | 28.494 | 31.694 | 35.447 | 40.348 | 46.201 | 52.483 | 59.858 |
| 14.8 | 40.9493 | 0.1970 | -0.3091 | 28.832 | 32.111 | 35.949 | 40.949 | 46.901 | 53.263 | 60.698 |
| 15.0 | 41.5341 | 0.1978 | -0.2785 | 29.154 | 32.512 | 36.435 | 41.534 | 47.582 | 54.020 | 61.509 |
| 15.2 | 42.1027 | 0.1985 | -0.2470 | 29.459 | 32.897 | 36.906 | 42.103 | 48.244 | 54.754 | 62.291 |
| 15.4 | 42.6551 | 0.1992 | -0.2149 | 29.747 | 33.267 | 37.362 | 42.655 | 48.887 | 55.466 | 63.044 |
| 15.6 | 43.1929 | 0.2000 | -0.1817 | 30.019 | 33.621 | 37.804 | 43.193 | 49.514 | 56.156 | 63.771 |
| 15.8 | 43.7155 | 0.2007 | -0.1485 | 30.276 | 33.962 | 38.231 | 43.715 | 50.123 | 56.827 | 64.473 |
| 16.0 | 44.2241 | 0.2015 | -0.1146 | 30.517 | 34.288 | 38.645 | 44.224 | 50.716 | 57.478 | 65.152 |
| 16.2 | 44.7195 | 0.2022 | -0.0805 | 30.745 | 34.601 | 39.046 | 44.720 | 51.294 | 58.110 | 65.808 |
| 16.4 | 45.2023 | 0.2030 | -0.0457 | 30.958 | 34.901 | 39.435 | 45.202 | 51.858 | 58.725 | 66.442 |
| 16.6 | 45.6728 | 0.2038 | -0.0109 | 31.158 | 35.189 | 39.812 | 45.673 | 52.407 | 59.324 | 67.057 |
| 16.8 | 46.1313 | 0.2045 | 0.0241 | 31.344 | 35.465 | 40.178 | 46.131 | 52.943 | 59.906 | 67.653 |
| 17.0 | 46.5789 | 0.2053 | 0.0592 | 31.518 | 35.730 | 40.533 | 46.579 | 53.466 | 60.474 | 68.232 |
| 17.2 | 47.0155 | 0.2061 | 0.0944 | 31.679 | 35.983 | 40.877 | 47.016 | 53.977 | 61.027 | 68.794 |
| 17.4 | 47.4426 | 0.2068 | 0.1299 | 31.828 | 36.226 | 41.212 | 47.443 | 54.477 | 61.567 | 69.340 |
| 17.6 | 47.8601 | 0.2076 | 0.1654 | 31.965 | 36.459 | 41.538 | 47.860 | 54.966 | 62.094 | 69.871 |
| 17.8 | 48.2698 | 0.2084 | 0.2011 | 32.091 | 36.683 | 41.856 | 48.270 | 55.446 | 62.610 | 70.390 |
| 18.0 | 48.6706 | 0.2092 | 0.2367 | 32.206 | 36.897 | 42.165 | 48.671 | 55.916 | 63.115 | 70.896 |
| 18.2 | 49.0647 | 0.2099 | 0.2726 | 32.310 | 37.103 | 42.467 | 49.065 | 56.378 | 63.611 | 71.390 |
| 18.4 | 49.4524 | 0.2107 | 0.3084 | 32.404 | 37.302 | 42.763 | 49.452 | 56.833 | 64.098 | 71.875 |
| 18.6 | 49.8349 | 0.2115 | 0.3443 | 32.488 | 37.493 | 43.053 | 49.835 | 57.282 | 64.578 | 72.353 |
| 18.8 | 50.2136 | 0.2123 | 0.3801 | 32.564 | 37.678 | 43.338 | 50.214 | 57.727 | 65.053 | 72.824 |
| 19.0 | 50.5894 | 0.2131 | 0.4161 | 32.631 | 37.858 | 43.621 | 50.589 | 58.168 | 65.524 | 73.290 |
| 19.2 | 50.9632 | 0.2139 | 0.4519 | 32.691 | 38.033 | 43.900 | 50.963 | 58.608 | 65.992 | 73.754 |
| 19.4 | 51.3359 | 0.2147 | 0.4879 | 32.742 | 38.204 | 44.177 | 51.336 | 59.046 | 66.458 | 74.216 |
| 19.6 | 51.7073 | 0.2155 | 0.5238 | 32.786 | 38.370 | 44.452 | 51.707 | 59.483 | 66.923 | 74.675 |
| 19.8 | 52.0780 | 0.2163 | 0.5599 | 32.821 | 38.532 | 44.725 | 52.078 | 59.919 | 67.386 | 75.133 |

**Table.** Weight (kg) for males with NS: Estimated M, S, L parameters from centile values by age (3rd, 10th, 25th, 50th, 75th, 90th, 97th)

| age | M | S | L | p_3 | p_10 | p_25 | p_50 | p_75 | p_90 | p_97 |
| --- | --- | --- | --- | --- | --- | --- | --- | --- | --- | --- |
| 0.0 | 3.1950 | 0.1956 | 0.8327 | 2.059 | 2.412 | 2.778 | 3.195 | 3.621 | 4.012 | 4.404 |
| 0.2 | 4.1481 | 0.1919 | 0.8023 | 2.709 | 3.154 | 3.618 | 4.148 | 4.692 | 5.192 | 5.695 |
| 0.4 | 5.0690 | 0.1883 | 0.7673 | 3.353 | 3.882 | 4.435 | 5.069 | 5.722 | 6.325 | 6.934 |
| 0.6 | 5.9480 | 0.1847 | 0.7352 | 3.982 | 4.586 | 5.219 | 5.948 | 6.701 | 7.399 | 8.105 |
| 0.8 | 6.7739 | 0.1813 | 0.7016 | 4.587 | 5.256 | 5.961 | 6.774 | 7.617 | 8.401 | 9.197 |
| 1.0 | 7.5380 | 0.1780 | 0.6685 | 5.160 | 5.885 | 6.651 | 7.538 | 8.461 | 9.321 | 10.197 |
| 1.2 | 8.2402 | 0.1749 | 0.6352 | 5.698 | 6.471 | 7.289 | 8.240 | 9.233 | 10.161 | 11.108 |
| 1.4 | 8.8821 | 0.1718 | 0.6009 | 6.201 | 7.013 | 7.877 | 8.882 | 9.935 | 10.923 | 11.934 |
| 1.6 | 9.4670 | 0.1689 | 0.5693 | 6.668 | 7.514 | 8.415 | 9.467 | 10.572 | 11.611 | 12.678 |
| 1.8 | 9.9955 | 0.1662 | 0.5346 | 7.100 | 7.972 | 8.904 | 9.996 | 11.145 | 12.230 | 13.346 |
| 2.0 | 10.4720 | 0.1637 | 0.5014 | 7.496 | 8.390 | 9.348 | 10.472 | 11.660 | 12.783 | 13.943 |
| 2.2 | 10.9012 | 0.1613 | 0.4675 | 7.860 | 8.772 | 9.750 | 10.901 | 12.121 | 13.279 | 14.476 |
| 2.4 | 11.2903 | 0.1590 | 0.4347 | 8.195 | 9.121 | 10.116 | 11.290 | 12.538 | 13.725 | 14.956 |
| 2.6 | 11.6459 | 0.1570 | 0.4007 | 8.506 | 9.442 | 10.452 | 11.646 | 12.918 | 14.132 | 15.394 |
| 2.8 | 11.9747 | 0.1551 | 0.3681 | 8.796 | 9.742 | 10.763 | 11.975 | 13.269 | 14.506 | 15.797 |
| 3.0 | 12.2832 | 0.1534 | 0.3360 | 9.069 | 10.023 | 11.056 | 12.283 | 13.598 | 14.858 | 16.176 |
| 3.2 | 12.5759 | 0.1518 | 0.3017 | 9.330 | 10.291 | 11.334 | 12.576 | 13.910 | 15.193 | 16.538 |
| 3.4 | 12.8574 | 0.1504 | 0.2683 | 9.580 | 10.549 | 11.601 | 12.857 | 14.211 | 15.516 | 16.888 |
| 3.6 | 13.1317 | 0.1492 | 0.2342 | 9.824 | 10.799 | 11.860 | 13.132 | 14.505 | 15.832 | 17.232 |
| 3.8 | 13.4037 | 0.1481 | 0.2018 | 10.063 | 11.045 | 12.117 | 13.404 | 14.797 | 16.148 | 17.576 |
| 4.0 | 13.6770 | 0.1472 | 0.1688 | 10.301 | 11.291 | 12.374 | 13.677 | 15.092 | 16.468 | 17.926 |
| 4.2 | 13.9535 | 0.1464 | 0.1343 | 10.540 | 11.539 | 12.633 | 13.953 | 15.392 | 16.794 | 18.285 |
| 4.4 | 14.2342 | 0.1458 | 0.1012 | 10.779 | 11.788 | 12.895 | 14.234 | 15.697 | 17.128 | 18.654 |
| 4.6 | 14.5208 | 0.1453 | 0.0680 | 11.021 | 12.040 | 13.161 | 14.521 | 16.010 | 17.472 | 19.035 |
| 4.8 | 14.8138 | 0.1449 | 0.0349 | 11.265 | 12.296 | 13.432 | 14.814 | 16.332 | 17.826 | 19.430 |
| 5.0 | 15.1142 | 0.1447 | 0.0018 | 11.513 | 12.556 | 13.709 | 15.114 | 16.663 | 18.193 | 19.839 |
| 5.2 | 15.4222 | 0.1445 | -0.0319 | 11.765 | 12.821 | 13.992 | 15.422 | 17.004 | 18.571 | 20.264 |
| 5.4 | 15.7361 | 0.1445 | -0.0657 | 12.019 | 13.090 | 14.279 | 15.736 | 17.353 | 18.960 | 20.703 |
| 5.6 | 16.0560 | 0.1447 | -0.0982 | 12.275 | 13.361 | 14.570 | 16.056 | 17.710 | 19.360 | 21.155 |
| 5.8 | 16.3798 | 0.1449 | -0.1318 | 12.532 | 13.634 | 14.864 | 16.380 | 18.073 | 19.768 | 21.620 |
| 6.0 | 16.7079 | 0.1452 | -0.1655 | 12.791 | 13.909 | 15.161 | 16.708 | 18.442 | 20.185 | 22.096 |
| 6.2 | 17.0392 | 0.1457 | -0.1984 | 13.049 | 14.185 | 15.459 | 17.039 | 18.817 | 20.610 | 22.584 |
| 6.4 | 17.3731 | 0.1462 | -0.2331 | 13.308 | 14.462 | 15.759 | 17.373 | 19.196 | 21.042 | 23.084 |
| 6.6 | 17.7102 | 0.1469 | -0.2653 | 13.566 | 14.739 | 16.061 | 17.710 | 19.580 | 21.482 | 23.594 |
| 6.8 | 18.0491 | 0.1476 | -0.2989 | 13.824 | 15.016 | 16.363 | 18.049 | 19.968 | 21.929 | 24.116 |
| 7.0 | 18.3897 | 0.1484 | -0.3325 | 14.082 | 15.293 | 16.665 | 18.390 | 20.360 | 22.382 | 24.648 |
| 7.2 | 18.7328 | 0.1493 | -0.3651 | 14.338 | 15.570 | 16.969 | 18.733 | 20.757 | 22.842 | 25.193 |
| 7.4 | 19.0781 | 0.1503 | -0.3979 | 14.595 | 15.847 | 17.273 | 19.078 | 21.158 | 23.311 | 25.749 |
| 7.6 | 19.4261 | 0.1513 | -0.4322 | 14.853 | 16.125 | 17.579 | 19.426 | 21.564 | 23.788 | 26.320 |
| 7.8 | 19.7782 | 0.1524 | -0.4651 | 15.111 | 16.405 | 17.888 | 19.778 | 21.976 | 24.274 | 26.905 |
| 8.0 | 20.1337 | 0.1536 | -0.4992 | 15.371 | 16.686 | 18.199 | 20.134 | 22.394 | 24.770 | 27.507 |
| 8.2 | 20.4947 | 0.1549 | -0.5322 | 15.633 | 16.971 | 18.513 | 20.495 | 22.820 | 25.278 | 28.125 |
| 8.4 | 20.8609 | 0.1562 | -0.5656 | 15.898 | 17.258 | 18.832 | 20.861 | 23.254 | 25.797 | 28.762 |
| 8.6 | 21.2340 | 0.1575 | -0.5984 | 16.166 | 17.550 | 19.156 | 21.234 | 23.697 | 26.330 | 29.419 |
| 8.8 | 21.6138 | 0.1589 | -0.6324 | 16.439 | 17.847 | 19.485 | 21.614 | 24.150 | 26.877 | 30.099 |
| 9.0 | 22.0024 | 0.1602 | -0.6659 | 16.718 | 18.150 | 19.822 | 22.002 | 24.614 | 27.440 | 30.801 |
| 9.2 | 22.3997 | 0.1617 | -0.6988 | 17.002 | 18.459 | 20.165 | 22.400 | 25.090 | 28.018 | 31.528 |
| 9.4 | 22.8058 | 0.1631 | -0.7319 | 17.292 | 18.775 | 20.516 | 22.806 | 25.577 | 28.614 | 32.280 |
| 9.6 | 23.2203 | 0.1646 | -0.7655 | 17.589 | 19.097 | 20.874 | 23.220 | 26.076 | 29.225 | 33.057 |
| 9.8 | 23.6444 | 0.1660 | -0.7988 | 17.893 | 19.427 | 21.240 | 23.644 | 26.587 | 29.853 | 33.860 |
| 10.0 | 24.0776 | 0.1674 | -0.8323 | 18.204 | 19.764 | 21.614 | 24.078 | 27.109 | 30.497 | 34.689 |
| 10.2 | 24.5202 | 0.1688 | -0.8655 | 18.522 | 20.109 | 21.997 | 24.520 | 27.643 | 31.158 | 35.544 |
| 10.4 | 24.9729 | 0.1702 | -0.8990 | 18.849 | 20.463 | 22.388 | 24.973 | 28.190 | 31.835 | 36.426 |
| 10.6 | 25.4352 | 0.1715 | -0.9327 | 19.184 | 20.825 | 22.789 | 25.435 | 28.748 | 32.530 | 37.336 |
| 10.8 | 25.9082 | 0.1728 | -0.9659 | 19.528 | 21.196 | 23.199 | 25.908 | 29.319 | 33.242 | 38.273 |
| 11.0 | 26.3921 | 0.1741 | -0.9989 | 19.881 | 21.577 | 23.619 | 26.392 | 29.903 | 33.971 | 39.238 |
| 11.2 | 26.8873 | 0.1753 | -1.0324 | 20.245 | 21.969 | 24.049 | 26.887 | 30.501 | 34.717 | 40.232 |
| 11.4 | 27.3939 | 0.1764 | -1.0658 | 20.619 | 22.371 | 24.491 | 27.394 | 31.111 | 35.481 | 41.255 |
| 11.6 | 27.9131 | 0.1775 | -1.0991 | 21.005 | 22.784 | 24.944 | 27.913 | 31.736 | 36.264 | 42.308 |
| 11.8 | 28.4448 | 0.1785 | -1.1325 | 21.402 | 23.210 | 25.409 | 28.445 | 32.375 | 37.065 | 43.392 |
| 12.0 | 28.9903 | 0.1794 | -1.1656 | 21.811 | 23.648 | 25.888 | 28.990 | 33.029 | 37.886 | 44.507 |
| 12.2 | 29.5491 | 0.1802 | -1.1991 | 22.234 | 24.099 | 26.379 | 29.549 | 33.698 | 38.726 | 45.654 |
| 12.4 | 30.1220 | 0.1810 | -1.2324 | 22.669 | 24.563 | 26.884 | 30.122 | 34.383 | 39.585 | 46.834 |
| 12.6 | 30.7105 | 0.1817 | -1.2658 | 23.119 | 25.042 | 27.404 | 30.710 | 35.085 | 40.465 | 48.047 |
| 12.8 | 31.3134 | 0.1823 | -1.2989 | 23.582 | 25.534 | 27.938 | 31.313 | 35.802 | 41.365 | 49.293 |
| 13.0 | 31.9321 | 0.1828 | -1.3322 | 24.060 | 26.042 | 28.487 | 31.932 | 36.536 | 42.286 | 50.574 |
| 13.2 | 32.5661 | 0.1833 | -1.3651 | 24.552 | 26.564 | 29.050 | 32.566 | 37.288 | 43.227 | 51.889 |
| 13.4 | 33.2153 | 0.1837 | -1.3978 | 25.058 | 27.100 | 29.629 | 33.215 | 38.055 | 44.188 | 53.236 |
| 13.6 | 33.8797 | 0.1840 | -1.4305 | 25.579 | 27.651 | 30.222 | 33.879 | 38.838 | 45.169 | 54.616 |
| 13.8 | 34.5578 | 0.1842 | -1.4632 | 26.113 | 28.215 | 30.829 | 34.557 | 39.636 | 46.167 | 56.028 |
| 14.0 | 35.2508 | 0.1844 | -1.4950 | 26.660 | 28.793 | 31.450 | 35.249 | 40.450 | 47.184 | 57.470 |
| 14.2 | 35.9564 | 0.1845 | -1.5269 | 27.220 | 29.384 | 32.083 | 35.954 | 41.276 | 48.217 | 58.940 |
| 14.4 | 36.6743 | 0.1845 | -1.5585 | 27.792 | 29.987 | 32.729 | 36.671 | 42.115 | 49.264 | 60.436 |
| 14.6 | 37.4017 | 0.1845 | -1.5896 | 28.374 | 30.599 | 33.384 | 37.397 | 42.964 | 50.322 | 61.954 |
| 14.8 | 38.1383 | 0.1845 | -1.6201 | 28.965 | 31.221 | 34.048 | 38.132 | 43.821 | 51.391 | 63.491 |
| 15.0 | 38.8813 | 0.1844 | -1.6501 | 29.563 | 31.850 | 34.718 | 38.874 | 44.684 | 52.467 | 65.045 |
| 15.2 | 39.6300 | 0.1842 | -1.6799 | 30.169 | 32.485 | 35.395 | 39.620 | 45.552 | 53.548 | 66.611 |
| 15.4 | 40.3832 | 0.1840 | -1.7087 | 30.779 | 33.125 | 36.077 | 40.371 | 46.423 | 54.633 | 68.187 |
| 15.6 | 41.1391 | 0.1838 | -1.7371 | 31.394 | 33.769 | 36.762 | 41.124 | 47.296 | 55.719 | 69.770 |
| 15.8 | 41.8970 | 0.1836 | -1.7647 | 32.012 | 34.416 | 37.449 | 41.879 | 48.169 | 56.805 | 71.355 |
| 16.0 | 42.6546 | 0.1833 | -1.7919 | 32.633 | 35.065 | 38.136 | 42.633 | 49.040 | 57.887 | 72.939 |
| 16.2 | 43.4115 | 0.1830 | -1.8182 | 33.254 | 35.714 | 38.824 | 43.386 | 49.908 | 58.966 | 74.521 |
| 16.4 | 44.1664 | 0.1826 | -1.8440 | 33.876 | 36.363 | 39.511 | 44.136 | 50.772 | 60.038 | 76.095 |
| 16.6 | 44.9189 | 0.1823 | -1.8692 | 34.498 | 37.011 | 40.196 | 44.884 | 51.631 | 61.104 | 77.662 |
| 16.8 | 45.6677 | 0.1819 | -1.8937 | 35.119 | 37.658 | 40.878 | 45.627 | 52.484 | 62.161 | 79.216 |
| 17.0 | 46.4119 | 0.1815 | -1.9175 | 35.737 | 38.302 | 41.557 | 46.366 | 53.330 | 63.208 | 80.758 |
| 17.2 | 47.1514 | 0.1810 | -1.9407 | 36.354 | 38.943 | 42.232 | 47.099 | 54.169 | 64.245 | 82.284 |
| 17.4 | 47.8860 | 0.1806 | -1.9631 | 36.968 | 39.581 | 42.903 | 47.827 | 55.000 | 65.272 | 83.794 |
| 17.6 | 48.6158 | 0.1801 | -1.9850 | 37.580 | 40.216 | 43.570 | 48.550 | 55.824 | 66.289 | 85.288 |
| 17.8 | 49.3411 | 0.1797 | -2.0062 | 38.190 | 40.848 | 44.234 | 49.267 | 56.641 | 67.296 | 86.764 |
| 18.0 | 50.0612 | 0.1792 | -2.0269 | 38.797 | 41.476 | 44.893 | 49.980 | 57.451 | 68.292 | 88.224 |
| 18.2 | 50.7774 | 0.1787 | -2.0468 | 39.402 | 42.102 | 45.549 | 50.688 | 58.254 | 69.280 | 89.666 |
| 18.4 | 51.4890 | 0.1782 | -2.0663 | 40.004 | 42.726 | 46.201 | 51.391 | 59.051 | 70.258 | 91.092 |
| 18.6 | 52.1974 | 0.1777 | -2.0853 | 40.605 | 43.347 | 46.851 | 52.090 | 59.843 | 71.228 | 92.502 |
| 18.8 | 52.9024 | 0.1772 | -2.1035 | 41.204 | 43.965 | 47.498 | 52.786 | 60.629 | 72.191 | 93.895 |
| 19.0 | 53.6046 | 0.1767 | -2.1213 | 41.802 | 44.583 | 48.142 | 53.478 | 61.411 | 73.147 | 95.274 |
| 19.2 | 54.3050 | 0.1762 | -2.1385 | 42.399 | 45.199 | 48.785 | 54.168 | 62.190 | 74.097 | 96.638 |
| 19.4 | 55.0033 | 0.1757 | -2.1553 | 42.995 | 45.814 | 49.427 | 54.856 | 62.964 | 75.041 | 97.987 |
| 19.6 | 55.6999 | 0.1752 | -2.1716 | 43.591 | 46.428 | 50.067 | 55.542 | 63.736 | 75.979 | 99.322 |
| 19.8 | 56.3957 | 0.1747 | -2.1874 | 44.187 | 47.042 | 50.707 | 56.227 | 64.506 | 76.912 | 100.642 |

**Table.** Height (cm) for females with NS: Estimated M, S, L parameters from centile values by age (3rd, 10th, 25th, 50th, 75th, 90th, 97th)

| age | M | S | L | p_3 | p_10 | p_25 | p_50 | p_75 | p_90 | p_97 |
| --- | --- | --- | --- | --- | --- | --- | --- | --- | --- | --- |
| 0.0 | 50.7311 | 0.0534 | -1.8734 | 46.274 | 47.570 | 48.995 | 50.731 | 52.657 | 54.582 | 56.696 |
| 0.2 | 54.1081 | 0.0533 | -1.8272 | 49.346 | 50.733 | 52.255 | 54.108 | 56.160 | 58.207 | 60.450 |
| 0.4 | 57.4114 | 0.0533 | -1.7812 | 52.350 | 53.826 | 55.444 | 57.411 | 59.586 | 61.752 | 64.119 |
| 0.6 | 60.6143 | 0.0533 | -1.7370 | 55.262 | 56.824 | 58.536 | 60.614 | 62.908 | 65.188 | 67.675 |
| 0.8 | 63.6920 | 0.0533 | -1.6896 | 58.058 | 59.705 | 61.507 | 63.692 | 66.099 | 68.487 | 71.087 |
| 1.0 | 66.6202 | 0.0533 | -1.6461 | 60.718 | 62.445 | 64.334 | 66.620 | 69.135 | 71.626 | 74.332 |
| 1.2 | 69.3923 | 0.0533 | -1.5998 | 63.234 | 65.038 | 67.010 | 69.392 | 72.009 | 74.595 | 77.400 |
| 1.4 | 72.0056 | 0.0533 | -1.5542 | 65.604 | 67.482 | 69.532 | 72.006 | 74.717 | 77.394 | 80.290 |
| 1.6 | 74.4572 | 0.0533 | -1.5094 | 67.826 | 69.775 | 71.898 | 74.457 | 77.258 | 80.018 | 82.998 |
| 1.8 | 76.7449 | 0.0532 | -1.4627 | 69.898 | 71.913 | 74.105 | 76.745 | 79.629 | 82.464 | 85.521 |
| 2.0 | 78.8675 | 0.0532 | -1.4183 | 71.820 | 73.896 | 76.154 | 78.867 | 81.828 | 84.733 | 87.859 |
| 2.2 | 80.8366 | 0.0532 | -1.3720 | 73.600 | 75.735 | 78.053 | 80.837 | 83.867 | 86.837 | 90.025 |
| 2.4 | 82.6681 | 0.0532 | -1.3247 | 75.254 | 77.444 | 79.820 | 82.668 | 85.764 | 88.792 | 92.036 |
| 2.6 | 84.3759 | 0.0532 | -1.2809 | 76.796 | 79.038 | 81.468 | 84.376 | 87.532 | 90.614 | 93.910 |
| 2.8 | 85.9762 | 0.0532 | -1.2351 | 78.239 | 80.531 | 83.011 | 85.976 | 89.189 | 92.320 | 95.662 |
| 3.0 | 87.4844 | 0.0532 | -1.1899 | 79.598 | 81.937 | 84.466 | 87.484 | 90.750 | 93.926 | 97.311 |
| 3.2 | 88.9103 | 0.0532 | -1.1444 | 80.881 | 83.266 | 85.841 | 88.910 | 92.225 | 95.445 | 98.868 |
| 3.4 | 90.2646 | 0.0531 | -1.0992 | 82.099 | 84.527 | 87.147 | 90.265 | 93.626 | 96.885 | 100.345 |
| 3.6 | 91.5560 | 0.0531 | -1.0542 | 83.259 | 85.730 | 88.392 | 91.556 | 94.962 | 98.258 | 101.751 |
| 3.8 | 92.7958 | 0.0531 | -1.0068 | 84.371 | 86.883 | 89.587 | 92.796 | 96.244 | 99.575 | 103.098 |
| 4.0 | 93.9917 | 0.0531 | -0.9618 | 85.443 | 87.996 | 90.740 | 93.992 | 97.480 | 100.845 | 104.397 |
| 4.2 | 95.1494 | 0.0531 | -0.9171 | 86.480 | 89.073 | 91.856 | 95.149 | 98.677 | 102.074 | 105.653 |
| 4.4 | 96.2732 | 0.0531 | -0.8718 | 87.486 | 90.117 | 92.939 | 96.273 | 99.839 | 103.266 | 106.871 |
| 4.6 | 97.3671 | 0.0531 | -0.8247 | 88.463 | 91.134 | 93.993 | 97.367 | 100.969 | 104.425 | 108.054 |
| 4.8 | 98.4342 | 0.0530 | -0.7801 | 89.417 | 92.125 | 95.021 | 98.434 | 102.072 | 105.556 | 109.208 |
| 5.0 | 99.4793 | 0.0530 | -0.7347 | 90.350 | 93.095 | 96.028 | 99.479 | 103.152 | 106.663 | 110.337 |
| 5.2 | 100.5063 | 0.0530 | -0.6893 | 91.266 | 94.049 | 97.018 | 100.506 | 104.212 | 107.750 | 111.445 |
| 5.4 | 101.5198 | 0.0530 | -0.6440 | 92.169 | 94.989 | 97.994 | 101.520 | 105.259 | 108.823 | 112.539 |
| 5.6 | 102.5249 | 0.0530 | -0.5983 | 93.064 | 95.922 | 98.962 | 102.525 | 106.297 | 109.886 | 113.622 |
| 5.8 | 103.5258 | 0.0530 | -0.5523 | 93.955 | 96.850 | 99.926 | 103.526 | 107.331 | 110.944 | 114.700 |
| 6.0 | 104.5273 | 0.0530 | -0.5065 | 94.846 | 97.779 | 100.891 | 104.527 | 108.365 | 112.003 | 115.778 |
| 6.2 | 105.5321 | 0.0530 | -0.4615 | 95.740 | 98.710 | 101.859 | 105.532 | 109.402 | 113.066 | 116.860 |
| 6.4 | 106.5422 | 0.0529 | -0.4167 | 96.638 | 99.647 | 102.832 | 106.542 | 110.445 | 114.134 | 117.948 |
| 6.6 | 107.5605 | 0.0529 | -0.3693 | 97.542 | 100.590 | 103.812 | 107.560 | 111.497 | 115.210 | 119.043 |
| 6.8 | 108.5883 | 0.0529 | -0.3245 | 98.456 | 101.543 | 104.802 | 108.588 | 112.558 | 116.296 | 120.149 |
| 7.0 | 109.6281 | 0.0529 | -0.2782 | 99.379 | 102.506 | 105.804 | 109.628 | 113.631 | 117.395 | 121.268 |
| 7.2 | 110.6793 | 0.0529 | -0.2338 | 100.313 | 103.480 | 106.816 | 110.679 | 114.717 | 118.506 | 122.400 |
| 7.4 | 111.7413 | 0.0529 | -0.1869 | 101.255 | 104.464 | 107.839 | 111.741 | 115.813 | 119.628 | 123.542 |
| 7.6 | 112.8119 | 0.0529 | -0.1432 | 102.206 | 105.456 | 108.870 | 112.812 | 116.918 | 120.760 | 124.695 |
| 7.8 | 113.8910 | 0.0529 | -0.0966 | 103.163 | 106.455 | 109.909 | 113.891 | 118.032 | 121.900 | 125.855 |
| 8.0 | 114.9762 | 0.0528 | -0.0508 | 104.125 | 107.460 | 110.954 | 114.976 | 119.152 | 123.047 | 127.022 |
| 8.2 | 116.0660 | 0.0528 | -0.0044 | 105.090 | 108.469 | 112.003 | 116.066 | 120.277 | 124.198 | 128.194 |
| 8.4 | 117.1575 | 0.0528 | 0.0396 | 106.058 | 109.480 | 113.054 | 117.158 | 121.403 | 125.351 | 129.368 |
| 8.6 | 118.2489 | 0.0528 | 0.0855 | 107.024 | 110.490 | 114.105 | 118.249 | 122.530 | 126.503 | 130.541 |
| 8.8 | 119.3382 | 0.0528 | 0.1310 | 107.988 | 111.498 | 115.154 | 119.338 | 123.654 | 127.653 | 131.711 |
| 9.0 | 120.4227 | 0.0528 | 0.1765 | 108.947 | 112.501 | 116.198 | 120.423 | 124.773 | 128.798 | 132.876 |
| 9.2 | 121.5030 | 0.0528 | 0.2235 | 109.901 | 113.500 | 117.238 | 121.503 | 125.887 | 129.938 | 134.034 |
| 9.4 | 122.5782 | 0.0528 | 0.2677 | 110.851 | 114.494 | 118.273 | 122.578 | 126.997 | 131.073 | 135.189 |
| 9.6 | 123.6512 | 0.0527 | 0.3140 | 111.798 | 115.486 | 119.306 | 123.651 | 128.104 | 132.204 | 136.339 |
| 9.8 | 124.7227 | 0.0527 | 0.3595 | 112.743 | 116.476 | 120.337 | 124.723 | 129.209 | 133.334 | 137.488 |
| 10.0 | 125.7938 | 0.0527 | 0.4054 | 113.687 | 117.465 | 121.368 | 125.794 | 130.314 | 134.464 | 138.636 |
| 10.2 | 126.8651 | 0.0527 | 0.4513 | 114.631 | 118.455 | 122.399 | 126.865 | 131.419 | 135.593 | 139.783 |
| 10.4 | 127.9362 | 0.0527 | 0.4965 | 115.574 | 119.444 | 123.430 | 127.936 | 132.524 | 136.722 | 140.931 |
| 10.6 | 129.0081 | 0.0527 | 0.5413 | 116.518 | 120.434 | 124.462 | 129.008 | 133.629 | 137.852 | 142.079 |
| 10.8 | 130.0808 | 0.0527 | 0.5876 | 117.461 | 121.424 | 125.494 | 130.081 | 134.735 | 138.982 | 143.227 |
| 11.0 | 131.1541 | 0.0527 | 0.6324 | 118.405 | 122.415 | 126.527 | 131.154 | 135.842 | 140.113 | 144.376 |
| 11.2 | 132.2267 | 0.0526 | 0.6781 | 119.347 | 123.404 | 127.559 | 132.227 | 136.948 | 141.243 | 145.524 |
| 11.4 | 133.2960 | 0.0526 | 0.7243 | 120.285 | 124.391 | 128.588 | 133.296 | 138.050 | 142.369 | 146.667 |
| 11.6 | 134.3583 | 0.0526 | 0.7695 | 121.217 | 125.370 | 129.610 | 134.358 | 139.146 | 143.487 | 147.803 |
| 11.8 | 135.4111 | 0.0526 | 0.8147 | 122.140 | 126.341 | 130.623 | 135.411 | 140.231 | 144.595 | 148.928 |
| 12.0 | 136.4511 | 0.0526 | 0.8608 | 123.050 | 127.299 | 131.623 | 136.451 | 141.303 | 145.690 | 150.038 |
| 12.2 | 137.4739 | 0.0526 | 0.9055 | 123.945 | 128.241 | 132.607 | 137.474 | 142.357 | 146.766 | 151.130 |
| 12.4 | 138.4763 | 0.0526 | 0.9523 | 124.820 | 129.164 | 133.571 | 138.476 | 143.390 | 147.819 | 152.197 |
| 12.6 | 139.4513 | 0.0526 | 0.9973 | 125.670 | 130.061 | 134.509 | 139.451 | 144.394 | 148.844 | 153.236 |
| 12.8 | 140.3953 | 0.0525 | 1.0424 | 126.492 | 130.929 | 135.416 | 140.395 | 145.367 | 149.835 | 154.240 |
| 13.0 | 141.3029 | 0.0525 | 1.0889 | 127.280 | 131.762 | 136.289 | 141.303 | 146.301 | 150.787 | 155.203 |
| 13.2 | 142.1713 | 0.0525 | 1.1342 | 128.033 | 132.559 | 137.124 | 142.171 | 147.195 | 151.697 | 156.123 |
| 13.4 | 142.9987 | 0.0525 | 1.1792 | 128.748 | 133.318 | 137.919 | 142.999 | 148.046 | 152.563 | 156.999 |
| 13.6 | 143.7840 | 0.0525 | 1.2254 | 129.424 | 134.037 | 138.673 | 143.784 | 148.854 | 153.385 | 157.827 |
| 13.8 | 144.5261 | 0.0525 | 1.2711 | 130.061 | 134.715 | 139.386 | 144.526 | 149.617 | 154.160 | 158.608 |
| 14.0 | 145.2232 | 0.0525 | 1.3158 | 130.658 | 135.352 | 140.055 | 145.223 | 150.334 | 154.887 | 159.340 |
| 14.2 | 145.8759 | 0.0524 | 1.3613 | 131.214 | 135.947 | 140.682 | 145.876 | 151.004 | 155.566 | 160.023 |
| 14.4 | 146.4860 | 0.0524 | 1.4079 | 131.730 | 136.502 | 141.267 | 146.486 | 151.630 | 156.200 | 160.658 |
| 14.6 | 147.0545 | 0.0524 | 1.4530 | 132.209 | 137.018 | 141.812 | 147.055 | 152.213 | 156.790 | 161.249 |
| 14.8 | 147.5834 | 0.0524 | 1.4990 | 132.652 | 137.497 | 142.319 | 147.583 | 152.756 | 157.337 | 161.795 |
| 15.0 | 148.0749 | 0.0524 | 1.5445 | 133.061 | 137.941 | 142.790 | 148.075 | 153.259 | 157.844 | 162.301 |
| 15.2 | 148.5301 | 0.0524 | 1.5899 | 133.437 | 138.351 | 143.226 | 148.530 | 153.725 | 158.313 | 162.767 |
| 15.4 | 148.9537 | 0.0524 | 1.6358 | 133.784 | 138.731 | 143.631 | 148.954 | 154.158 | 158.748 | 163.198 |
| 15.6 | 149.3476 | 0.0524 | 1.6805 | 134.105 | 139.084 | 144.008 | 149.348 | 154.560 | 159.152 | 163.597 |
| 15.8 | 149.7160 | 0.0523 | 1.7262 | 134.401 | 139.413 | 144.360 | 149.716 | 154.936 | 159.528 | 163.968 |
| 16.0 | 150.0620 | 0.0523 | 1.7721 | 134.677 | 139.721 | 144.690 | 150.062 | 155.289 | 159.880 | 164.314 |
| 16.2 | 150.3878 | 0.0523 | 1.8179 | 134.935 | 140.010 | 145.001 | 150.388 | 155.621 | 160.210 | 164.638 |
| 16.4 | 150.6947 | 0.0523 | 1.8628 | 135.176 | 140.281 | 145.294 | 150.695 | 155.933 | 160.521 | 164.942 |
| 16.6 | 150.9859 | 0.0523 | 1.9088 | 135.402 | 140.537 | 145.572 | 150.986 | 156.229 | 160.815 | 165.228 |
| 16.8 | 151.2610 | 0.0523 | 1.9542 | 135.613 | 140.779 | 145.834 | 151.261 | 156.508 | 161.092 | 165.497 |
| 17.0 | 151.5229 | 0.0523 | 2.0002 | 135.812 | 141.008 | 146.083 | 151.523 | 156.774 | 161.354 | 165.751 |
| 17.2 | 151.7721 | 0.0523 | 2.0458 | 135.999 | 141.225 | 146.320 | 151.772 | 157.027 | 161.603 | 165.992 |
| 17.4 | 152.0097 | 0.0522 | 2.0911 | 136.176 | 141.431 | 146.546 | 152.010 | 157.267 | 161.840 | 166.220 |
| 17.6 | 152.2371 | 0.0522 | 2.1365 | 136.343 | 141.628 | 146.762 | 152.237 | 157.497 | 162.066 | 166.437 |
| 17.8 | 152.4550 | 0.0522 | 2.1820 | 136.501 | 141.815 | 146.969 | 152.455 | 157.717 | 162.282 | 166.644 |
| 18.0 | 152.6638 | 0.0522 | 2.2274 | 136.651 | 141.994 | 147.167 | 152.664 | 157.928 | 162.488 | 166.841 |
| 18.2 | 152.8653 | 0.0522 | 2.2730 | 136.793 | 142.166 | 147.358 | 152.865 | 158.131 | 162.687 | 167.030 |
| 18.4 | 153.0609 | 0.0522 | 2.3185 | 136.931 | 142.333 | 147.543 | 153.061 | 158.328 | 162.879 | 167.212 |
| 18.6 | 153.2529 | 0.0522 | 2.3643 | 137.064 | 142.496 | 147.725 | 153.253 | 158.521 | 163.067 | 167.391 |
| 18.8 | 153.4421 | 0.0522 | 2.4094 | 137.195 | 142.656 | 147.904 | 153.442 | 158.712 | 163.253 | 167.567 |
| 19.0 | 153.6311 | 0.0521 | 2.4555 | 137.325 | 142.816 | 148.083 | 153.631 | 158.902 | 163.438 | 167.742 |
| 19.2 | 153.8212 | 0.0521 | 2.5011 | 137.456 | 142.977 | 148.263 | 153.821 | 159.094 | 163.624 | 167.919 |
| 19.4 | 154.0121 | 0.0521 | 2.5462 | 137.587 | 143.139 | 148.443 | 154.012 | 159.286 | 163.812 | 168.097 |
| 19.6 | 154.2040 | 0.0521 | 2.5924 | 137.718 | 143.301 | 148.625 | 154.204 | 159.479 | 164.000 | 168.276 |
| 19.8 | 154.3966 | 0.0521 | 2.6379 | 137.850 | 143.464 | 148.807 | 154.397 | 159.673 | 164.189 | 168.456 |

**Table.** Height (cm) for males with NS: Estimated M, S, L parameters from centile values by age (3rd, 10th, 25th, 50th, 75th, 90th, 97th)

| age | M | S | L | p_3 | p_10 | p_25 | p_50 | p_75 | p_90 | p_97 |
| --- | --- | --- | --- | --- | --- | --- | --- | --- | --- | --- |
| 0.0 | 50.8818 | 0.0881 | -0.6696 | 43.485 | 45.636 | 48.002 | 50.882 | 54.062 | 57.218 | 60.648 |
| 0.2 | 54.1769 | 0.0855 | -0.6960 | 46.518 | 48.748 | 51.199 | 54.177 | 57.461 | 60.717 | 64.250 |
| 0.4 | 57.4105 | 0.0830 | -0.7231 | 49.517 | 51.819 | 54.345 | 57.410 | 60.787 | 64.130 | 67.753 |
| 0.6 | 60.5611 | 0.0806 | -0.7498 | 52.462 | 54.827 | 57.419 | 60.561 | 64.017 | 67.434 | 71.133 |
| 0.8 | 63.6068 | 0.0783 | -0.7772 | 55.332 | 57.751 | 60.400 | 63.607 | 67.129 | 70.608 | 74.369 |
| 1.0 | 66.5272 | 0.0760 | -0.8045 | 58.105 | 60.570 | 63.267 | 66.527 | 70.104 | 73.632 | 77.442 |
| 1.2 | 69.3131 | 0.0739 | -0.8317 | 60.772 | 63.275 | 66.010 | 69.313 | 72.932 | 76.498 | 80.343 |
| 1.4 | 71.9578 | 0.0718 | -0.8586 | 63.323 | 65.856 | 68.621 | 71.958 | 75.609 | 79.201 | 83.071 |
| 1.6 | 74.4543 | 0.0699 | -0.8857 | 65.749 | 68.306 | 71.094 | 74.454 | 78.127 | 81.737 | 85.621 |
| 1.8 | 76.7961 | 0.0680 | -0.9130 | 68.042 | 70.616 | 73.420 | 76.796 | 80.482 | 84.101 | 87.991 |
| 2.0 | 78.9791 | 0.0663 | -0.9392 | 70.195 | 72.781 | 75.595 | 78.979 | 82.670 | 86.290 | 90.178 |
| 2.2 | 81.0099 | 0.0646 | -0.9662 | 72.213 | 74.804 | 77.623 | 81.010 | 84.700 | 88.316 | 92.195 |
| 2.4 | 82.8983 | 0.0631 | -0.9935 | 74.101 | 76.695 | 79.514 | 82.898 | 86.583 | 90.189 | 94.056 |
| 2.6 | 84.6560 | 0.0617 | -1.0201 | 75.868 | 78.461 | 81.277 | 84.656 | 88.331 | 91.926 | 95.776 |
| 2.8 | 86.2930 | 0.0603 | -1.0481 | 77.522 | 80.112 | 82.923 | 86.293 | 89.956 | 93.537 | 97.370 |
| 3.0 | 87.8203 | 0.0591 | -1.0744 | 79.070 | 81.655 | 84.460 | 87.820 | 91.471 | 95.036 | 98.851 |
| 3.2 | 89.2501 | 0.0579 | -1.1019 | 80.523 | 83.103 | 85.900 | 89.250 | 92.887 | 96.438 | 100.235 |
| 3.4 | 90.5948 | 0.0569 | -1.1280 | 81.891 | 84.465 | 87.255 | 90.595 | 94.219 | 97.755 | 101.536 |
| 3.6 | 91.8670 | 0.0559 | -1.1559 | 83.186 | 85.755 | 88.537 | 91.867 | 95.479 | 99.003 | 102.769 |
| 3.8 | 93.0800 | 0.0550 | -1.1827 | 84.419 | 86.982 | 89.759 | 93.080 | 96.682 | 100.195 | 103.949 |
| 4.0 | 94.2454 | 0.0543 | -1.2085 | 85.599 | 88.159 | 90.930 | 94.245 | 97.840 | 101.345 | 105.089 |
| 4.2 | 95.3709 | 0.0535 | -1.2365 | 86.736 | 89.292 | 92.060 | 95.371 | 98.960 | 102.461 | 106.199 |
| 4.4 | 96.4642 | 0.0529 | -1.2641 | 87.834 | 90.389 | 93.156 | 96.464 | 100.051 | 103.550 | 107.287 |
| 4.6 | 97.5320 | 0.0523 | -1.2903 | 88.901 | 91.456 | 94.223 | 97.532 | 101.120 | 104.619 | 108.358 |
| 4.8 | 98.5811 | 0.0518 | -1.3171 | 89.942 | 92.500 | 95.269 | 98.581 | 102.173 | 105.677 | 109.422 |
| 5.0 | 99.6176 | 0.0514 | -1.3445 | 90.965 | 93.526 | 96.299 | 99.618 | 103.217 | 106.729 | 110.484 |
| 5.2 | 100.6440 | 0.0510 | -1.3713 | 91.970 | 94.537 | 97.317 | 100.644 | 104.254 | 107.778 | 111.547 |
| 5.4 | 101.6619 | 0.0507 | -1.3989 | 92.961 | 95.535 | 98.323 | 101.662 | 105.286 | 108.825 | 112.612 |
| 5.6 | 102.6716 | 0.0504 | -1.4261 | 93.937 | 96.520 | 99.319 | 102.672 | 106.312 | 109.870 | 113.679 |
| 5.8 | 103.6741 | 0.0502 | -1.4525 | 94.900 | 97.493 | 100.305 | 103.674 | 107.335 | 110.914 | 114.748 |
| 6.0 | 104.6705 | 0.0500 | -1.4796 | 95.851 | 98.456 | 101.283 | 104.670 | 108.354 | 111.958 | 115.821 |
| 6.2 | 105.6601 | 0.0498 | -1.5056 | 96.789 | 99.409 | 102.251 | 105.660 | 109.369 | 113.000 | 116.896 |
| 6.4 | 106.6418 | 0.0497 | -1.5340 | 97.716 | 100.350 | 103.210 | 106.642 | 110.378 | 114.040 | 117.972 |
| 6.6 | 107.6150 | 0.0497 | -1.5606 | 98.629 | 101.279 | 104.158 | 107.615 | 111.382 | 115.076 | 119.047 |
| 6.8 | 108.5789 | 0.0496 | -1.5871 | 99.529 | 102.196 | 105.095 | 108.579 | 112.378 | 116.107 | 120.119 |
| 7.0 | 109.5319 | 0.0496 | -1.6143 | 100.415 | 103.100 | 106.020 | 109.532 | 113.365 | 117.131 | 121.187 |
| 7.2 | 110.4741 | 0.0496 | -1.6424 | 101.287 | 103.991 | 106.933 | 110.474 | 114.343 | 118.148 | 122.251 |
| 7.4 | 111.4073 | 0.0496 | -1.6688 | 102.147 | 104.870 | 107.836 | 111.407 | 115.313 | 119.159 | 123.310 |
| 7.6 | 112.3313 | 0.0497 | -1.6960 | 102.996 | 105.739 | 108.728 | 112.331 | 116.276 | 120.163 | 124.365 |
| 7.8 | 113.2470 | 0.0498 | -1.7225 | 103.834 | 106.598 | 109.611 | 113.247 | 117.231 | 121.162 | 125.416 |
| 8.0 | 114.1547 | 0.0498 | -1.7495 | 104.663 | 107.447 | 110.486 | 114.155 | 118.179 | 122.155 | 126.463 |
| 8.2 | 115.0559 | 0.0499 | -1.7767 | 105.484 | 108.289 | 111.353 | 115.056 | 119.122 | 123.144 | 127.507 |
| 8.4 | 115.9511 | 0.0500 | -1.8045 | 106.298 | 109.124 | 112.214 | 115.951 | 120.060 | 124.129 | 128.549 |
| 8.6 | 116.8431 | 0.0501 | -1.8305 | 107.107 | 109.955 | 113.071 | 116.843 | 120.995 | 125.111 | 129.589 |
| 8.8 | 117.7318 | 0.0503 | -1.8565 | 107.912 | 110.782 | 113.924 | 117.732 | 121.927 | 126.092 | 130.628 |
| 9.0 | 118.6190 | 0.0504 | -1.8843 | 108.716 | 111.608 | 114.776 | 118.619 | 122.858 | 127.072 | 131.668 |
| 9.2 | 119.5051 | 0.0505 | -1.9114 | 109.518 | 112.432 | 115.626 | 119.505 | 123.789 | 128.052 | 132.709 |
| 9.4 | 120.3912 | 0.0506 | -1.9393 | 110.321 | 113.257 | 116.477 | 120.391 | 124.719 | 129.032 | 133.750 |
| 9.6 | 121.2783 | 0.0507 | -1.9654 | 111.125 | 114.082 | 117.329 | 121.278 | 125.650 | 130.013 | 134.791 |
| 9.8 | 122.1662 | 0.0508 | -1.9931 | 111.931 | 114.909 | 118.182 | 122.166 | 126.582 | 130.994 | 135.834 |
| 10.0 | 123.0561 | 0.0509 | -2.0196 | 112.740 | 115.739 | 119.037 | 123.056 | 127.515 | 131.976 | 136.876 |
| 10.2 | 123.9482 | 0.0510 | -2.0468 | 113.553 | 116.573 | 119.895 | 123.948 | 128.450 | 132.959 | 137.919 |
| 10.4 | 124.8443 | 0.0511 | -2.0736 | 114.372 | 117.411 | 120.758 | 124.844 | 129.388 | 133.944 | 138.963 |
| 10.6 | 125.7438 | 0.0511 | -2.1005 | 115.196 | 118.255 | 121.625 | 125.744 | 130.328 | 134.931 | 140.008 |
| 10.8 | 126.6490 | 0.0512 | -2.1263 | 116.028 | 119.105 | 122.499 | 126.649 | 131.273 | 135.921 | 141.054 |
| 11.0 | 127.5594 | 0.0512 | -2.1545 | 116.869 | 119.964 | 123.379 | 127.559 | 132.222 | 136.914 | 142.102 |
| 11.2 | 128.4773 | 0.0512 | -2.1811 | 117.719 | 120.831 | 124.268 | 128.477 | 133.177 | 137.911 | 143.153 |
| 11.4 | 129.4037 | 0.0512 | -2.2084 | 118.580 | 121.709 | 125.166 | 129.404 | 134.139 | 138.914 | 144.209 |
| 11.6 | 130.3408 | 0.0512 | -2.2360 | 119.455 | 122.600 | 126.076 | 130.341 | 135.110 | 139.926 | 145.270 |
| 11.8 | 131.2903 | 0.0512 | -2.2627 | 120.344 | 123.505 | 127.000 | 131.290 | 136.093 | 140.946 | 146.339 |
| 12.0 | 132.2530 | 0.0511 | -2.2901 | 121.250 | 124.425 | 127.938 | 132.253 | 137.087 | 141.977 | 147.416 |
| 12.2 | 133.2309 | 0.0510 | -2.3169 | 122.173 | 125.362 | 128.892 | 133.231 | 138.095 | 143.020 | 148.503 |
| 12.4 | 134.2238 | 0.0509 | -2.3437 | 123.113 | 126.316 | 129.862 | 134.224 | 139.117 | 144.075 | 149.601 |
| 12.6 | 135.2321 | 0.0508 | -2.3701 | 124.071 | 127.286 | 130.849 | 135.232 | 140.153 | 145.143 | 150.709 |
| 12.8 | 136.2557 | 0.0507 | -2.3979 | 125.047 | 128.274 | 131.852 | 136.256 | 141.203 | 146.223 | 151.829 |
| 13.0 | 137.2956 | 0.0505 | -2.4250 | 126.041 | 129.280 | 132.872 | 137.296 | 142.268 | 147.317 | 152.960 |
| 13.2 | 138.3503 | 0.0504 | -2.4516 | 127.051 | 130.302 | 133.908 | 138.350 | 143.347 | 148.424 | 154.102 |
| 13.4 | 139.4188 | 0.0502 | -2.4779 | 128.077 | 131.339 | 134.958 | 139.419 | 144.438 | 149.542 | 155.253 |
| 13.6 | 140.4984 | 0.0500 | -2.5057 | 129.117 | 132.389 | 136.021 | 140.498 | 145.540 | 150.668 | 156.412 |
| 13.8 | 141.5881 | 0.0498 | -2.5325 | 130.168 | 133.450 | 137.094 | 141.588 | 146.650 | 151.803 | 157.577 |
| 14.0 | 142.6857 | 0.0496 | -2.5603 | 131.230 | 134.521 | 138.176 | 142.686 | 147.767 | 152.943 | 158.746 |
| 14.2 | 143.7887 | 0.0494 | -2.5868 | 132.298 | 135.599 | 139.264 | 143.789 | 148.889 | 154.085 | 159.916 |
| 14.4 | 144.8929 | 0.0492 | -2.6138 | 133.370 | 136.679 | 140.355 | 144.893 | 150.010 | 155.227 | 161.083 |
| 14.6 | 145.9937 | 0.0490 | -2.6406 | 134.441 | 137.758 | 141.443 | 145.994 | 151.127 | 156.362 | 162.242 |
| 14.8 | 147.0877 | 0.0487 | -2.6679 | 135.508 | 138.832 | 142.525 | 147.088 | 152.236 | 157.488 | 163.390 |
| 15.0 | 148.1711 | 0.0485 | -2.6939 | 136.566 | 139.897 | 143.598 | 148.171 | 153.333 | 158.600 | 164.522 |
| 15.2 | 149.2392 | 0.0483 | -2.7212 | 137.613 | 140.949 | 144.657 | 149.239 | 154.413 | 159.694 | 165.634 |
| 15.4 | 150.2898 | 0.0480 | -2.7483 | 138.645 | 141.986 | 145.700 | 150.290 | 155.473 | 160.767 | 166.723 |
| 15.6 | 151.3184 | 0.0477 | -2.7758 | 139.659 | 143.003 | 146.722 | 151.318 | 156.511 | 161.815 | 167.785 |
| 15.8 | 152.3222 | 0.0475 | -2.8024 | 140.650 | 143.998 | 147.720 | 152.322 | 157.522 | 162.835 | 168.817 |
| 16.0 | 153.2984 | 0.0472 | -2.8291 | 141.617 | 144.967 | 148.692 | 153.298 | 158.504 | 163.824 | 169.816 |
| 16.2 | 154.2438 | 0.0470 | -2.8569 | 142.557 | 145.908 | 149.635 | 154.244 | 159.453 | 164.779 | 170.780 |
| 16.4 | 155.1567 | 0.0467 | -2.8832 | 143.467 | 146.818 | 150.546 | 155.157 | 160.369 | 165.699 | 171.706 |
| 16.6 | 156.0357 | 0.0465 | -2.9113 | 144.347 | 147.697 | 151.425 | 156.036 | 161.249 | 166.582 | 172.594 |
| 16.8 | 156.8801 | 0.0462 | -2.9373 | 145.194 | 148.543 | 152.270 | 156.880 | 162.094 | 167.427 | 173.442 |
| 17.0 | 157.6872 | 0.0460 | -2.9653 | 146.007 | 149.354 | 153.079 | 157.687 | 162.900 | 168.234 | 174.251 |
| 17.2 | 158.4578 | 0.0457 | -2.9911 | 146.785 | 150.130 | 153.852 | 158.458 | 163.668 | 169.001 | 175.018 |
| 17.4 | 159.1939 | 0.0455 | -3.0177 | 147.531 | 150.873 | 154.592 | 159.194 | 164.401 | 169.731 | 175.748 |
| 17.6 | 159.8969 | 0.0452 | -3.0451 | 148.247 | 151.585 | 155.299 | 159.897 | 165.100 | 170.427 | 176.442 |
| 17.8 | 160.5690 | 0.0450 | -3.0736 | 148.934 | 152.267 | 155.977 | 160.569 | 165.767 | 171.091 | 177.104 |
| 18.0 | 161.2137 | 0.0448 | -3.1002 | 149.594 | 152.923 | 156.627 | 161.214 | 166.406 | 171.725 | 177.735 |
| 18.2 | 161.8327 | 0.0445 | -3.1261 | 150.230 | 153.553 | 157.252 | 161.833 | 167.019 | 172.333 | 178.338 |
| 18.4 | 162.4293 | 0.0443 | -3.1538 | 150.845 | 154.163 | 157.856 | 162.429 | 167.609 | 172.917 | 178.918 |
| 18.6 | 163.0073 | 0.0441 | -3.1806 | 151.442 | 154.754 | 158.441 | 163.007 | 168.180 | 173.482 | 179.478 |
| 18.8 | 163.5703 | 0.0439 | -3.2073 | 152.025 | 155.331 | 159.012 | 163.570 | 168.735 | 174.031 | 180.021 |
| 19.0 | 164.1220 | 0.0436 | -3.2345 | 152.597 | 155.897 | 159.571 | 164.122 | 169.279 | 174.568 | 180.553 |
| 19.2 | 164.6647 | 0.0434 | -3.2617 | 153.161 | 156.454 | 160.121 | 164.665 | 169.814 | 175.096 | 181.075 |
| 19.4 | 165.2009 | 0.0432 | -3.2879 | 153.718 | 157.005 | 160.665 | 165.201 | 170.342 | 175.617 | 181.589 |
| 19.6 | 165.7311 | 0.0430 | -3.3156 | 154.270 | 157.550 | 161.204 | 165.731 | 170.864 | 176.132 | 182.098 |
| 19.8 | 166.2587 | 0.0428 | -3.3424 | 154.819 | 158.093 | 161.739 | 166.259 | 171.383 | 176.644 | 182.603 |

**Table.** BMI (kg/m²) for females with NS: Estimated M, S, L parameters from centile values by age (3rd, 10th, 25th, 50th, 75th, 90th, 97th)

| age | M | S | L | p_3 | p_10 | p_25 | p_50 | p_75 | p_90 | p_97 |
| --- | --- | --- | --- | --- | --- | --- | --- | --- | --- | --- |
| 0.0 | 14.1497 | 0.0924 | 0.0205 | 11.889 | 12.568 | 13.294 | 14.150 | 15.059 | 15.926 | 16.830 |
| 0.2 | 14.3311 | 0.0929 | 0.0125 | 12.032 | 12.722 | 13.461 | 14.331 | 15.257 | 16.141 | 17.063 |
| 0.4 | 14.5076 | 0.0933 | 0.0053 | 12.171 | 12.872 | 13.622 | 14.508 | 15.450 | 16.350 | 17.290 |
| 0.6 | 14.6768 | 0.0938 | 0.0008 | 12.303 | 13.015 | 13.777 | 14.677 | 15.635 | 16.551 | 17.508 |
| 0.8 | 14.8368 | 0.0943 | -0.0077 | 12.428 | 13.149 | 13.923 | 14.837 | 15.811 | 16.743 | 17.717 |
| 1.0 | 14.9866 | 0.0947 | -0.0159 | 12.544 | 13.275 | 14.059 | 14.987 | 15.976 | 16.923 | 17.914 |
| 1.2 | 15.1242 | 0.0952 | -0.0213 | 12.649 | 13.389 | 14.184 | 15.124 | 16.128 | 17.090 | 18.096 |
| 1.4 | 15.2486 | 0.0957 | -0.0290 | 12.743 | 13.492 | 14.296 | 15.249 | 16.266 | 17.242 | 18.264 |
| 1.6 | 15.3588 | 0.0962 | -0.0348 | 12.825 | 13.582 | 14.395 | 15.359 | 16.389 | 17.378 | 18.414 |
| 1.8 | 15.4539 | 0.0966 | -0.0410 | 12.894 | 13.658 | 14.480 | 15.454 | 16.496 | 17.497 | 18.547 |
| 2.0 | 15.5324 | 0.0971 | -0.0485 | 12.949 | 13.720 | 14.549 | 15.532 | 16.586 | 17.598 | 18.661 |
| 2.2 | 15.5948 | 0.0976 | -0.0562 | 12.991 | 13.767 | 14.603 | 15.595 | 16.658 | 17.681 | 18.756 |
| 2.4 | 15.6427 | 0.0981 | -0.0614 | 13.021 | 13.801 | 14.643 | 15.643 | 16.715 | 17.747 | 18.832 |
| 2.6 | 15.6757 | 0.0986 | -0.0694 | 13.038 | 13.823 | 14.669 | 15.676 | 16.756 | 17.797 | 18.892 |
| 2.8 | 15.6953 | 0.0991 | -0.0736 | 13.043 | 13.832 | 14.683 | 15.695 | 16.783 | 17.831 | 18.935 |
| 3.0 | 15.7012 | 0.0996 | -0.0825 | 13.038 | 13.829 | 14.684 | 15.701 | 16.795 | 17.851 | 18.963 |
| 3.2 | 15.6961 | 0.1001 | -0.0883 | 13.023 | 13.817 | 14.674 | 15.696 | 16.796 | 17.857 | 18.977 |
| 3.4 | 15.6812 | 0.1006 | -0.0949 | 13.000 | 13.795 | 14.656 | 15.681 | 16.786 | 17.853 | 18.980 |
| 3.6 | 15.6579 | 0.1011 | -0.1028 | 12.971 | 13.767 | 14.629 | 15.658 | 16.767 | 17.839 | 18.972 |
| 3.8 | 15.6280 | 0.1016 | -0.1123 | 12.936 | 13.733 | 14.597 | 15.628 | 16.741 | 17.818 | 18.958 |
| 4.0 | 15.5938 | 0.1021 | -0.1184 | 12.897 | 13.695 | 14.560 | 15.594 | 16.710 | 17.792 | 18.937 |
| 4.2 | 15.5562 | 0.1026 | -0.1242 | 12.855 | 13.654 | 14.520 | 15.556 | 16.676 | 17.762 | 18.912 |
| 4.4 | 15.5161 | 0.1031 | -0.1292 | 12.811 | 13.610 | 14.478 | 15.516 | 16.639 | 17.729 | 18.884 |
| 4.6 | 15.4743 | 0.1037 | -0.1369 | 12.766 | 13.566 | 14.434 | 15.474 | 16.601 | 17.694 | 18.855 |
| 4.8 | 15.4328 | 0.1042 | -0.1447 | 12.722 | 13.521 | 14.391 | 15.433 | 16.562 | 17.660 | 18.826 |
| 5.0 | 15.3920 | 0.1047 | -0.1499 | 12.677 | 13.478 | 14.348 | 15.392 | 16.524 | 17.626 | 18.797 |
| 5.2 | 15.3527 | 0.1052 | -0.1579 | 12.635 | 13.435 | 14.307 | 15.353 | 16.488 | 17.594 | 18.771 |
| 5.4 | 15.3155 | 0.1057 | -0.1651 | 12.594 | 13.395 | 14.267 | 15.315 | 16.455 | 17.565 | 18.747 |
| 5.6 | 15.2810 | 0.1063 | -0.1713 | 12.555 | 13.356 | 14.230 | 15.281 | 16.424 | 17.539 | 18.727 |
| 5.8 | 15.2495 | 0.1068 | -0.1796 | 12.519 | 13.321 | 14.196 | 15.250 | 16.396 | 17.516 | 18.711 |
| 6.0 | 15.2221 | 0.1073 | -0.1858 | 12.486 | 13.289 | 14.166 | 15.222 | 16.373 | 17.498 | 18.699 |
| 6.2 | 15.1993 | 0.1079 | -0.1922 | 12.456 | 13.261 | 14.140 | 15.199 | 16.355 | 17.485 | 18.694 |
| 6.4 | 15.1812 | 0.1084 | -0.1990 | 12.431 | 13.237 | 14.118 | 15.181 | 16.342 | 17.478 | 18.694 |
| 6.6 | 15.1691 | 0.1090 | -0.2051 | 12.410 | 13.218 | 14.102 | 15.169 | 16.335 | 17.478 | 18.702 |
| 6.8 | 15.1633 | 0.1095 | -0.2116 | 12.395 | 13.205 | 14.092 | 15.163 | 16.335 | 17.485 | 18.717 |
| 7.0 | 15.1646 | 0.1100 | -0.2189 | 12.386 | 13.198 | 14.088 | 15.165 | 16.343 | 17.500 | 18.742 |
| 7.2 | 15.1732 | 0.1106 | -0.2258 | 12.382 | 13.198 | 14.091 | 15.173 | 16.359 | 17.524 | 18.776 |
| 7.4 | 15.1891 | 0.1112 | -0.2338 | 12.385 | 13.203 | 14.101 | 15.189 | 16.383 | 17.557 | 18.820 |
| 7.6 | 15.2130 | 0.1117 | -0.2409 | 12.394 | 13.216 | 14.118 | 15.213 | 16.415 | 17.599 | 18.873 |
| 7.8 | 15.2440 | 0.1123 | -0.2458 | 12.408 | 13.234 | 14.142 | 15.244 | 16.455 | 17.649 | 18.935 |
| 8.0 | 15.2825 | 0.1128 | -0.2540 | 12.429 | 13.259 | 14.173 | 15.283 | 16.503 | 17.708 | 19.008 |
| 8.2 | 15.3290 | 0.1134 | -0.2579 | 12.455 | 13.291 | 14.211 | 15.329 | 16.560 | 17.776 | 19.089 |
| 8.4 | 15.3810 | 0.1140 | -0.2660 | 12.487 | 13.328 | 14.254 | 15.381 | 16.623 | 17.852 | 19.179 |
| 8.6 | 15.4395 | 0.1145 | -0.2741 | 12.524 | 13.370 | 14.303 | 15.440 | 16.693 | 17.935 | 19.278 |
| 8.8 | 15.5040 | 0.1151 | -0.2791 | 12.565 | 13.417 | 14.358 | 15.504 | 16.770 | 18.024 | 19.383 |
| 9.0 | 15.5728 | 0.1157 | -0.2871 | 12.610 | 13.468 | 14.416 | 15.573 | 16.852 | 18.120 | 19.496 |
| 9.2 | 15.6464 | 0.1162 | -0.2949 | 12.659 | 13.524 | 14.479 | 15.647 | 16.938 | 18.221 | 19.614 |
| 9.4 | 15.7243 | 0.1168 | -0.3001 | 12.710 | 13.582 | 14.546 | 15.724 | 17.030 | 18.328 | 19.738 |
| 9.6 | 15.8063 | 0.1174 | -0.3069 | 12.765 | 13.644 | 14.617 | 15.806 | 17.126 | 18.439 | 19.868 |
| 9.8 | 15.8918 | 0.1180 | -0.3141 | 12.823 | 13.709 | 14.690 | 15.892 | 17.226 | 18.555 | 20.003 |
| 10.0 | 15.9809 | 0.1186 | -0.3217 | 12.884 | 13.777 | 14.767 | 15.981 | 17.330 | 18.676 | 20.143 |
| 10.2 | 16.0730 | 0.1192 | -0.3273 | 12.946 | 13.847 | 14.847 | 16.073 | 17.437 | 18.800 | 20.287 |
| 10.4 | 16.1680 | 0.1198 | -0.3346 | 13.011 | 13.920 | 14.929 | 16.168 | 17.548 | 18.928 | 20.436 |
| 10.6 | 16.2659 | 0.1204 | -0.3425 | 13.079 | 13.995 | 15.014 | 16.266 | 17.662 | 19.060 | 20.589 |
| 10.8 | 16.3673 | 0.1210 | -0.3476 | 13.148 | 14.073 | 15.102 | 16.367 | 17.780 | 19.196 | 20.746 |
| 11.0 | 16.4708 | 0.1216 | -0.3561 | 13.220 | 14.153 | 15.192 | 16.471 | 17.900 | 19.335 | 20.908 |
| 11.2 | 16.5771 | 0.1222 | -0.3621 | 13.293 | 14.235 | 15.284 | 16.577 | 18.024 | 19.477 | 21.073 |
| 11.4 | 16.6861 | 0.1228 | -0.3687 | 13.368 | 14.319 | 15.378 | 16.686 | 18.151 | 19.624 | 21.243 |
| 11.6 | 16.7981 | 0.1234 | -0.3756 | 13.446 | 14.405 | 15.476 | 16.798 | 18.281 | 19.774 | 21.417 |
| 11.8 | 16.9131 | 0.1241 | -0.3824 | 13.526 | 14.494 | 15.576 | 16.913 | 18.414 | 19.928 | 21.595 |
| 12.0 | 17.0311 | 0.1247 | -0.3900 | 13.608 | 14.586 | 15.679 | 17.031 | 18.551 | 20.086 | 21.779 |
| 12.2 | 17.1523 | 0.1253 | -0.3964 | 13.692 | 14.680 | 15.784 | 17.152 | 18.692 | 20.248 | 21.967 |
| 12.4 | 17.2766 | 0.1259 | -0.4033 | 13.779 | 14.776 | 15.892 | 17.277 | 18.836 | 20.414 | 22.160 |
| 12.6 | 17.4036 | 0.1265 | -0.4106 | 13.868 | 14.875 | 16.003 | 17.404 | 18.983 | 20.584 | 22.357 |
| 12.8 | 17.5334 | 0.1272 | -0.4161 | 13.958 | 14.975 | 16.116 | 17.533 | 19.134 | 20.758 | 22.558 |
| 13.0 | 17.6659 | 0.1278 | -0.4231 | 14.051 | 15.078 | 16.231 | 17.666 | 19.288 | 20.935 | 22.764 |
| 13.2 | 17.8004 | 0.1285 | -0.4309 | 14.145 | 15.183 | 16.349 | 17.800 | 19.444 | 21.115 | 22.974 |
| 13.4 | 17.9362 | 0.1291 | -0.4376 | 14.240 | 15.288 | 16.467 | 17.936 | 19.602 | 21.297 | 23.186 |
| 13.6 | 18.0731 | 0.1298 | -0.4437 | 14.335 | 15.394 | 16.586 | 18.073 | 19.761 | 21.481 | 23.400 |
| 13.8 | 18.2099 | 0.1304 | -0.4503 | 14.430 | 15.500 | 16.705 | 18.210 | 19.920 | 21.665 | 23.615 |
| 14.0 | 18.3459 | 0.1311 | -0.4579 | 14.525 | 15.605 | 16.823 | 18.346 | 20.078 | 21.850 | 23.830 |
| 14.2 | 18.4810 | 0.1317 | -0.4645 | 14.618 | 15.709 | 16.940 | 18.481 | 20.236 | 22.033 | 24.045 |
| 14.4 | 18.6137 | 0.1324 | -0.4714 | 14.709 | 15.811 | 17.055 | 18.614 | 20.391 | 22.214 | 24.258 |
| 14.6 | 18.7437 | 0.1330 | -0.4789 | 14.799 | 15.910 | 17.167 | 18.744 | 20.544 | 22.392 | 24.468 |
| 14.8 | 18.8711 | 0.1337 | -0.4854 | 14.885 | 16.007 | 17.277 | 18.871 | 20.694 | 22.568 | 24.676 |
| 15.0 | 18.9947 | 0.1344 | -0.4910 | 14.968 | 16.100 | 17.383 | 18.995 | 20.840 | 22.739 | 24.879 |
| 15.2 | 19.1138 | 0.1350 | -0.4985 | 15.048 | 16.190 | 17.485 | 19.114 | 20.981 | 22.906 | 25.078 |
| 15.4 | 19.2288 | 0.1357 | -0.5048 | 15.124 | 16.276 | 17.583 | 19.229 | 21.118 | 23.068 | 25.272 |
| 15.6 | 19.3391 | 0.1364 | -0.5127 | 15.197 | 16.358 | 17.677 | 19.339 | 21.250 | 23.226 | 25.462 |
| 15.8 | 19.4453 | 0.1371 | -0.5189 | 15.266 | 16.436 | 17.766 | 19.445 | 21.378 | 23.379 | 25.646 |
| 16.0 | 19.5472 | 0.1378 | -0.5260 | 15.332 | 16.510 | 17.852 | 19.547 | 21.501 | 23.527 | 25.826 |
| 16.2 | 19.6443 | 0.1385 | -0.5329 | 15.393 | 16.581 | 17.933 | 19.644 | 21.619 | 23.670 | 26.001 |
| 16.4 | 19.7372 | 0.1392 | -0.5393 | 15.451 | 16.647 | 18.010 | 19.737 | 21.733 | 23.808 | 26.171 |
| 16.6 | 19.8260 | 0.1398 | -0.5461 | 15.506 | 16.710 | 18.084 | 19.826 | 21.842 | 23.941 | 26.336 |
| 16.8 | 19.9108 | 0.1406 | -0.5523 | 15.557 | 16.769 | 18.153 | 19.911 | 21.947 | 24.071 | 26.497 |
| 17.0 | 19.9913 | 0.1413 | -0.5603 | 15.606 | 16.825 | 18.219 | 19.991 | 22.048 | 24.196 | 26.654 |
| 17.2 | 20.0681 | 0.1419 | -0.5673 | 15.651 | 16.878 | 18.282 | 20.068 | 22.144 | 24.316 | 26.806 |
| 17.4 | 20.1421 | 0.1427 | -0.5729 | 15.693 | 16.927 | 18.341 | 20.142 | 22.238 | 24.434 | 26.955 |
| 17.6 | 20.2124 | 0.1434 | -0.5803 | 15.733 | 16.974 | 18.397 | 20.212 | 22.328 | 24.548 | 27.101 |
| 17.8 | 20.2803 | 0.1441 | -0.5867 | 15.770 | 17.019 | 18.451 | 20.280 | 22.415 | 24.659 | 27.244 |
| 18.0 | 20.3459 | 0.1448 | -0.5935 | 15.806 | 17.061 | 18.503 | 20.346 | 22.500 | 24.767 | 27.385 |
| 18.2 | 20.4090 | 0.1455 | -0.6010 | 15.840 | 17.102 | 18.552 | 20.409 | 22.582 | 24.874 | 27.524 |
| 18.4 | 20.4704 | 0.1463 | -0.6076 | 15.872 | 17.141 | 18.600 | 20.470 | 22.663 | 24.978 | 27.661 |
| 18.6 | 20.5305 | 0.1470 | -0.6148 | 15.904 | 17.178 | 18.646 | 20.531 | 22.742 | 25.082 | 27.798 |
| 18.8 | 20.5893 | 0.1477 | -0.6217 | 15.934 | 17.215 | 18.692 | 20.589 | 22.820 | 25.184 | 27.933 |
| 19.0 | 20.6473 | 0.1485 | -0.6281 | 15.963 | 17.250 | 18.736 | 20.647 | 22.898 | 25.286 | 28.069 |
| 19.2 | 20.7046 | 0.1492 | -0.6349 | 15.992 | 17.285 | 18.780 | 20.705 | 22.974 | 25.387 | 28.204 |
| 19.4 | 20.7618 | 0.1500 | -0.6412 | 16.020 | 17.320 | 18.823 | 20.762 | 23.051 | 25.489 | 28.340 |
| 19.6 | 20.8180 | 0.1507 | -0.6487 | 16.048 | 17.354 | 18.866 | 20.818 | 23.127 | 25.590 | 28.477 |
| 19.8 | 20.8747 | 0.1515 | -0.6553 | 16.076 | 17.388 | 18.909 | 20.875 | 23.203 | 25.692 | 28.614 |

**Table.**
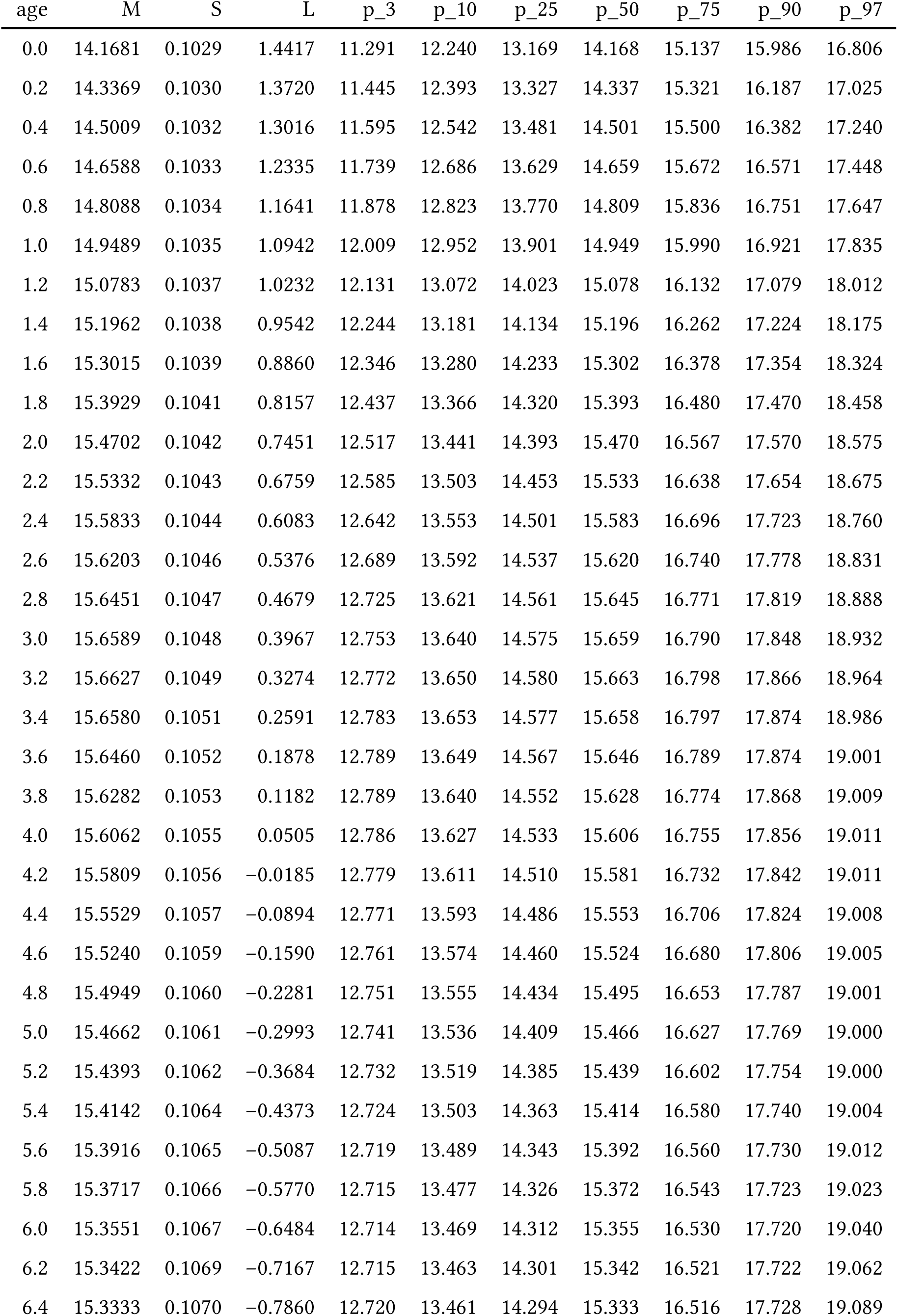
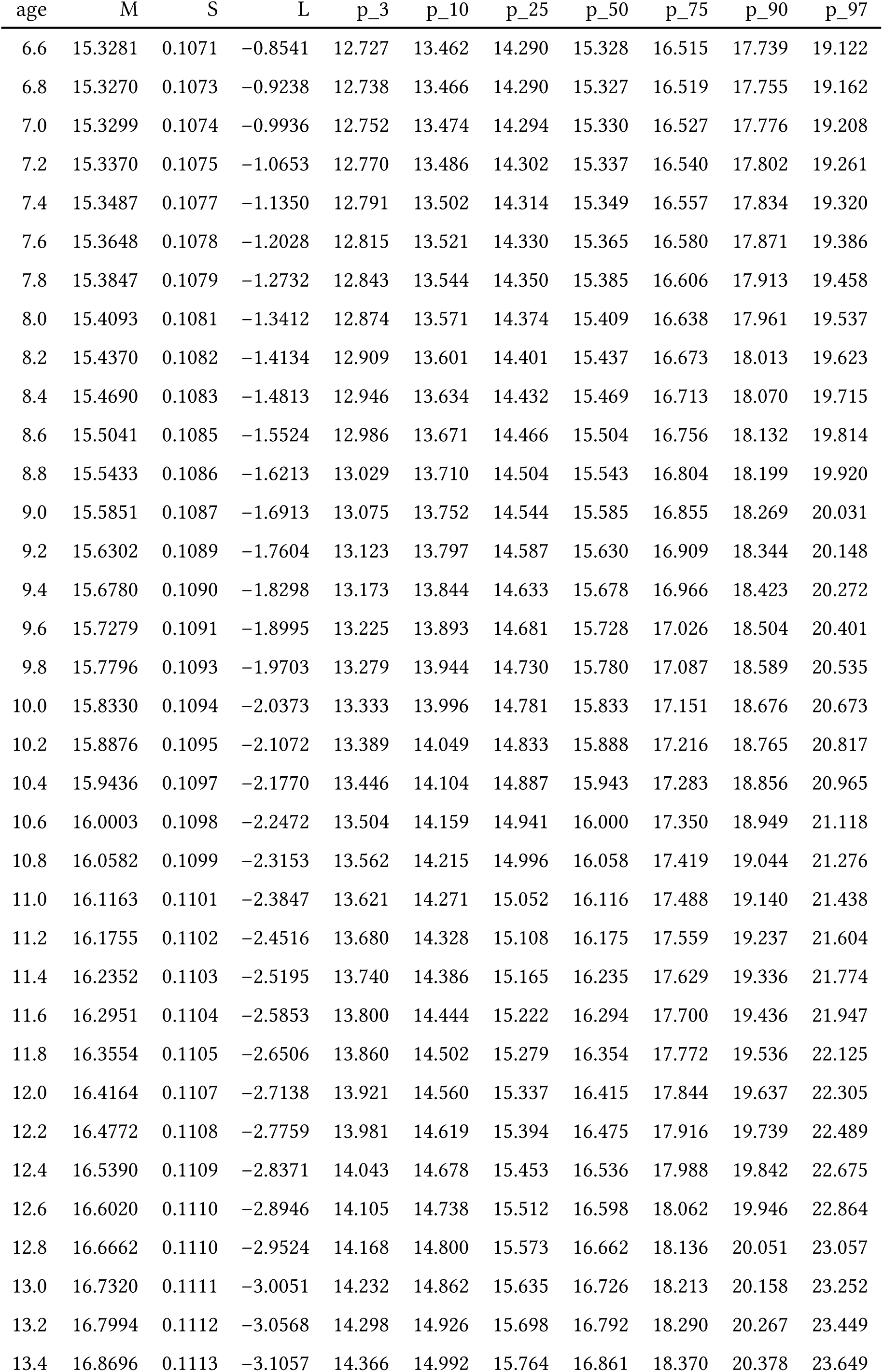
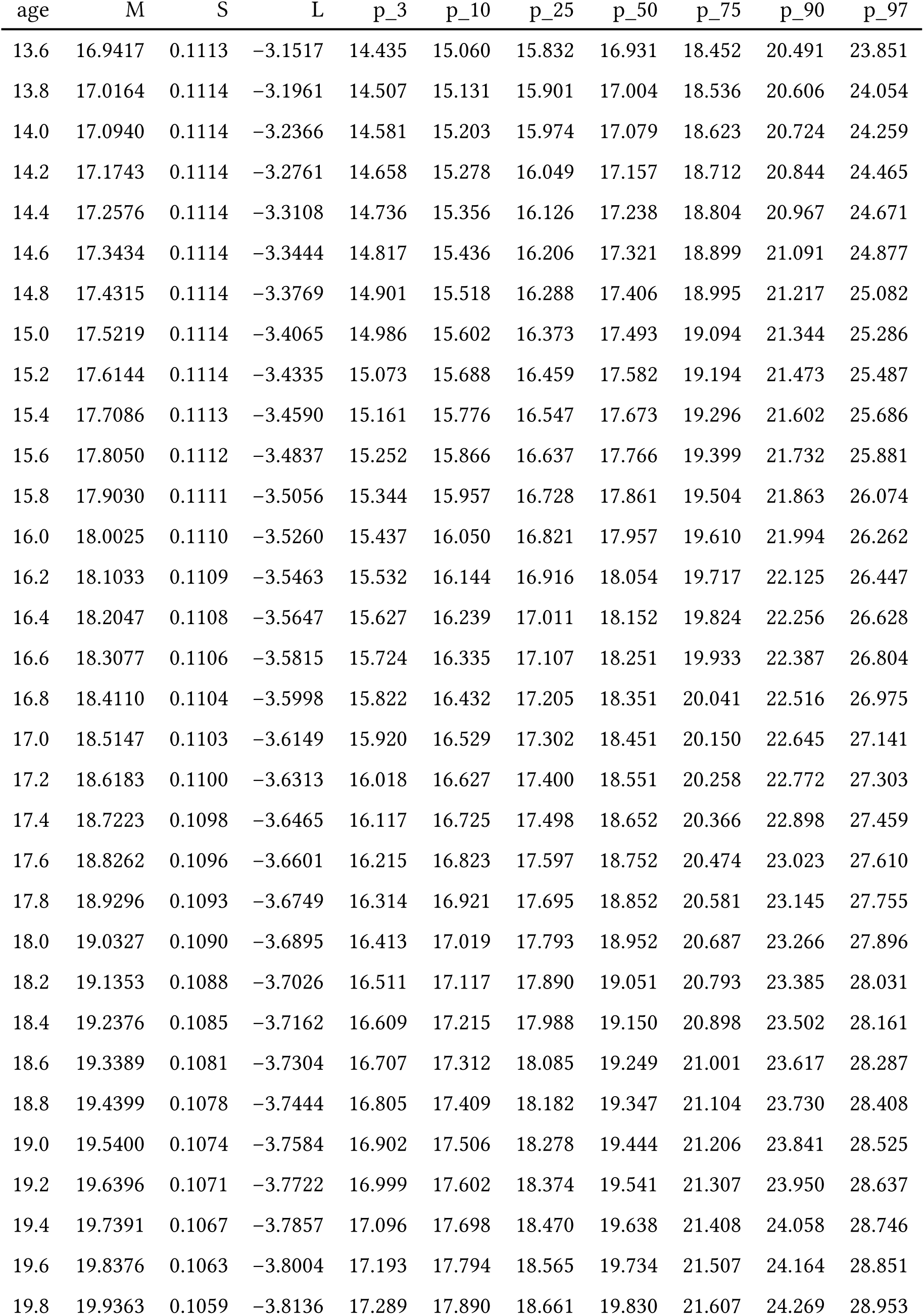
BMI (kg/m²) for males with NS: Estimated M, S, L parameters from centile values by age (3rd, 10th, 25th, 50th, 75th, 90th, 97th)

| age | M | S | L | p_3 | p_10 | p_25 | p_50 | p_75 | p_90 | p_97 |
| --- | --- | --- | --- | --- | --- | --- | --- | --- | --- | --- |
| 0.0 | 14.1681 | 0.1029 | 1.4417 | 11.291 | 12.240 | 13.169 | 14.168 | 15.137 | 15.986 | 16.806 |
| 0.2 | 14.3369 | 0.1030 | 1.3720 | 11.445 | 12.393 | 13.327 | 14.337 | 15.321 | 16.187 | 17.025 |
| 0.4 | 14.5009 | 0.1032 | 1.3016 | 11.595 | 12.542 | 13.481 | 14.501 | 15.500 | 16.382 | 17.240 |
| 0.6 | 14.6588 | 0.1033 | 1.2335 | 11.739 | 12.686 | 13.629 | 14.659 | 15.672 | 16.571 | 17.448 |
| 0.8 | 14.8088 | 0.1034 | 1.1641 | 11.878 | 12.823 | 13.770 | 14.809 | 15.836 | 16.751 | 17.647 |
| 1.0 | 14.9489 | 0.1035 | 1.0942 | 12.009 | 12.952 | 13.901 | 14.949 | 15.990 | 16.921 | 17.835 |
| 1.2 | 15.0783 | 0.1037 | 1.0232 | 12.131 | 13.072 | 14.023 | 15.078 | 16.132 | 17.079 | 18.012 |
| 1.4 | 15.1962 | 0.1038 | 0.9542 | 12.244 | 13.181 | 14.134 | 15.196 | 16.262 | 17.224 | 18.175 |
| 1.6 | 15.3015 | 0.1039 | 0.8860 | 12.346 | 13.280 | 14.233 | 15.302 | 16.378 | 17.354 | 18.324 |
| 1.8 | 15.3929 | 0.1041 | 0.8157 | 12.437 | 13.366 | 14.320 | 15.393 | 16.480 | 17.470 | 18.458 |
| 2.0 | 15.4702 | 0.1042 | 0.7451 | 12.517 | 13.441 | 14.393 | 15.470 | 16.567 | 17.570 | 18.575 |
| 2.2 | 15.5332 | 0.1043 | 0.6759 | 12.585 | 13.503 | 14.453 | 15.533 | 16.638 | 17.654 | 18.675 |
| 2.4 | 15.5833 | 0.1044 | 0.6083 | 12.642 | 13.553 | 14.501 | 15.583 | 16.696 | 17.723 | 18.760 |
| 2.6 | 15.6203 | 0.1046 | 0.5376 | 12.689 | 13.592 | 14.537 | 15.620 | 16.740 | 17.778 | 18.831 |
| 2.8 | 15.6451 | 0.1047 | 0.4679 | 12.725 | 13.621 | 14.561 | 15.645 | 16.771 | 17.819 | 18.888 |
| 3.0 | 15.6589 | 0.1048 | 0.3967 | 12.753 | 13.640 | 14.575 | 15.659 | 16.790 | 17.848 | 18.932 |
| 3.2 | 15.6627 | 0.1049 | 0.3274 | 12.772 | 13.650 | 14.580 | 15.663 | 16.798 | 17.866 | 18.964 |
| 3.4 | 15.6580 | 0.1051 | 0.2591 | 12.783 | 13.653 | 14.577 | 15.658 | 16.797 | 17.874 | 18.986 |
| 3.6 | 15.6460 | 0.1052 | 0.1878 | 12.789 | 13.649 | 14.567 | 15.646 | 16.789 | 17.874 | 19.001 |
| 3.8 | 15.6282 | 0.1053 | 0.1182 | 12.789 | 13.640 | 14.552 | 15.628 | 16.774 | 17.868 | 19.009 |
| 4.0 | 15.6062 | 0.1055 | 0.0505 | 12.786 | 13.627 | 14.533 | 15.606 | 16.755 | 17.856 | 19.011 |
| 4.2 | 15.5809 | 0.1056 | -0.0185 | 12.779 | 13.611 | 14.510 | 15.581 | 16.732 | 17.842 | 19.011 |
| 4.4 | 15.5529 | 0.1057 | -0.0894 | 12.771 | 13.593 | 14.486 | 15.553 | 16.706 | 17.824 | 19.008 |
| 4.6 | 15.5240 | 0.1059 | -0.1590 | 12.761 | 13.574 | 14.460 | 15.524 | 16.680 | 17.806 | 19.005 |
| 4.8 | 15.4949 | 0.1060 | -0.2281 | 12.751 | 13.555 | 14.434 | 15.495 | 16.653 | 17.787 | 19.001 |
| 5.0 | 15.4662 | 0.1061 | -0.2993 | 12.741 | 13.536 | 14.409 | 15.466 | 16.627 | 17.769 | 19.000 |
| 5.2 | 15.4393 | 0.1062 | -0.3684 | 12.732 | 13.519 | 14.385 | 15.439 | 16.602 | 17.754 | 19.000 |
| 5.4 | 15.4142 | 0.1064 | -0.4373 | 12.724 | 13.503 | 14.363 | 15.414 | 16.580 | 17.740 | 19.004 |
| 5.6 | 15.3916 | 0.1065 | -0.5087 | 12.719 | 13.489 | 14.343 | 15.392 | 16.560 | 17.730 | 19.012 |
| 5.8 | 15.3717 | 0.1066 | -0.5770 | 12.715 | 13.477 | 14.326 | 15.372 | 16.543 | 17.723 | 19.023 |
| 6.0 | 15.3551 | 0.1067 | -0.6484 | 12.714 | 13.469 | 14.312 | 15.355 | 16.530 | 17.720 | 19.040 |
| 6.2 | 15.3422 | 0.1069 | -0.7167 | 12.715 | 13.463 | 14.301 | 15.342 | 16.521 | 17.722 | 19.062 |
| 6.4 | 15.3333 | 0.1070 | -0.7860 | 12.720 | 13.461 | 14.294 | 15.333 | 16.516 | 17.728 | 19.089 |
| 6.6 | 15.3281 | 0.1071 | -0.8541 | 12.727 | 13.462 | 14.290 | 15.328 | 16.515 | 17.739 | 19.122 |
| 6.8 | 15.3270 | 0.1073 | -0.9238 | 12.738 | 13.466 | 14.290 | 15.327 | 16.519 | 17.755 | 19.162 |
| 7.0 | 15.3299 | 0.1074 | -0.9936 | 12.752 | 13.474 | 14.294 | 15.330 | 16.527 | 17.776 | 19.208 |
| 7.2 | 15.3370 | 0.1075 | -1.0653 | 12.770 | 13.486 | 14.302 | 15.337 | 16.540 | 17.802 | 19.261 |
| 7.4 | 15.3487 | 0.1077 | -1.1350 | 12.791 | 13.502 | 14.314 | 15.349 | 16.557 | 17.834 | 19.320 |
| 7.6 | 15.3648 | 0.1078 | -1.2028 | 12.815 | 13.521 | 14.330 | 15.365 | 16.580 | 17.871 | 19.386 |
| 7.8 | 15.3847 | 0.1079 | -1.2732 | 12.843 | 13.544 | 14.350 | 15.385 | 16.606 | 17.913 | 19.458 |
| 8.0 | 15.4093 | 0.1081 | -1.3412 | 12.874 | 13.571 | 14.374 | 15.409 | 16.638 | 17.961 | 19.537 |
| 8.2 | 15.4370 | 0.1082 | -1.4134 | 12.909 | 13.601 | 14.401 | 15.437 | 16.673 | 18.013 | 19.623 |
| 8.4 | 15.4690 | 0.1083 | -1.4813 | 12.946 | 13.634 | 14.432 | 15.469 | 16.713 | 18.070 | 19.715 |
| 8.6 | 15.5041 | 0.1085 | -1.5524 | 12.986 | 13.671 | 14.466 | 15.504 | 16.756 | 18.132 | 19.814 |
| 8.8 | 15.5433 | 0.1086 | -1.6213 | 13.029 | 13.710 | 14.504 | 15.543 | 16.804 | 18.199 | 19.920 |
| 9.0 | 15.5851 | 0.1087 | -1.6913 | 13.075 | 13.752 | 14.544 | 15.585 | 16.855 | 18.269 | 20.031 |
| 9.2 | 15.6302 | 0.1089 | -1.7604 | 13.123 | 13.797 | 14.587 | 15.630 | 16.909 | 18.344 | 20.148 |
| 9.4 | 15.6780 | 0.1090 | -1.8298 | 13.173 | 13.844 | 14.633 | 15.678 | 16.966 | 18.423 | 20.272 |
| 9.6 | 15.7279 | 0.1091 | -1.8995 | 13.225 | 13.893 | 14.681 | 15.728 | 17.026 | 18.504 | 20.401 |
| 9.8 | 15.7796 | 0.1093 | -1.9703 | 13.279 | 13.944 | 14.730 | 15.780 | 17.087 | 18.589 | 20.535 |
| 10.0 | 15.8330 | 0.1094 | -2.0373 | 13.333 | 13.996 | 14.781 | 15.833 | 17.151 | 18.676 | 20.673 |
| 10.2 | 15.8876 | 0.1095 | -2.1072 | 13.389 | 14.049 | 14.833 | 15.888 | 17.216 | 18.765 | 20.817 |
| 10.4 | 15.9436 | 0.1097 | -2.1770 | 13.446 | 14.104 | 14.887 | 15.943 | 17.283 | 18.856 | 20.965 |
| 10.6 | 16.0003 | 0.1098 | -2.2472 | 13.504 | 14.159 | 14.941 | 16.000 | 17.350 | 18.949 | 21.118 |
| 10.8 | 16.0582 | 0.1099 | -2.3153 | 13.562 | 14.215 | 14.996 | 16.058 | 17.419 | 19.044 | 21.276 |
| 11.0 | 16.1163 | 0.1101 | -2.3847 | 13.621 | 14.271 | 15.052 | 16.116 | 17.488 | 19.140 | 21.438 |
| 11.2 | 16.1755 | 0.1102 | -2.4516 | 13.680 | 14.328 | 15.108 | 16.175 | 17.559 | 19.237 | 21.604 |
| 11.4 | 16.2352 | 0.1103 | -2.5195 | 13.740 | 14.386 | 15.165 | 16.235 | 17.629 | 19.336 | 21.774 |
| 11.6 | 16.2951 | 0.1104 | -2.5853 | 13.800 | 14.444 | 15.222 | 16.294 | 17.700 | 19.436 | 21.947 |
| 11.8 | 16.3554 | 0.1105 | -2.6506 | 13.860 | 14.502 | 15.279 | 16.354 | 17.772 | 19.536 | 22.125 |
| 12.0 | 16.4164 | 0.1107 | -2.7138 | 13.921 | 14.560 | 15.337 | 16.415 | 17.844 | 19.637 | 22.305 |
| 12.2 | 16.4772 | 0.1108 | -2.7759 | 13.981 | 14.619 | 15.394 | 16.475 | 17.916 | 19.739 | 22.489 |
| 12.4 | 16.5390 | 0.1109 | -2.8371 | 14.043 | 14.678 | 15.453 | 16.536 | 17.988 | 19.842 | 22.675 |
| 12.6 | 16.6020 | 0.1110 | -2.8946 | 14.105 | 14.738 | 15.512 | 16.598 | 18.062 | 19.946 | 22.864 |
| 12.8 | 16.6662 | 0.1110 | -2.9524 | 14.168 | 14.800 | 15.573 | 16.662 | 18.136 | 20.051 | 23.057 |
| 13.0 | 16.7320 | 0.1111 | -3.0051 | 14.232 | 14.862 | 15.635 | 16.726 | 18.213 | 20.158 | 23.252 |
| 13.2 | 16.7994 | 0.1112 | -3.0568 | 14.298 | 14.926 | 15.698 | 16.792 | 18.290 | 20.267 | 23.449 |
| 13.4 | 16.8696 | 0.1113 | -3.1057 | 14.366 | 14.992 | 15.764 | 16.861 | 18.370 | 20.378 | 23.649 |
| 13.6 | 16.9417 | 0.1113 | -3.1517 | 14.435 | 15.060 | 15.832 | 16.931 | 18.452 | 20.491 | 23.851 |
| 13.8 | 17.0164 | 0.1114 | -3.1961 | 14.507 | 15.131 | 15.901 | 17.004 | 18.536 | 20.606 | 24.054 |
| 14.0 | 17.0940 | 0.1114 | -3.2366 | 14.581 | 15.203 | 15.974 | 17.079 | 18.623 | 20.724 | 24.259 |
| 14.2 | 17.1743 | 0.1114 | -3.2761 | 14.658 | 15.278 | 16.049 | 17.157 | 18.712 | 20.844 | 24.465 |
| 14.4 | 17.2576 | 0.1114 | -3.3108 | 14.736 | 15.356 | 16.126 | 17.238 | 18.804 | 20.967 | 24.671 |
| 14.6 | 17.3434 | 0.1114 | -3.3444 | 14.817 | 15.436 | 16.206 | 17.321 | 18.899 | 21.091 | 24.877 |
| 14.8 | 17.4315 | 0.1114 | -3.3769 | 14.901 | 15.518 | 16.288 | 17.406 | 18.995 | 21.217 | 25.082 |
| 15.0 | 17.5219 | 0.1114 | -3.4065 | 14.986 | 15.602 | 16.373 | 17.493 | 19.094 | 21.344 | 25.286 |
| 15.2 | 17.6144 | 0.1114 | -3.4335 | 15.073 | 15.688 | 16.459 | 17.582 | 19.194 | 21.473 | 25.487 |
| 15.4 | 17.7086 | 0.1113 | -3.4590 | 15.161 | 15.776 | 16.547 | 17.673 | 19.296 | 21.602 | 25.686 |
| 15.6 | 17.8050 | 0.1112 | -3.4837 | 15.252 | 15.866 | 16.637 | 17.766 | 19.399 | 21.732 | 25.881 |
| 15.8 | 17.9030 | 0.1111 | -3.5056 | 15.344 | 15.957 | 16.728 | 17.861 | 19.504 | 21.863 | 26.074 |
| 16.0 | 18.0025 | 0.1110 | -3.5260 | 15.437 | 16.050 | 16.821 | 17.957 | 19.610 | 21.994 | 26.262 |
| 16.2 | 18.1033 | 0.1109 | -3.5463 | 15.532 | 16.144 | 16.916 | 18.054 | 19.717 | 22.125 | 26.447 |
| 16.4 | 18.2047 | 0.1108 | -3.5647 | 15.627 | 16.239 | 17.011 | 18.152 | 19.824 | 22.256 | 26.628 |
| 16.6 | 18.3077 | 0.1106 | -3.5815 | 15.724 | 16.335 | 17.107 | 18.251 | 19.933 | 22.387 | 26.804 |
| 16.8 | 18.4110 | 0.1104 | -3.5998 | 15.822 | 16.432 | 17.205 | 18.351 | 20.041 | 22.516 | 26.975 |
| 17.0 | 18.5147 | 0.1103 | -3.6149 | 15.920 | 16.529 | 17.302 | 18.451 | 20.150 | 22.645 | 27.141 |
| 17.2 | 18.6183 | 0.1100 | -3.6313 | 16.018 | 16.627 | 17.400 | 18.551 | 20.258 | 22.772 | 27.303 |
| 17.4 | 18.7223 | 0.1098 | -3.6465 | 16.117 | 16.725 | 17.498 | 18.652 | 20.366 | 22.898 | 27.459 |
| 17.6 | 18.8262 | 0.1096 | -3.6601 | 16.215 | 16.823 | 17.597 | 18.752 | 20.474 | 23.023 | 27.610 |
| 17.8 | 18.9296 | 0.1093 | -3.6749 | 16.314 | 16.921 | 17.695 | 18.852 | 20.581 | 23.145 | 27.755 |
| 18.0 | 19.0327 | 0.1090 | -3.6895 | 16.413 | 17.019 | 17.793 | 18.952 | 20.687 | 23.266 | 27.896 |
| 18.2 | 19.1353 | 0.1088 | -3.7026 | 16.511 | 17.117 | 17.890 | 19.051 | 20.793 | 23.385 | 28.031 |
| 18.4 | 19.2376 | 0.1085 | -3.7162 | 16.609 | 17.215 | 17.988 | 19.150 | 20.898 | 23.502 | 28.161 |
| 18.6 | 19.3389 | 0.1081 | -3.7304 | 16.707 | 17.312 | 18.085 | 19.249 | 21.001 | 23.617 | 28.287 |
| 18.8 | 19.4399 | 0.1078 | -3.7444 | 16.805 | 17.409 | 18.182 | 19.347 | 21.104 | 23.730 | 28.408 |
| 19.0 | 19.5400 | 0.1074 | -3.7584 | 16.902 | 17.506 | 18.278 | 19.444 | 21.206 | 23.841 | 28.525 |
| 19.2 | 19.6396 | 0.1071 | -3.7722 | 16.999 | 17.602 | 18.374 | 19.541 | 21.307 | 23.950 | 28.637 |
| 19.4 | 19.7391 | 0.1067 | -3.7857 | 17.096 | 17.698 | 18.470 | 19.638 | 21.408 | 24.058 | 28.746 |
| 19.6 | 19.8376 | 0.1063 | -3.8004 | 17.193 | 17.794 | 18.565 | 19.734 | 21.507 | 24.164 | 28.851 |
| 19.8 | 19.9363 | 0.1059 | -3.8136 | 17.289 | 17.890 | 18.661 | 19.830 | 21.607 | 24.269 | 28.953 |

## Notes

### Competing Interest Statement

JHC is the developer of the PediTools website of growth calculators which generates revenue from Google Adsense; Google Adsense had no input on the content presented.

### Funding Statement

This study did not receive any funding.

### Author Declarations

All data used was obtained from the Supplementary Materials from Cappa et al 2024

### Summary of Updates

Add full R code for the LMS estimation function (including an example of use) to the Supplementary Materials

